# Lipid-regulatory mechanisms drive cerebrovascular disease in asymptomatic individuals at low risk for late-life dementia

**DOI:** 10.1101/2024.05.08.24307060

**Authors:** Patricia Genius, Blanca Rodríguez-Fernández, Carolina Minguillon, Anna Brugulat-Serrat, Jordi Huguet, Manel Esteller, Carole H. Sudre, Marta Cortés Canteli, Catarina Tristão-Pereira, Inés García Lunar, Arcadi Navarro, Juan Domingo Gispert, Natalia Vilor-Tejedor, ALFA study

**Affiliations:** Barcelonaβeta Brain Research Center (BBRC), Pasqual Maragall Foundation, C/ Wellington 30, Sant Martí, 08005, Barcelona, Spain; IMIM (Hospital del Mar Medical Research Institute), C/ Dr. Aiguader 88, 08003, Barcelona, Spain; Centre for Genomic Regulation (CRG), Barcelona Institute of Science and Technology (BIST), C/ Dr. Aiguader 88, 08003, Barcelona, Spain; Autonomous University of Barcelona, Barcelona, Plaça Cívica, 08193 Bellaterra, Barcelona, Spain; Centro de Investigación Biomédica en Red de Fragilidad y Envejecimiento Saludable (CIBERFES), Instituto de Salud Carlos III, C/ Monforte de Lemos 3-5. Pabellón 11, Planta 0, 28029, Madrid, Spain; Global Brain Health Institute, 675 Nelson Rising Lane, Suite 190. San Francisco, CA 94158, University of California, USA; Josep Carreras Leukaemia Research Institute (IJC), Ctra de Can Ruti, Camí de les Escoles, s/n, 08916 Badalona, Barcelona, Catalonia, Spain; Institució Catalana de Recerca i Estudis Avançats (ICREA), Passeig Lluís Companys 23, 08010, Barcelona, Catalonia, Spain; Physiological Sciences Department, School of Medicine and Health Sciences, University of Barcelona (UB), C/ Casanova 143, 08014, Barcelona, Catalonia, Spain; Centro de Investigación Biomédica en Red Cancer (CIBERONC), Av. Monforte de Lemos, 3-5, Pabellón 11, Planta 0, 28029 Madrid, Spain; MRC Unit for Lifelong Health and Ageing, Department of Population Science and Experimental Medicine, Gower Street, University College London, UK; Centre for Medical Image Computing, Department of Computer Science, Gower Street, University College London, UK; Department of Neurodegenerative Disease, The Dementia Research Centre, UCL Queen Square Institute of Neurology, Gower Street, University College London, UK; School of Biomedical Engineering and Imaging Sciences, King’s College London, Strand, London, UK; Centro Nacional de Investigaciones Cardiovasculares (CNIC), C./ Melchor Fernández Almagro 3, Fuencarral-El Pardo, 28029 Madrid, Spain; Cardiology Department, University Hospital La Moraleja, Madrid, Spain. CIBER de Enfermedades Cardiovasculares (CIBERCV), Madrid, Spain; Pompeu Fabra University, C/ de Ramon Trias Fargas 25-27, Sant Martí, 08005, Barcelona Barcelona, Spain; Institute of Evolutionary Biology (CSIC-UPF), Department of Experimental and Health Sciences, Universitat Pompeu Fabra, Pg. Marítim de la Barceloneta 37, Ciutat Vella, 08003, Barcelona, Spain; Centro de Investigación Biomédica en Red Bioingeniería, Biomateriales y Nanomedicina, Instituto de Salud Carlos III, Av. Monforte de Lemos 3-5, Pabellón 11, Planta 0, 28029 Madrid, Spain; Radboud University Nijmegen Medical Center, Department of Genetics, Geert Grooteplein Zuid 10, 6525 GA Nijmegen, the Netherlands

**Keywords:** white matter hyperintensities, polygenic risk scores, risk factors, cerebrovascular disease, dementia

## Abstract

**Background:** Cerebrovascular lesions, particularly white matter hyperintensities (WMH), are often found in middle-aged individuals with a low cardiovascular risk profile. Understanding modifiable mechanisms leading to cerebrovascular disease is fundamental for implementing preventive strategies. This study aimed to elucidate the biological mechanisms underlying the presence of WMH in cognitively unimpaired (CU) middle-aged individuals.

**Methods:** We included 1,072 CU participants from the ALFA study with a low cardiovascular risk profile for late-life dementia based on the CAIDE score. We assessed genetic predisposition to WMH using polygenic scoring (PRS_WMH_). Covariate-adjusted Spearman’s rank correlation tests evaluated the association between the PRS_WMH_ and white matter hyperintensities volumes (WMHV). A logistic regression model was performed to explore the association between the PRS_WMH_ and WMH severity, as measured with the Fazekas score. An enrichment analysis of the PRS-annotated genes unveiled the biological mechanisms leading to WMH burden. Group-specific effects were explored based on dementia-related cardiovascular risk factors.

**Results:** Genetic predisposition to WMH was associated with larger WMHV, even after controlling for confounders, but was not associated with WMH severity. Lipid-related biological processes were driving WMH genetic risk. Individuals genetically predisposed to WMH, who displayed larger WMHV, were either hypercholesterolemic, older than 55 or with lower educational attainment.

**Interpretation:** Lipid-related mechanisms contribute to WMH in individuals at low cardiovascular risk for late-life dementia. These individuals should be considered for lifestyle- and lipid-modifying therapies to prevent dementia later in life.

**Funding:** “La Caixa” Foundation, the TriBEKa Imaging Platform, the Universities and Research Secretariat of the Catalan Government, the Spanish Research Agency.

## Introduction

White matter hyperintensities (WMH) are brain white matter lesions commonly detected in the brain of elderly individuals through magnetic resonance imaging (MRI)^1^. WMH are well-established markers of cerebral small vessel disease (SVS), but different pathophysiological pathways have been proposed to explain their existence.^2^ These are mainly categorized into vascular (e.g. due to ischemia/hypoperfusion) and non-vascular (e.g. due to gliosis, axonal loss) origin. WMH contribute to cognitive impairment and neurodegeneration^3^, and increase the risk of stroke, cognitive decline and dementia in the general population.^3-4^ Several modifiable and non-modifiable risk factors have been associated with larger white matter hyperintensities volumes (WMHV).^5^ Age and hypertension are the main risk factors for WMH^6-7^, but other cardiovascular risk factors^8^ (CVRF) also play a significant role. Nonetheless, the analysis of the cardiovascular risk profile may be insufficient for identifying individuals who may develop WMH lesions^9^. Multiple studies have suggested considering the role of genetic factors in WMH, given the strong heritability of this condition (∼40% to 70%)^5^. For instance, several studies have found significant associations between the *Apolipoprotein E* gene (*APOE*) *ε4* allele and larger WMHV. ^8-10-11-12^ However, contrary to the previous findings, other studies have suggested that there may be no association or even a reverse association between *APOE-ε4* and white matter lesions in Alzheimer’s disease (AD) patients^13^. These results highlight the potential importance of heterogeneous vascular risk factors in developing WMH in the absence of the *APOE-ε4* genotype. Beyond *APOE*, genome-wide association studies (GWAS) have identified novel genetic variants associated with WMH burden and microstructure^14-15^, which suggested a polygenic architecture of WMH.^16^ Moreover, the development of WMH has been linked to genetic *loci* that are implicated in inflammatory and glial proliferative pathways, contained in genes that are also implicated in AD, intracerebral hemorrhage, neuroinflammatory diseases and glioma, as well as blood pressure regulation^14^. Specifically, recent studies^17^ found that genes associated with WMHV closer to the ventricles (*i*.*e*. periventricular WMH), which are highly linked to cognitive decline and elevated systolic blood pressure, were mostly involved in vascular function and vascular disease. On the other hand, genes associated with more external WMHV (*i*.*e*. deep WMH), which are highly linked to obesity, gait dysfunction and arterial hypertension, were mostly involved in astrocyte and neuronal function. These findings suggest separate causes for regional WMH. Therefore, exploring the genetic basis of WMH as well as the gene-risk factors interaction can not only increase the understanding of the mechanisms behind WMH but can also lead to a better identification of vulnerable populations who may not be identified through traditional clinical screening.

The present study aimed to explore the biological mechanisms underlying the presence of WMH in cognitively unimpaired (CU) middle-aged individuals at low cardiovascular risk for late-life dementia. First, we assessed whether the genetic predisposition to WMH, assessed through the polygenic risk score of WMH (PRS_WMH_), was a proxy for larger global and regional WMHV as well as WMH severity in the sample. Second, we explored the biological mechanisms associated with larger WMHV by performing an enrichment analysis of the genes annotated to the genetic variants included in the PRS_WMH_. Third, we explored in which specific groups of individuals, WMH-related genetic factors were predisposing them to larger WMHV.

## Methods

### Study Population

Participants of this study are part of the ALFA study (ALzheimer’s and FAmilies). The present study included CU participants with available information on WMHV, genetic data and available information on AD-related risk factors (N=1,072) [Supplementary Figure 1]. For a full detailed description of the ALFA study see^18^. The study was approved by an independent Ethics Committee ‘Parc de Salut Mar’, Barcelona, and is registered at Clinicaltrials.gov (Identifier: NCT02485730).

### Sociodemographic, lifestyle and clinical factors

Sociodemographic, lifestyle and clinical factors were collected. Participants were considered hypertensives when (i) measured systolic blood pressure (SBP) was above 140 mmHg, (ii) they were under an antihypertensive treatment or (iii) it was self-reported. Body mass index (BMI) was derived from the height and weight and individuals were classified as underweight (BMI<18·5), normal weight (18·5≤BMI≤24·9), overweight (25≤BMI≤29·9), and obese (BMI≥30). Physical activity was measured using the Spanish short version of Minnesota Leisure Time Physical Activity Questionnaire^19^. Since total cholesterol levels were not available for all the participants, individuals were classified as hypercholesterolemic when they self-reported either the presence of dyslipidemia or the use of medication. Educational attainment was determined by assessing the number of years of formal education completed.

### Cardiovascular risk for late-life dementia: CAIDE score

The aforementioned risk factors were included in the Cardiovascular Risk Factors, Aging, and Incidence of Dementia I (CAIDE-I) risk score predicting 20-year risk for late-life dementia [Supplementary Figure 2]. In the standard protocol for the CAIDE score computation^20^, variables are dichotomized or categorized in tertiles based on standard cutoffs. In ALFA, participants were assigned a score depending on the group they belonged to for each one of the vascular risk factors^8^[Supplementary Table 1]. In the current study, individuals scoring below 10 (CAIDE-I score <10)^20^ were classified as low-risk for developing dementia late in life and were included in the final sample of the study. Individuals with a CAIDE-I score >9 were excluded from the study.

### Genetic data acquisition, quality control and imputation

DNA was obtained from blood samples through a salting out protocol. Genotyping was performed with the Illumina Infinium Neuro Consortium (NeuroChip) Array (build GRCh37/hg19). Quality control procedure was performed using PLINK software. Imputation was performed using the Michigan Imputation Server with the haplotype Reference Consortium Panel (HRC r1.1 2016)^21^ following default parameters and established guidelines. A full description of the genotyping, quality control and imputation procedures is available elsewhere^22^.

### Polygenicity of White Matter Hyperintensities

Polygenicity of WMH was calculated using the PRSice version 2 tool.^23^ Summary statistics from a recent GWAS for WMH^15^ were obtained to compute the PRS-WMH. We applied the clumping and thresholding method, which consisted on retaining the single nucleotide polymorphisms (SNPs) with the smallest p-value in each 250 kb window and removing the SNPs that were in linkage disequilibrium (r2 > 0·1). After removing highly correlated SNPs, the PRS was performed at different thresholds of SNPs inclusions, based on the p-value of the summary statistics from the GWAS. We worked with the PRS that included SNPs with a p-value<5·10^-6^ [Supplementary Table 2]. The PRS-WMH was computed by adding up the alleles carried by participants, weighted by the SNP allele effect size from the GWAS and normalizing by the total number of alleles [Supplementary Table 3].

### *Magnetic resonance imaging acquisition* and *WMH volume quantification*

MRIs were acquired on a Philips Ingenia CX 3T MRI scanner. The MRI protocol included a high resolution a 3D T1-weighted sequence (voxel size = 0·75 x 0·75 x 0·75 mm, TR/TE = 9·9/4·6 ms, Flip Angle = 8º) and a 3D fluid attenuation inversion recovery (T2-FLAIR) scan (voxel size = 1 x 1 x 1 mm, TR/TE/IR = 5000/312/1700 ms). All scans were visually assessed for quality and incidental findings by a trained neuroradiologist.^24^ A Bayesian algorithm was used to quantify WMHV (mm^3^) from T1-weighted, and T2-FLAIR MRI scans.^25^ Regional WMHV were quantified at five different distances from the ventricles and for six different regions in both the right and left hemispheres. To reduce the dimensionality, WMH were categorized into three subtypes based on the distance to the ventricles (periventricular, deep and juxtacortical), and computed the average volume between both hemispheres. Volumes were adjusted for total intracranial volume (TIV). Finally, a multiple factor analysis was performed to obtain a composite for each type of regional WMH (PC1 periventricular WMH, PC1 deep WMH and PC1 juxtacortical WMH) [Supplementary Methods]. Global WMHV was adjusted for TIV. Additionally, all MRIs were visually assessed by a trained neuroradiologist who was blinded to the *APOE* genotype of the participants. Images were rated using modifications of the Fazekas Scale ^26^, which separately categorizes the severity of WMH lesions on a scale from 0 to 3 (0, none or a single punctate WMH lesion; 1, multiple punctate lesions; 2, beginning confluency of lesions (bridging); and 3, large confluent lesions). WMH severity was defined based on the cut-off of 2 (Fazekas score <2: non-pathological WMH, Fazekas score ≥ 2: pathological WMH).

### Statistical analysis

A descriptive analysis of the sample of the study was performed using chi-square tests for categorical variables and parametric (t-test) and non-parametric tests (Wilcox test) for continuous normally and non-normally distributed variables, as appropriate. As both WMH and log-transformed WMHV were asymmetrically distributed and linearity of the residuals was not guaranteed, covariate-adjusted Spearman’s rank correlation test^27^ was performed to investigate whether the genetic predisposition to WMH was associated with larger WMHV, both globally and regionally. Partial correlations were adjusted for age and sex. Sensitivity analyses were performed adjusting for the CAIDE-I score, hypertension status and WMH severity (pathological vs non-pathological). We also performed a logistic regression model to explore the association between the genetic predisposition to WMH and WMH severity adjusting for age, sex and hypertension. Genetic variants associated with WMH were annotated to their nearest genes (distance of 10kb) using *biomaRt* package in R [Supplementary Table 4]. Next, enrichment analysis was conducted, using *clusterProfiler 4*.*6* package^28^ in R, to identify the primary biological pathways (Gene Ontology; GO terms) linked with the genes that confer higher risk of WMH. We used the one-sided version of Fisher’s exact test to determine whether known biological functions were overrepresented in the gene list and calculate the probability of observing a set of genes in a particular biological pathway by chance. The enrichment analysis was performed (i) including all WMH-related SNPs from the GWAS of reference with p-value<5·10^-6^, and (ii) including all SNPs from the aforementioned list that remained after the clumping. We conducted subsequent *post-hoc* analyses to assess the correlation between PRS_WMH_ and global WMHV, stratifying by CAIDE-I components. For each component, individuals were classified into more homogeneous groups. Individuals who belonged to the “underweight” group (n=3) were excluded and a binary variable was created to classify individuals into obese and non-obese groups (cut-off point BMI≥30). Individuals were classified as being active (≥150 minutes) or inactive (<150 minutes) based on the time of physical activity per week. Based on the distribution of the age, participants were divided into three groups: (i) 45-54 years old group, (ii) 55-64 years old group and (iii) 65-77 years old group. Similarly, individuals were classified into low (0-6 years), intermediate (7-9 years) and high (≥10 years) education groups. Models were adjusted for age and sex when required. All the analyses were performed using the R software version 4.2.2.

## Results

### Descriptive analysis

The study sample included 1,072 individuals at low risk to develop dementia late in life, where 64·3% (N=689) were women. The median age of the sample was 59 years [IQR 53-64] and the median value for total years of education was equal to 14 [IQR 11-17] [Table 1]. We observed significant differences when comparing between non-pathological and pathological groups based on WMH severity. In the group displaying pathological WMH, individuals were older (p-value<0·001), had higher risk to develop dementia late in life based on the CAIDE score (p-value<0·001), had a higher percentage of hypertensive (p-value<0·001) and diabetic (p-value=0·012) individuals. They also displayed higher TIV (p-value=0·001) and global WMH volume (p-value<0·001), which increased with higher Fazekas scores [Supplementary Figure 3A]. Non-significant differences were found in the genetic predisposition to WMH between groups. Nonetheless, the average polygenic score for WMH increased across the Fazekas-based groups, although it displayed the lowest value in the most severe group (Fazekas=3, N=10) [Supplementary Figure 3B].

**Table 1.** Demographic and cardiovascular characteristics of the study sample, also stratifying by WMH severity.

|  | Sample of the study<br>N=1072 | Non-pathological WMH<br>(Fazekas <2, N=980) | Pathological WMH<br>(Fazekas ≥2, N=83) | p-value |
| --- | --- | --- | --- | --- |
| Sex |  |  |  | 0.108 |
| Men | 383 (35.728%) | 344 (35.102%) | 37 (44.578%) |  |
| Women | 689 (64.272%) | 636 (64.898%) | 46 (55.422%) |  |
| Age | 59.000 [53.000;64.000] | 58.000 [53.000;63.000] | 64.000 [60.000;67.000] | <0.001 |
| Categorical Age |  |  |  | <0.001 |
| 47-55 yo | 377 (35.168%) | 365 (37.245%) | 10 (12.048%) |  |
| 56-64 yo | 469 (43.750%) | 427 (43.571%) | 36 (43.373%) |  |
| 65-77 yo | 226 (21.082%) | 188 (19.184%) | 37 (44.578%) |  |
| Years of Education | 14.000 [11.000;17.000] | 14.000 [11.000;17.000] | 14.000 [11.000;17.000] | 0.539 |
| Categorical Education |  |  |  | 0.986 |
| 5-10 yr | 225 (20.989%) | 206 (21.020%) | 18 (21.687%) |  |
| 11-14 yr | 317 (29.571%) | 290 (29.592%) | 24 (28.916%) |  |
| 15-18 yr | 530 (49.440%) | 484 (49.388%) | 41 (49.398%) |  |
| APOE ε4 carriership |  |  |  | 0.211 |
| Allele ε4 Carriers | 415 (38.713%) | 374 (38.163%) | 38 (45.783%) |  |
| Allele ε4 Non-Carriers | 657 (61.287%) | 606 (61.837%) | 45 (54.217%) |  |
| CAIDE-I score | 5.000 [4.000;7.000] | 5.000 [4.000;7.000] | 7.000 [5.000;8.000] | <0.001 |
| Hypertension |  |  |  | <0.001 |
| Hypertensive | 163 (15.205%) | 136 (13.878%) | 26 (31.325%) |  |
| Non-Hypertensive | 909 (84.795%) | 844 (86.122%) | 57 (68.675%) |  |
| Body Mass Index |  |  |  | 0.051 |
| Underweight | 3 (0.280%) | 3 (0.306%) | 0 (0.000%) |  |
| Normal Weight | 443 (41.325%) | 416 (42.449%) | 23 (27.711%) |  |
| Overweight | 471 (43.937%) | 419 (42.755%) | 47 (56.627%) |  |
| Obese | 155 (14.459%) | 142 (14.490%) | 13 (15.663%) |  |
| Hypercholesterolemia |  |  |  | 0.132 |
| Hypercholesterolemic | 270 (25.187%) | 250 (25.510%) | 28 (33.735%) |  |
| Non-Hypercholesterolemic | 802 (74.813%) | 730 (74.490%) | 55 (66.265%) |  |
| Minutes of physical activity (per month) | 2011.875<br>[1012.875;3514.375] | 1998.438 [983.250;3531.656] | 2452.500<br>[1139.375;3413.438] | 0.400 |
| Physical activity |  |  |  | 0.515 |
| Active ($\geq 150$ min/week) | 1037 (96.735%) | 946 (96.531%) | 82 (98.795%) | |
| Inactive ($< 150$ min/week) | 35 (3.265%) | 34 (3.469%) | 1 (1.205%) | |
| PRS-WMH | -0.002 [-0.005;<0.001] | -0.002 [-0.005;<0.001] | -0.001 [-0.004;0.001] | 0.168 |
| Total intracranial volume | 1441538.539<br>[1332020.471;1561860.173] | 1438698.539<br>[1329745.167;1554750.589] | 1517769.830<br>[1379781.759;1622791.098] | **0.001** |
| WMH volume | 0.001 [0.001;0.002] | 0.001 [0.001;0.002] | 0.006 [0.004;0.009] | **<0.001** |
| Fazekas scale |  |  |  | **<0.001** |
| NA | 9 (0.840%) |  |  |  |
| 0 | 343 (31.996%) | 343 (35.000%) | 0 (0.000%) |  |
| 1 | 637 (59.421%) | 637 (65.000%) | 0 (0.000%) |  |
| 2 | 73 (6.81%) | 0 (0.000%) | 73 (87.952%) |  |
| 3 | 10 (0.933%) | 0 (0.000%) | 10 (12.048%) |  |
| Time between clinical evaluation and MRI scan | 3.58 [3.11;4.33] | 3.595 [3.123;4.363] | 3.422 [3.089;3.970] | 0.058 |

*Legend: TIV (Total Intracranial Volume); MRI (Magnetic Resonance Imaging); WMH (White Matter Hyperintensities); yo (years old); yr (years). Median and interquartile range were reported for continuous non-normally distributed variables. Counts and percentage were reported for categorical variables. TIV was defined as mm<sup>3</sup>. WMH volume was adjusted for TIV and expressed for each 100 mm<sup>3</sup>. There were 9 missing values for the Fazekas scale score.*

### The polygenic risk score of WMH is a proxy of larger WMHV in individuals at low cardiovascular risk for late-life dementia, although it is not indicative of WMH severity

In the low-risk group for late-life dementia, genetic predisposition to WMH was positively associated with larger global WMHV, even after adjusting for the effect of age and sex (ρ=0·090 [0·029, 0·149], p-value=0·004) [Figure 1, Table 2]. Sensitivity analyses showed that results were still significant after adjusting the model for hypertension status (ρ=0·082 [0·021, 0·142], p-value=0·004), CAIDE-I score (ρ=0·090 [0·030, 0·150], p=0·003) and WMH severity (ρ=0·079 [0·019, 0·139], p=0·003) [Figure 1, Table 2,]. Regarding regional WMH, the PRS_WMH_ was associated with larger periventricular (ρ=0·089 [0·029, 0·148], p=0·004), deep (ρ=0·088 [0·028, 0·147], p=0·004) and juxtacortical (ρ=0·065 [0·005, 0·124], p=0·034) WMHV [Table 2, Figure 1]. Nonetheless, the correlation between genetic predisposition to WMH and the PC1 juxtacortical WMHV did not remain significant after adjusting for hypertension and WMH severity. Additionally, we explored whether PRS_WMH_ was associated with higher odds to present pathological WMH. Results from the logistic regression model did not show a significant association between higher genetic predisposition to WMH and WMH severity (p=0·471) [Table 3]. Nonetheless, the odds of displaying pathological WMH were higher with age (OR=2·028, p-value<0·001) and positive history of hypertension (OR=1·863, p-value=0·02).

**Table 2.**
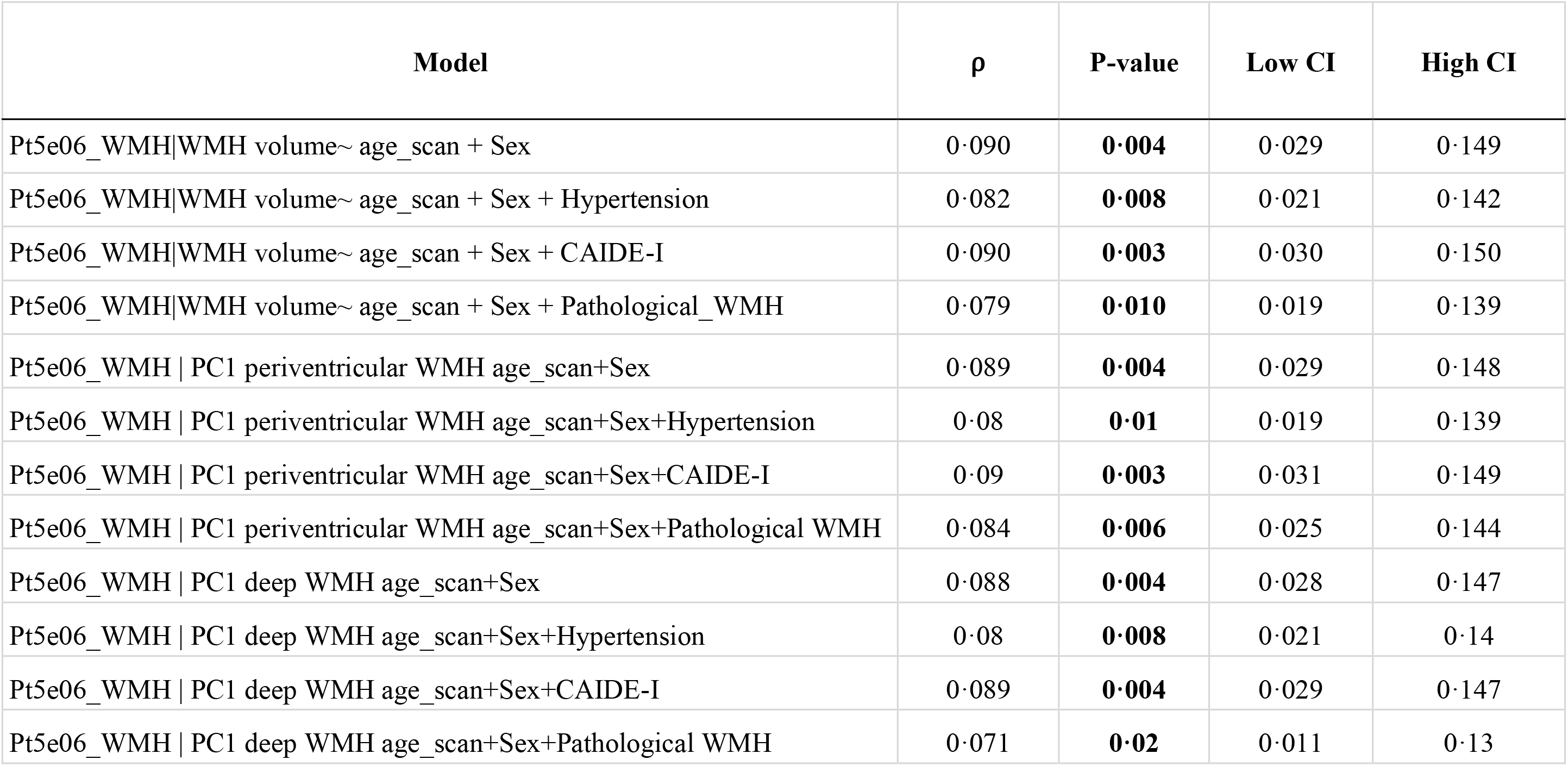
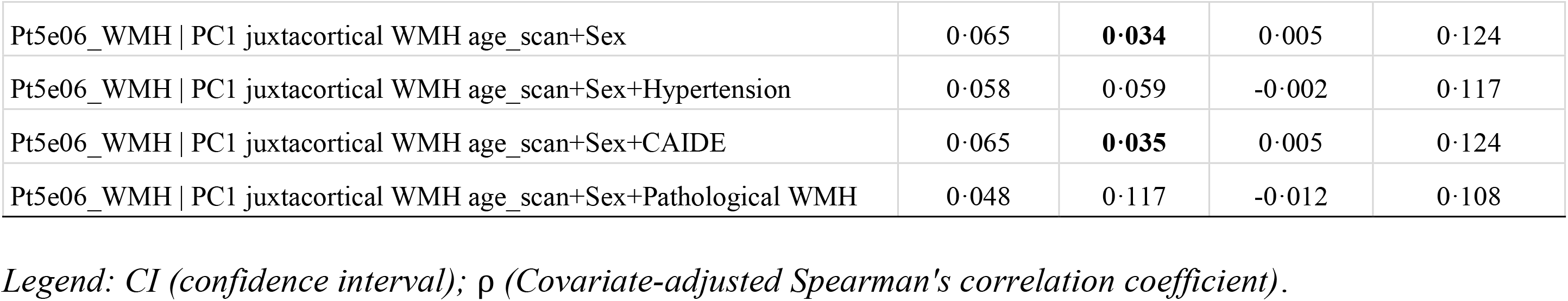
Results for the covariate-adjusted Spearman’s rank correlation test assessing the association between WMHV and PRS-WMH.

**Table 3.** Results for the logistic regression model assessing the association between WMH severity and PRS-WMH.

| Term | Beta | OR | SE | P-value | Low CI | High CI |
| --- | --- | --- | --- | --- | --- | --- |
| (Intercept) | -2.618 | 0.073 | 0.206 | **0** | -3.04 | -2.232 |
| PRS-WMH | 0.086 | 1.090 | 0.119 | 0.471 | -0.147 | 0.322 |
| Age | 0.707 | 2.028 | 0.127 | **0** | 0.463 | 0.961 |
| Sex:Women | -0.356 | 0.700 | 0.238 | 0.135 | -0.821 | 0.115 |
| Hypertension:Hypertensive | 0.622 | 1.863 | 0.267 | **0.02** | 0.085 | 1.136 |

Legend: CI (confidence interval); OR (Odds Ratio); SE (standard error). Age and PRS-WMH were standardized when included in the model. Non-pathological group was the reference group for WMH severity outcome.

**Figure 1.**
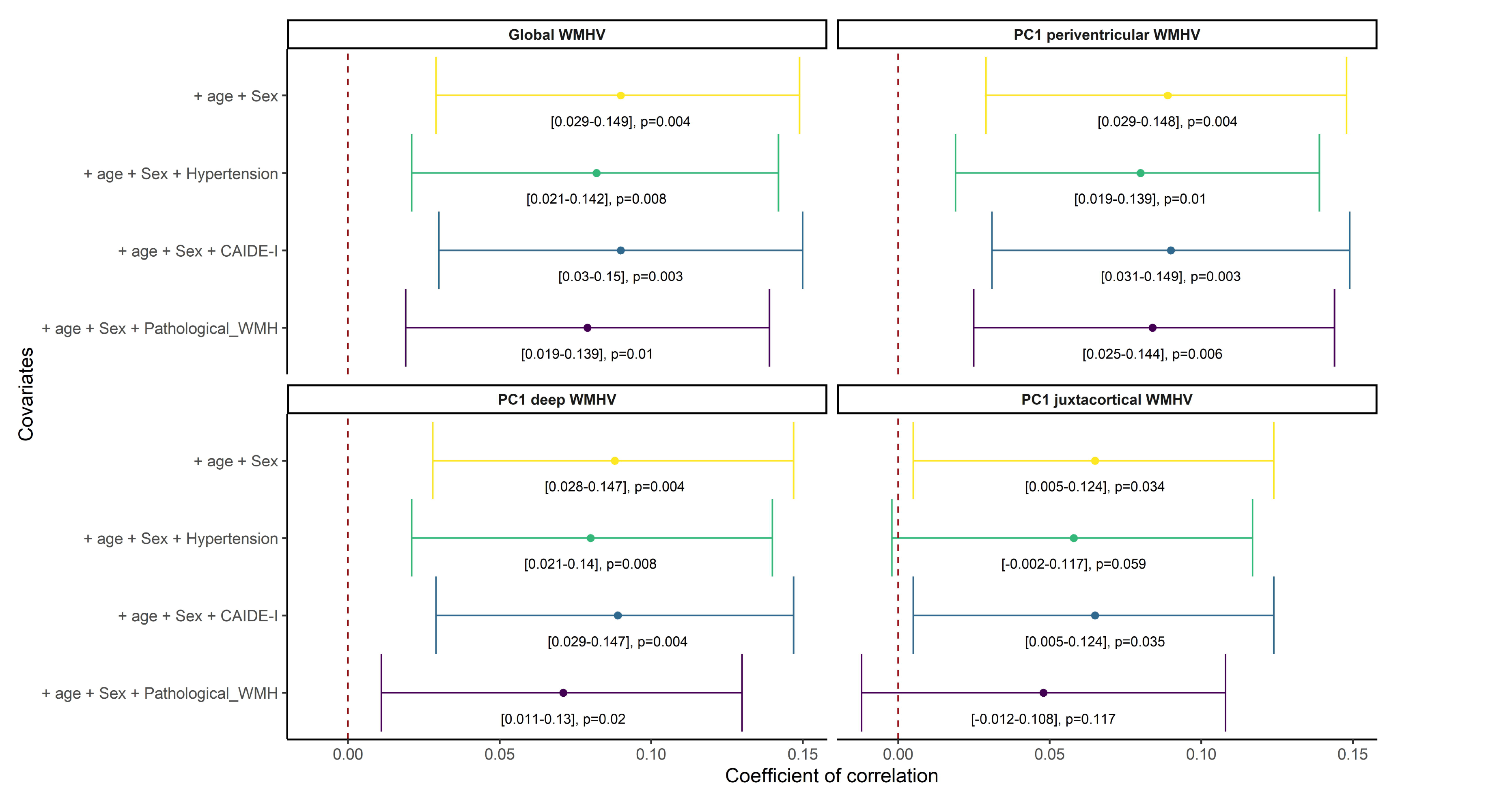
Covariate adjusted Spearman’s rank correlation test assessing the association between both global and regional WMHV and the genetic predisposition to WMH. *Legend: WMH* ( *White matter hyperintensities*), *WMHV* ( *White matter hyperintensities volumes*). *The main model* (*yellow*) *included age and sex as covariates. Sensitivity analyses additionally included hypertension status, the CAJDE-1 score and WMH severity. Confidence intervals and p-values were reported*.

### Genetic factors conferring higher risk to WMH are mainly involved in cellular and neural function highly interconnected through lipid-regulatory mechanisms

Enrichment analysis of the genes annotated to the variants within the PRS-WMH, identified 102 significantly enriched biological pathways (p-value <0·05) mainly involved in lipoprotein metabolic processes [Supplementary Table 6] [Figure 2A]. Moreover, the enrichment analysis of the genes annotated to the variants that remained after removing those highly correlated, identified 121 enriched biological pathways (p-value <0·05) mainly involved in dendritic spine organization development, transport across blood-brain barrier, vascular transport, regulation of synaptic plasticity and Rho protein signal transduction [Supplementary Table 7] [Figure 2B].

**Figure 2.**
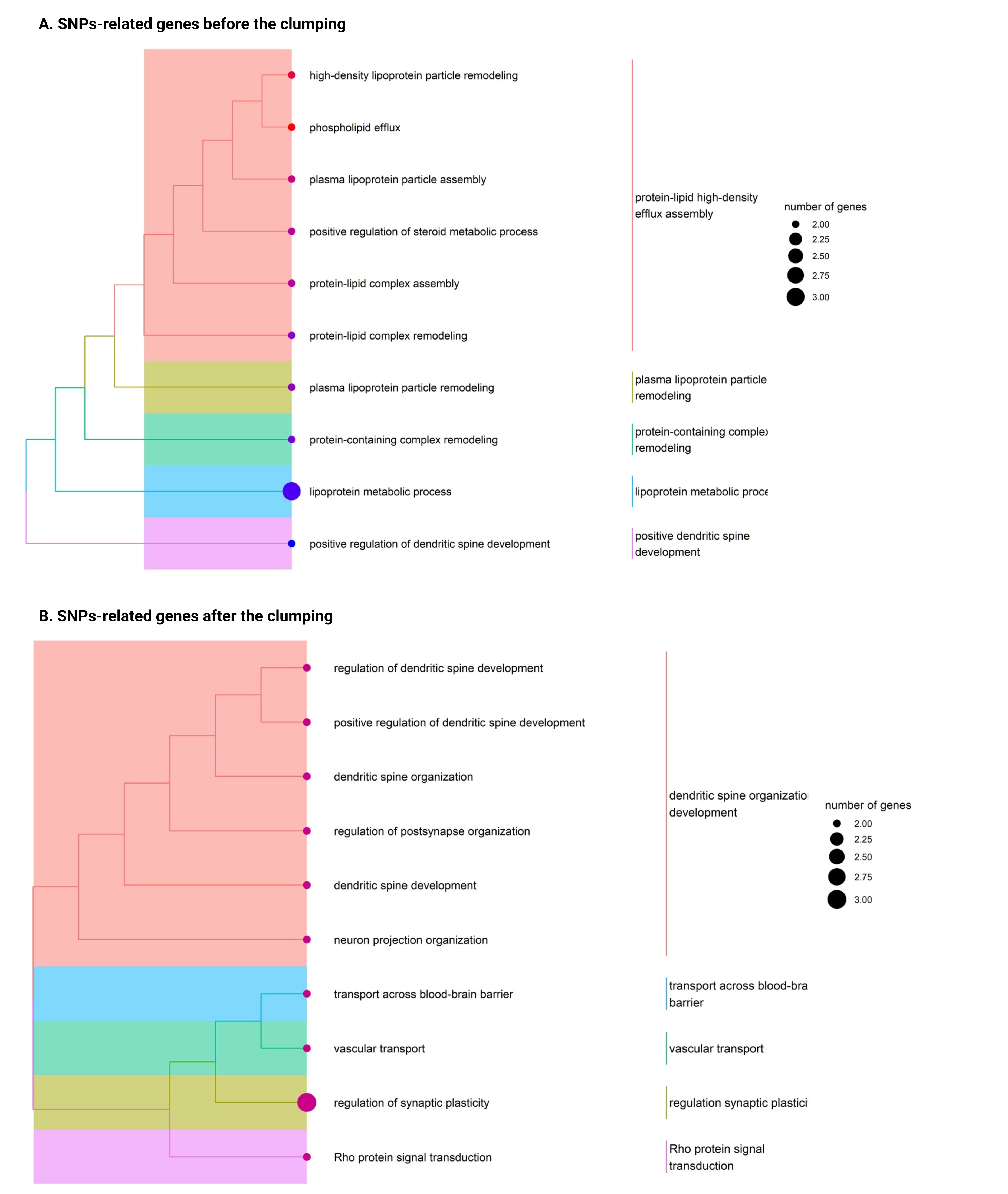
Enrichment analysis results display the main biological processes in which SNP-annotated genes are involved. *Legend: One-sided version of Fisher’s exact test was pe1formed to determine whether known biologicalfunctions were overrepresented or enriched in the gene list and calculate the probability of observing a set of genes in a particular biological pathway by chance. On the lefi* (*A*), *the enrichment analysis results show a more comprehensive interpretation of the full spectrum of genetic variants associated with WMH. On the right* (*B*), *the enrichment analysis results focused on the specific SNPs that remained after the clumping and were included in the PRS-WMH calculation. Significant results were reported at nominal p-value <0*.*05 and biological mechanisms were grouped into main functions based on their similarity*.

### The polygenic risk score of WMH is a proxy of larger WMHV in specific populations at low cardiovascular risk for late-life dementia

Genetic predisposition to WMH was positively associated with larger WMHV in non-hypertensives (ρ=0·095 [0·029, 0·161], p-value=0·005) and non-obese (ρ=0·09 [0·025, 0·154], p-value=0·007) individuals at low cardiovascular risk for late-life dementia [Table 4, Figure 3]. Moreover, genetic predisposition to WMH was positively associated with larger WMHV in women (ρ=0·081 [0·007, 0·154], p-value=0·031), hypercholesterolemic individuals (ρ=0·19 [0·072, 0·303], p-value=0·002) and participants with less than 11 years of education (ρ=0·205 [0·075, 0·328], p-value=0·002). Similarly, in participants older than 55 years, WMHV were positively correlated with a higher genetic predisposition to WMH (56-64 years-old group: ρ=0·137 [0·047, 0·225], p-value=0·003; 65-77 years-old group: ρ=0·144 [0·017, 0·267], p-value=0·027) [Table 4]. Regarding physical activity, genetic predisposition to WMH was positively associated with larger WMHV in both groups, although the correlation was stronger in the inactive group (ρ=0·207 [0·045, 0·368], p-value=0·013) than in the active one (ρ=0·072 [0·007, 0·137], p-value=0·031).

**Table 4.** Results for the covariate-adjusted Spearman’s rank correlation test assessing the association between WMHV and PRS-WMH stratifying by the CAIDE-components.

| Groups | Model | $\rho$ | P-value | Low CI | High CI |
| --- | --- | --- | --- | --- | --- |
| Women | PRS-WMH\|WMH volume ~ age_scan | 0.081 | **0.031** | 0.007 | 0.154 |
| Men | PRS-WMH\|WMH volume ~ age_scan | 0.1 | 0.061 | -0.005 | 0.202 |
| Hypertensive | PRS-WMH\|WMH volume ~ age_scan+Sex | 0.023 | 0.763 | -0.128 | 0.174 |
| Non-Hypertensive | PRS-WMH\|WMH volume ~ age_scan+Sex | 0.095 | **0.005** | 0.029 | 0.161 |
| Hypercholesterolemic | PRS-WMH\|WMH volume ~ age_scan+Sex | 0.19 | **0.002** | 0.072 | 0.303 |
| Non-Hypercholesterolemic | PRS-WMH\|WMH volume ~ age_scan+Sex | 0.052 | 0.148 | -0.018 | 0.121 |
| Obese | PRS-WMH\|WMH volume ~ age_scan+Sex | 0.064 | 0.451 | -0.102 | 0.225 |
| Non-Obese | PRS-WMH\|WMH volume ~ age_scan+Sex | 0.09 | **0.007** | 0.025 | 0.154 |
| Age 47- 55 yo | PRS-WMH\|WMH volume ~ Sex | -0.001 | 0.984 | -0.105 | 0.103 |
| Age 56-64 yo | PRS-WMH\|WMH volume ~ Sex | 0.137 | **0.003** | 0.047 | 0.225 |
| Age 65-77 yo | PRS-WMH\|WMH volume ~ Sex | 0.144 | **0.027** | 0.017 | 0.267 |
| Education 5-10 yr | PRS-WMH\|WMH volume ~ age_scan+Sex | 0.205 | **0.002** | 0.075 | 0.328 |
| Education 11-14 yr | PRS-WMH\|WMH volume ~ age_scan+Sex | 0.061 | 0.287 | -0.051 | 0.172 |
| Education 15-18 yr | PRS-WMH\|WMH volume ~ age_scan+Sex | 0.065 | 0.136 | -0.02 | 0.15 |
| Smoker | PRS-WMH\|WMH volume ~ age_scan+Sex | 0.091 | 0.137 | -0.029 | 0.208 |
| Non-Smoker | PRS-WMH\|WMH volume ~ age_scan+Sex | 0.09 | **0.004** | 0.029 | 0.149 |
| Active | PRS-WMH\|WMH volume ~ age_scan+Sex | 0.072 | **0.031** | 0.007 | 0.137 |
| Inactive | PRS-WMH\|WMH volume ~ age_scan+Sex | 0.207 | **0.013** | 0.045 | 0.358 |

Legend: CI (confidence interval); $\rho$ (Covariate-adjusted Spearman's correlation coefficient).

**Figure 3.**
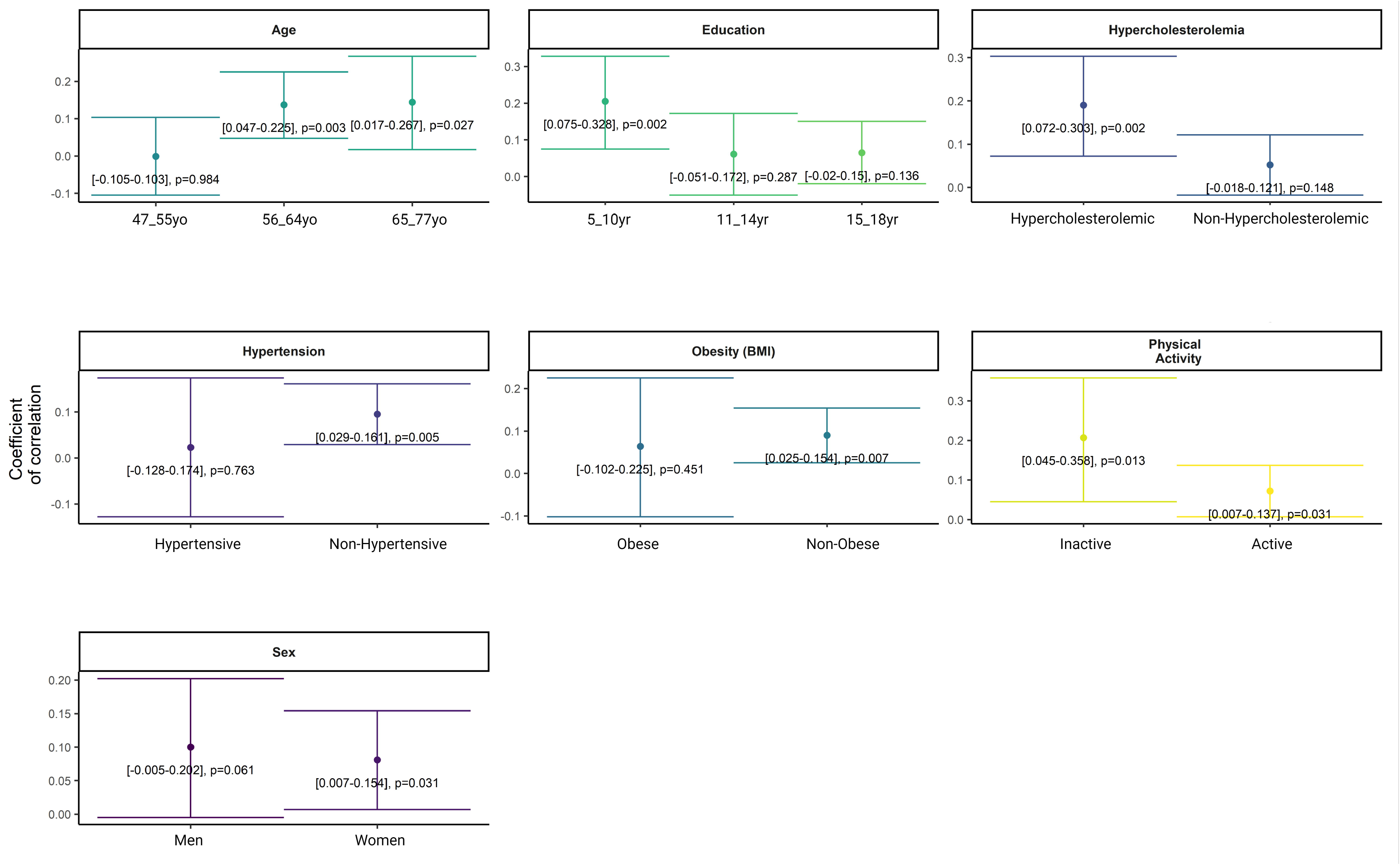
Stratified covariate-adjusted Spearman’s rank correlation test assessing the association between WMHV and PRS-WMH in individuals at low cardiovascular risk for late-life dementia. *Legend: Models were stratified by CA/DE-I components. Age and sex were included as covariates when required. Corifidence intervals and p-values were reported*.

## Discussion

In this study, we explored the biological processes associated with developing WMH in CU middle-aged individuals at low cardiovascular risk for late-life dementia. We found that in at-low risk individuals, WMH were mainly driven by lipid-related biological processes. Our results suggest that targeting cholesterol levels in middle age could be a feasible approach to prevent the development of cerebrovascular disease in individuals with a genetic predisposition to WMH, even if they show a low-risk cardiovascular profile as determined by traditional risk factors.

As a primary result, we found that higher genetic predisposition to WMH was associated with larger WMHV, even after controlling for the effect of age, sex, hypertension and the predicted 20-years risk of dementia based on the cardiovascular profile. These findings support the pertinence of studying biological pathways linked to genetic factors leading to WMH development in individuals with a low-risk profile. Remarkably, our results also showed that the PRS_WMH_ was associated with larger WMHV, independently of the spatial distribution. Uniquely for the correlation with juxtacortical WMHV, we lost the significance of the association when controlling for hypertension status and WMH severity. Recent studies found region-specific effects of genetic factors on WMHV.^17^ Some studies^29^ suggested that, beyond the specific spatial pattern of the regional WMH subtypes, juxtacortical WMHV could also indicate cerebrovascular disease progression, from periventricular to more external areas. Therefore, in line with this hypothesis, we could suggest that in our study, the PRS_WMH_ was associated with larger periventricular WMHV, even after controlling for covariates, because they represent earlier markers, less affected by modifiable risk factors. On the contrary, juxtacortical WMHV would reflect later stages in the disease progression, and WMH-related modifiable risk factors could explain a higher variability. A better understanding of these mechanisms will contribute to a better characterization of the pathophysiology of WMH. When we explored the association between the PRS_WMH_ and greater WMH severity, we did not find significant results. Only age and hypertension were significantly associated with pathological WMH. Age and hypertension are the main risk factors of WMH, and previous studies have reported the association of both risk factors with WMH severity.^30^ The lack of association between WMH severity and the genetic predisposition to WMH could be due to the sample size. The study was defined by CU middle-aged individuals with a limited number of participants with scores for the Fazekas scale above one. This limited number influenced the sample size when categorizing individuals into severity groups, and consequently reduced the statistical power of the analysis. Finally, Fazekas scores in our sample were associated with distinct ranges of WMHV that overlapped, which could explain the lack of significant association between the PRS_WMH_ and WMH severity compared to the significant results with the continuous WMHV. The overlap of WMHV ranges across the Fazekas scale has been reported in other studies, specifically in the severe groups ^31^, which suggest the preferential use of volumetric segmentation.

In a second step, we explored through which biological pathways WMH-related genetic factors were increasing the risk of displaying larger WMH. Results showed that the risk of presenting WMH was mainly driven by lipid-regulatory mechanisms, primarily focused on cellular and neural function. In this line, *APOE* likely plays a role in this process, as it is associated with WMHV and lipidic processes. On the one hand, *APOE* isoforms are differentially associated with WMHV^8,11^. CU *APOE*-ε4 homozygous have a higher risk of pathological WMH levels compared to heterozygotes^11^, while *ε2* carriers exhibit a protective effect on global WMH^8^. On the other hand, *APOE* is associated with lipid regulation, influencing plasma levels and contributing to lipid homeostatic control ^32-33^. The ε4 allele is associated with a higher risk of atherosclerosis and higher plasma levels of total and low-density lipoprotein (LDL) cholesterol^34^. The PRS_WMH_ included a low number of genetic variants in the *APOE* region with a relatively smaller effect size compared to genetic variants in other relevant *loci*. Our results suggest that, beyond *APOE*, other genes contribute to the genetic risk of displaying larger WMHV. In this line, in the GWAS of reference^15^, most genome-wide significant SNPs were located in chromosomes 2 (*e*.*g. STON1, COG2*), 16 (*e*.*g. C16orf95, SLC38A1*) and 17 (*e*.*g. DCAKD, TRIM65*). Post-gwas analyses showed associations between these *loci* and the expression levels of certain genes that were cell-specific for astrocytes and oligodendrocytes. Oligodendrocytes provide the myelin that surrounds axons and are highly enriched in cholesterol, as myelin is the major pool of cholesterol (70–80%) in the brain^35^. Astrocytes synthesize cholesterol in aging brains whose neurons’ ability to produce cholesterol is impaired. In sum, the link between cholesterol metabolism and small vessel disease is complex and spans vascular, neuronal and glial processes.

Finally, *post-hoc* analyses showed that genetic predisposition to WMH was associated with larger WMHV in the risk-groups for older age, low educational attainment and hypercholesterolemia. A recent study^36^ showed that hyperlipidemia was among the greatest modifiable risk factors affecting WMH. These results may highlight the role of abnormal lipid metabolism in the presence of WMH. Previous findings^37^ showed that cholesterol-lowering treatments (*i*.*e*. statins) in individuals with familial hypercholesterolemia (FH) result in non-differences in the occurrence of WMH between FH-heterozygous individuals and controls. These findings highlight the feasibility of implementing cholesterol-lowering therapies to modify the influence of genetic risk on the disease incidence. Regarding the observed association in individuals with low educational attainment, a study^6^ showed a direct negative association between education and WMHV in the general population. This negative association could be explained by differences in socioeconomic ^38-39^ and lifestyle factors^40-41^ between low- and high-educated individuals that contribute to the presence of WMH. Finally, the PRS-WMH was also associated with larger WMHV in individuals older than 55 years. Age is the main non-modifiable risk factor for WMH, which are usually observed in individuals above 60 years. However, they are common in healthy middle-aged individuals, with a prevalence ranging from 40-70% in the fifth decade of age^42^. On the contrary, the PRS_WMH_ was related to larger WMHV in the non-risk group for hypertension, the main modifiable risk factor of WMH. A recent multi ancestry meta-analysis of WMH-GWAS^43^ showed new *loci* significantly associated with WMHV independently of hypertension status, suggesting the relevant role of genetics explaining the presence of WMHV even in groups clinically defined as non-risk groups (*i*.*e*. normotensive individuals).

Our study highlights the suitability of the ALFA study, composed of CU individuals enriched with genetic factors for dementia, to identify at-risk individuals for cerebrovascular disease. At-risk individuals have been identified even though the study sample is healthier than expected from an age-matched cohort selected from the general population, as it does not include individuals with relevant medical pathology or neurological diseases^18^. Nonetheless, this study also presents some limitations. We can not provide a complete interpretation of the complex interplay between modifiable^44-45^ and non-modifiable risk factors contributing to WMH severity. In addition, the study has a cross-sectional design and does not evaluate the relative contribution of CVRF and genetics to the rate of progression of WMHV. Further analyses are needed to better understand their contribution to cerebrovascular disease progression. Finally, other diseases-related genetic factors may be associated with WMHV, which could reveal alternative pathways leading to larger WMH. Additional analyses are needed to explore both the genetic correlation and common biological pathways between WMH and other conditions.

## Data Availability

All data produced in the present study are available upon reasonable request to the authors.

## Declarations

### Contributors

PG: conceptualization and design of the study, data analysis, writing (original draft), interpretation of the data. BR-F: writing (review and editing). CM: resources and writing (review and editing). AB-S: data curation, and writing (review and editing). JH: data curation, and writing (review and editing). ME: data acquisition and curation. CH-S: data curation, and writing (review and editing). MC-C: writing (review and editing). CT-P: writing (review and editing). IG-L: writing (review and editing). AN: writing (review and editing). JDG: conceptualization and design of the study, supervision, writing (original draft), interpretation of the data. NV-T: conceptualization and design of the study, supervision, writing (original draft), interpretation of the data. All authors had full access to all the data in the study, reviewed and edited the manuscript, approved the final version of the manuscript, and had final responsibility for the decision to submit for publication.

### Data sharing

De-identified data supporting the findings of this study are available on request from the corresponding author (NV-T). Requests are evaluated by the Scientific Committee at Barcelonaβeta Brain Research Center and, if granted, data are shared and regulated by a Data Sharing Agreement.

## Acknowledgements

This publication is part of the ALFA study (ALzheimer and FAmilies). The authors would like to express their most sincere gratitude to the ALFA project participants and relatives without whom this research would have not been possible. Collaborators of the ALFA Study are: Müge Akinci, Federica Anastasi, Felipe Hernández-Vilamizar, Armand Gonzalez-Escalante, Annabella Beteta, Raffaele Cacciaglia, Lidia Canals, Alba Cañas, Carme Deulofeu, Maria Emilio, Irene Cumplido-Mayoral, Marta del Campo, Carme Deulofeu, Ruth Dominguez, Maria Emilio, Sherezade Fuentes, Marina García, Laura Hernández, Gema Huesa, Jordi Huguet, Laura Iglesias, Esther Jiménez, Helena Blasco, Javier Torres, David López-Martos, Paula Marne, Tania Menchón, Paula Ortiz-Romero, Marina de Diego, José Contador-Muñana, Eleni Palpatzis, Wiesje Pelkmans, Albina Polo, Sandra Pradas, Mahnaz Shekari, Lluís Solsona, Anna Soteras, Núria Tort-Colet and Marc Vilanova.

## Abbreviations

AD: Alzheimer’s disease
ALFA: ALzheimer’s and Families
APOE: Apolipoprotein E
BMI: Body mass index
CAC: Coronary artery calcification
CAIDE: Cardiovascular Risk Factors, Aging, and Incidence of Dementia
CEPT: Cholesteryl Ester Transfer Protein
CHD: Coronary heart disease
CU: Cognitively unimpaired
CVD: Cardiovascular Disease
CVRF: Cardiovascular Risk Factors
DNA: Deoxyribonucleic acid
DBP: Diastolic blood pressure
GWAS: Genome-wide association studies
LDL: Low-density lipoprotein
MRI: Magnetic resonance imaging
PRS: Polygenic risk score
SBP: Systolic blood pressure
SNP: Single nucleotide polymorphism
TIV: Total intracranial volume
WMH: White matter hyperintensities
WMHV: White matter hyperintensities volumes

