## Supplemental Material for "Lipid-regulatory mechanisms drive cerebrovascular disease in asymptomatic individuals at low risk for late-life dementia"

### Supplementary Methods

### Multiple Factor Analysis: reduction of the dimensionality

A multiple factor analysis (MFA) was performed to reduce the dimensionality of the data and to obtain a composite for each class of regional WMH. The MFA works by performing both group-specific and a global principal component analysis (PCA). We were interested in the group-specific PCA. The main goal was to obtain the first principal component (PC1) from each PCA that explained the highest variability in the data.

A component should satisfy the following properties:

1. Each component is a linear combination of the initial variables.
2. Pairs of components are uncorrelated.
3. Components are sorted in decreasing order regarding the variance they explain.

The obtention of the components works as follows:

(1) Regional WMHV for six brain regions (infratentorial, basal ganglia, frontal, parietal, occipital and temporal) are grouped in three main blocks based on the distance to the ventricles: periventricular, deep or juxtacortical and the spatial distribution. A total of 18 variables are available, 6 per group.

(2) Group-specific PCA are performed for each WMH subtype. The PC1 of each group-specific PCA is selected and corresponds to the combination of the 6 region-specific WMHV that explains the highest variability in the sample. All PCA-components are defined as linear combinations of all region-specific volumes and are uncorrelated between them. Subsequent principal components explain the rest of variability in the data.

(3) The first principal components for each subtype are defined in the models as outcomes for regional WMHV: PC1 periventricular WMHV, PC1 deep WMHV and PC1 juxtacortical WMHV.

### Principal components description

#### *PC1 periventricular WMHV*

The PC1 for periventricular WMHV explained 90% of periventricular WMHV variability in the sample. The most important variables contributing to the component were the frontal periventricular WMH along with the parietal and basal ganglia WMH. The temporal, occipital and infratentorial variables had a weaker contribution to the component.

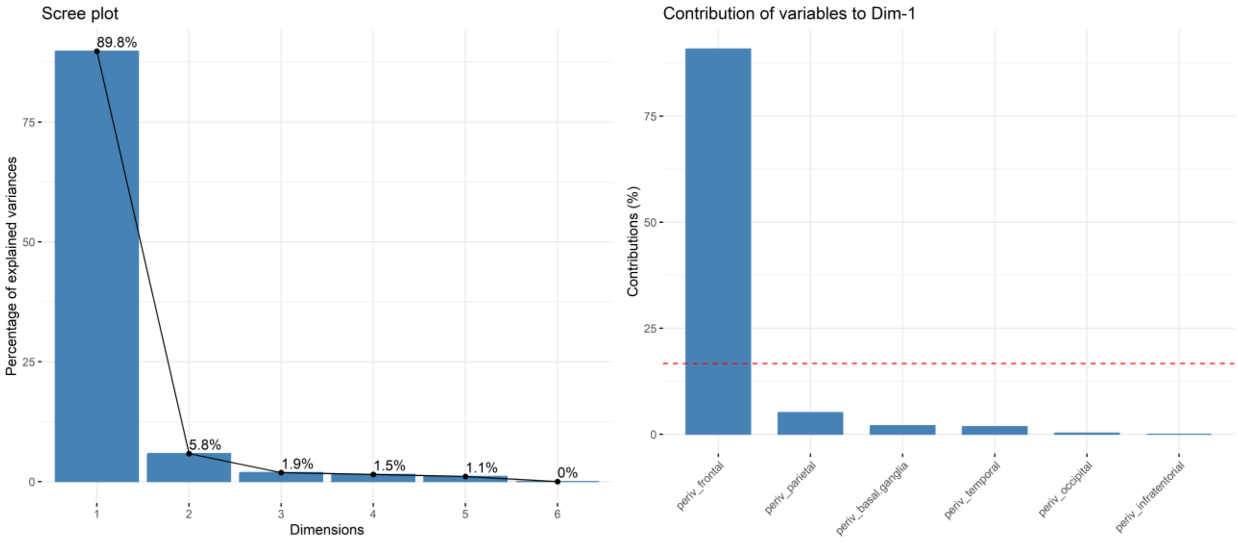

**Supplementary Figure 4.** Principal component analysis results for periventricular WMHV. On the left is described the variability explained by each independent component. On the right, the contribution of each variable to the first component.

***PC1 deep WMHV***

The PC1 for deep WMHV explained 79% of the variability for deep WMHV in the sample. The most important variables contributing to the component were the frontal deep WMH along with the parietal and temporal WMH. The occipital, basal ganglia and infratentorial variables had a weaker contribution to the component.

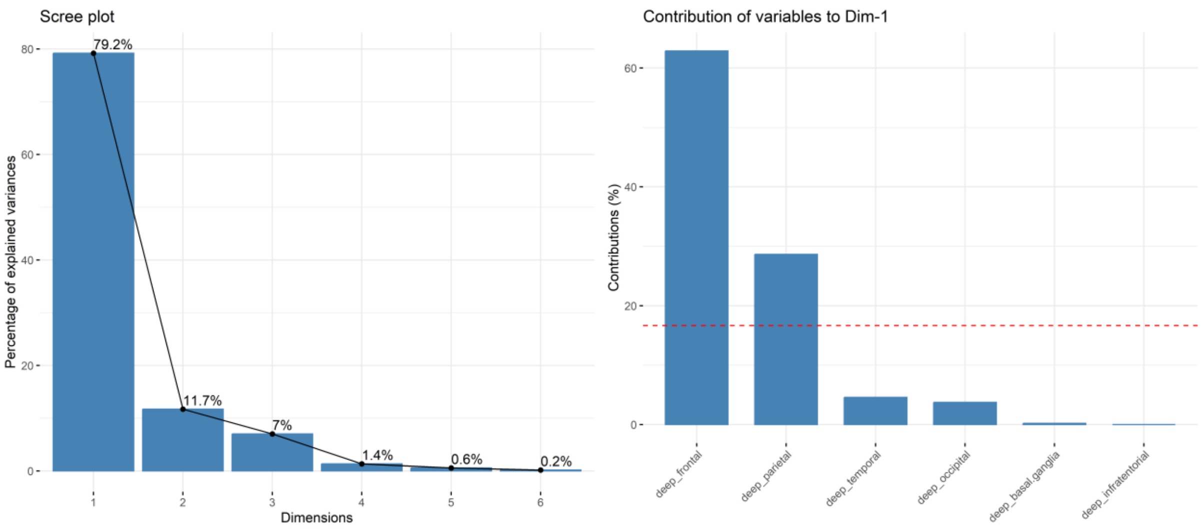

**Supplementary Figure 5.** Principal component analysis results for deep WMHV. On the left is described the variability explained by each independent component. On the right, the contribution of each variable to the first component.

***PC1 juxtacortical WMHV***

The first principal component for juxtacortical WMHV explained 88% of the variability for juxtacortical WMHV in the sample. The most important variables contributing to the component were the frontal juxtacortical WMH along with the parietal and temporal WMH. The occipital, basal ganglia and infratentorial variables had a weaker contribution to the component, same as for the deep WMHV.

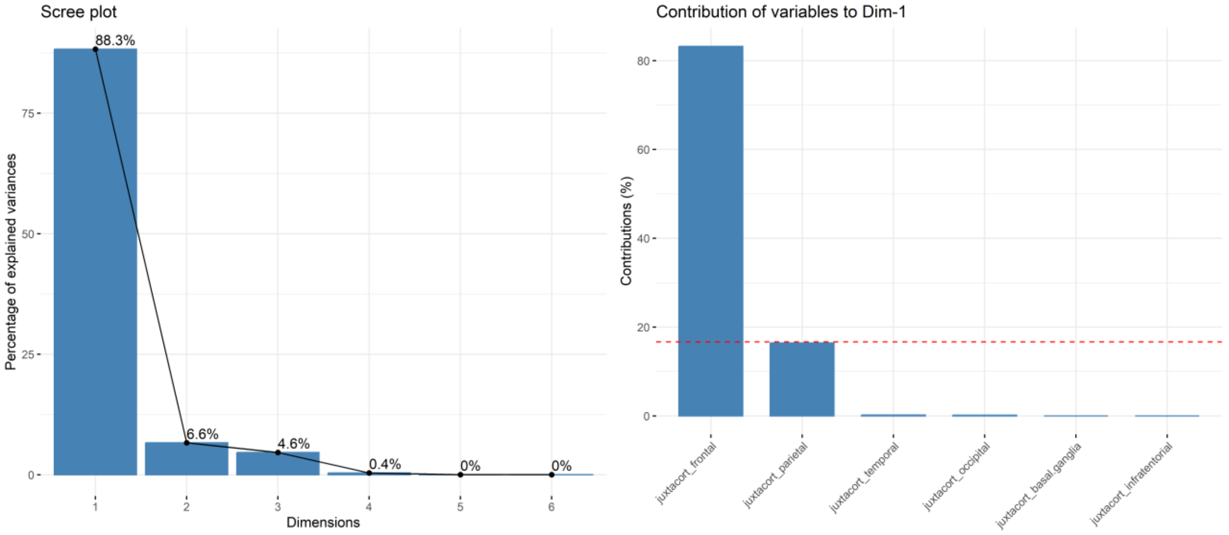

**Supplementary Figure 6.** Principal component analysis results for juxtacortical WMHV. On the left is described the variability explained by each independent component. On the right, the contribution of each variable to the first component.

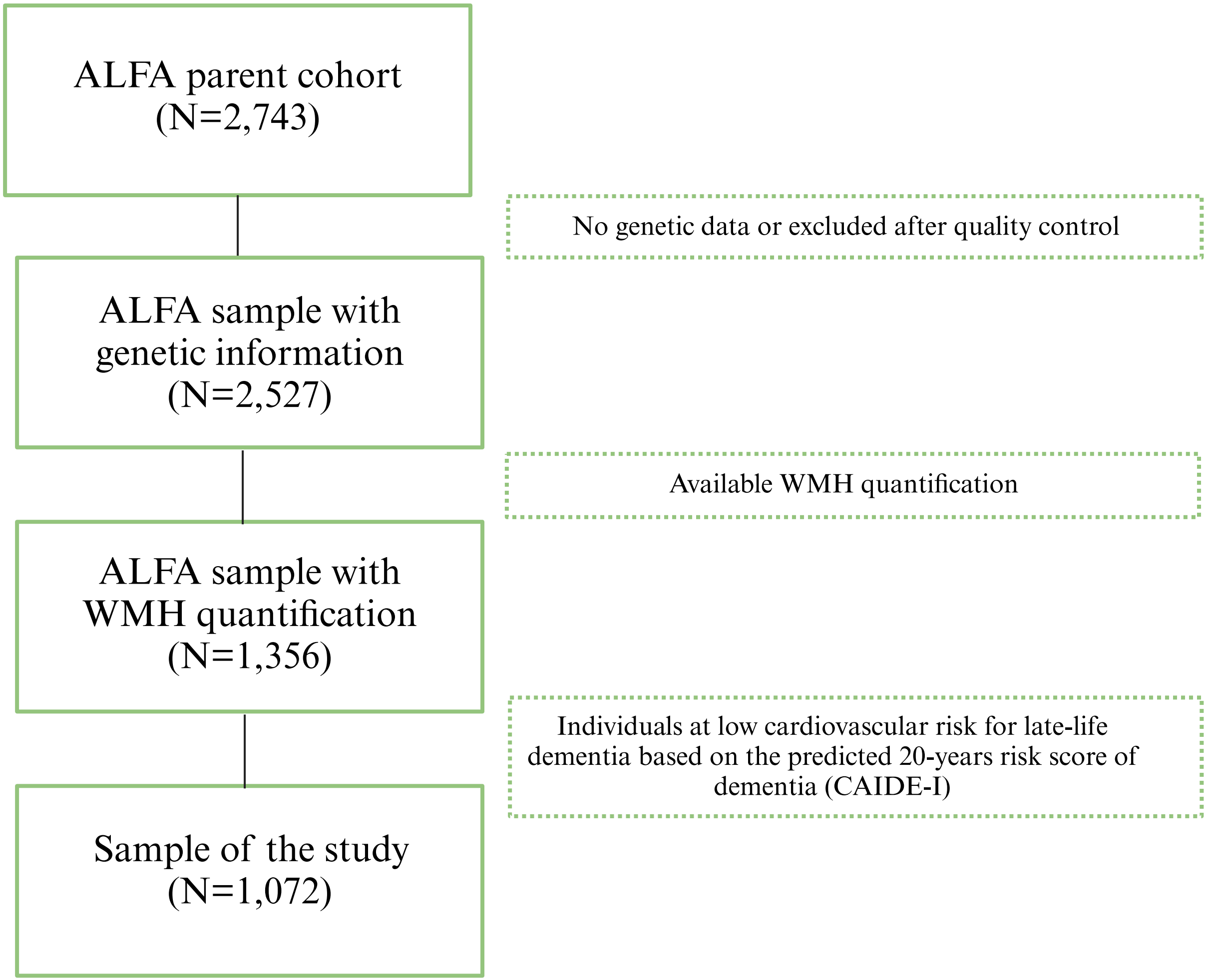

**Supplementary Figure 1.** Flowchart of the sample of the study. *Legend: ALFA (Alzheimer's and Families), WMH (White matter hyperintensities). All individuals had available quantification of WMHV, genetic data and predicted 20-years risk of dementia based on cardiovascular risk factors.*

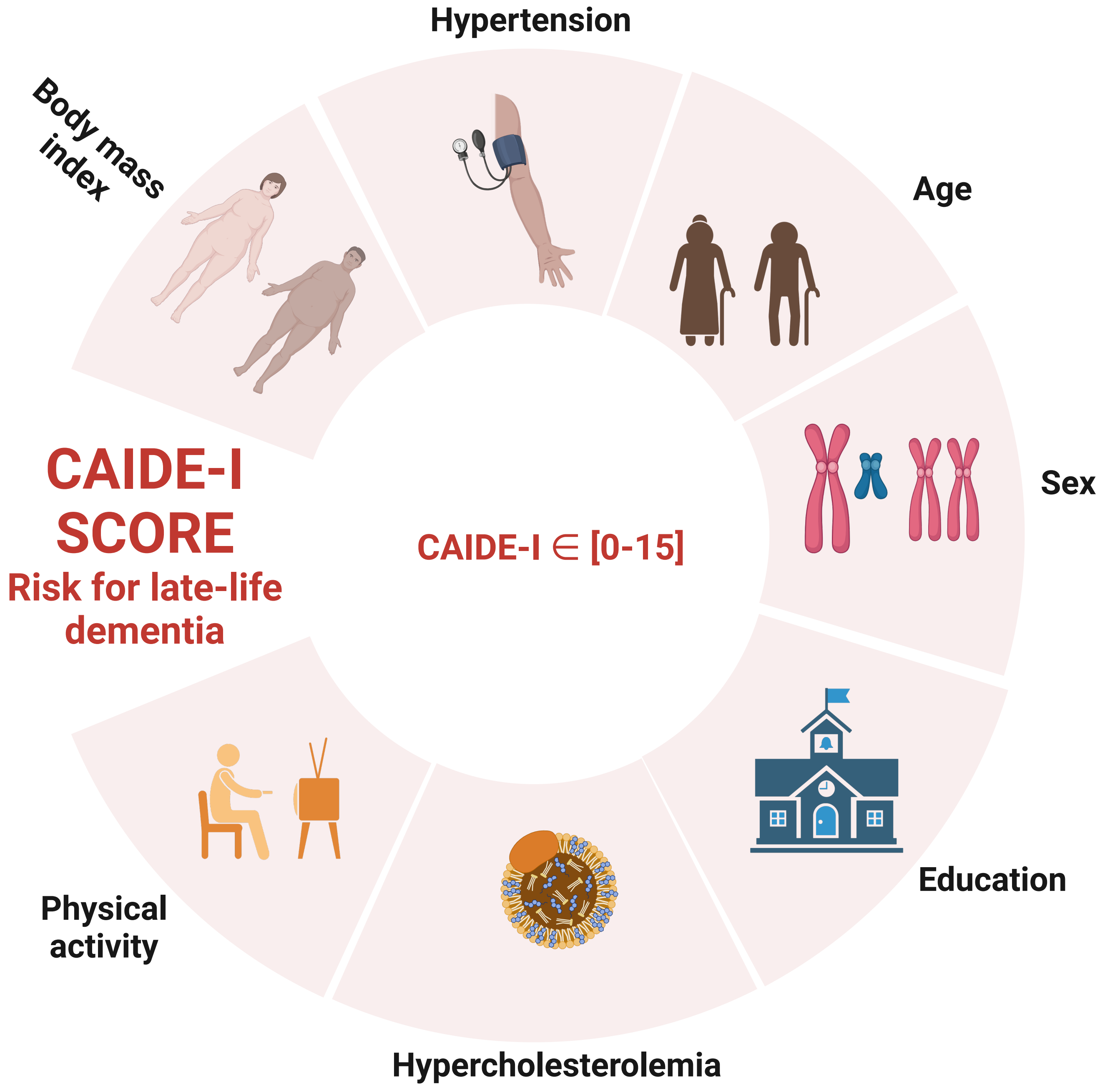

**Supplementary Figure 2.** Variables included in the predicted 20-years risk of dementia based on cardiovascular risk factors (CAIDE-I). *Legend: the probability of dementia late in life according to the CAIDE-I risk score categories are (i) CAIDE-I [0-5]: 1%; (ii) CAIDE-I [6-7] : 1.9%; (iii) CAIDE-I [8-9]: 4.2%; (iv) CAIDE-I [10-11]: 7.4% and CAIDE-I [12-15]: 16.4% (Kivipelto et al., 2006).*

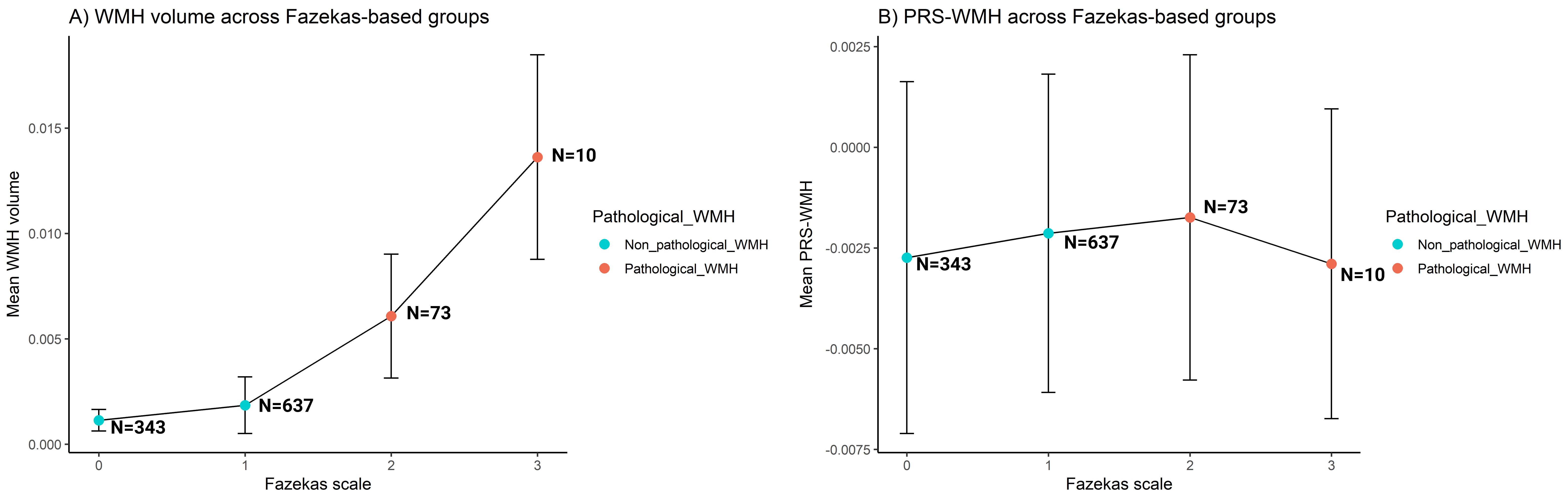

**Supplementary Figure 3.** Mean value and standard error of global WMHV along the WMH severity continuum based on the Fazekas scale. *Global WMHV were adjusted for TIV and expressed per 100mm³.*

| Risk factor | CAIDE-I risk score |
|---|---|
| **Age, years** | |
| <47 | 0 |
| 47-53 | 3 |
| >53 | 4 |
| **Education, years** | |
| ≥10 | 0 |
| 7-9 | 2 |
| 0-6 | 3 |
| **Sex** | |
| Women | 0 |
| Men | 1 |
| **Systolic blood pressure** | |
| ≤140 mmHg | 0 |
| >140 mmHg | 2 |
| **BMI, kg/m2** | |
| ≤30 | 0 |
| >30 | 2 |
| **Hypercholesterolemia** | |
| No | 0 |
| Yes | 2 |
| **Physical activity** | |
| Active | 0 |
| Inactive | 1 |
| **TOTAL** | **15** |

**Supplementary Table 1.** Cardiovascular risk factors contributing to dementia included in the CAIDE-I score and their corresponding punctuation. *Legend: higher punctuation involves higher risk for dementia. Cutoffs and punctuations were derived from Kivipelto et al., 2006.*

| **GWAS of reference** | |
|---|---|
| *Disease/Condition Classification* | Brain Endophenotypes |
| *Disease/Condition Description* | *White Matter Hyperintensities* |
| *Acronym* | *WMH* |
| *N* | 18,381 |
| *N Cases* | Continuous outcome |
| *N Controls* | Continuous outcome |
| *Genetic Ancestry* | Caucasian |
| *Genomic Annotation* | GRCh37/hg19 |
| *Base GWAS-First Author* | *Persyn et al., 2020* |
| *Link Manuscript* | https://pubmed.ncbi.nlm.nih.gov/32358547/ |
| *Access GWAS-Summary (LINK)* | http://www.kp4cd.org/dataset_downloads/stroke |
| **Summary statistic of the GWAS** | |
| *Nº of SNPs in the GWAS* | 9,712,6789 |
| *Nº of SNPs included in the PRS after the clumping* | 607,224 |
| *Nº of SNPs included in the PRS after the clumping with p-value < 5·10-6* | 25 |

**Supplementary Table 2**: Detailed information of the GWAS of reference and the summary statistics for the PRS-WMH computation. *Legend: GWAS (Genome-wide association study); SNP (single nucleotide polymorphism); N (sample size); PRS (Polygenic risk score). The optimal threshold for the p-value was determined to be* $5{\cdot}10^{-6}$ *as it included a sufficient number of SNPs to capture genetic variability in the sample and accounted for the highest variance in WMH volumes compared to the rest of scores.*

| CHR | BP | SNP | A1 | A2 | BETA | SE | P | Gene |
|---|---|---|---|---|---|---|---|---|
| 17 | 73888354 | rs34974290 | A | G | 0,110 | 0,012 | 0,000 | TRIM65 |
| 2 | 56128091 | rs7596872 | A | C | 0,122 | 0,016 | 0,000 | EFEMP1 |
| 17 | 43127708 | rs12950988 | A | G | -0,071 | 0,010 | 0,000 | DCAKD |
| 6 | 151018909 | rs6940540 | G | T | 0,059 | 0,010 | 0,000 | PLEKHG1 |
| 2 | 43103440 | rs12615761 | G | T | -0,073 | 0,013 | 0,000 | |
| 7 | 100361675 | rs2293767 | A | G | -0,059 | 0,011 | 0,000 | ZAN |
| 16 | 87237568 | rs12928520 | T | C | 0,055 | 0,010 | 0,000 | |
| 10 | 105614452 | rs11191829 | T | G | -0,054 | 0,010 | 0,000 | SH3PXD2A |
| 16 | 51442679 | rs1948948 | T | C | -0,052 | 0,010 | 0,000 | LOC102723323 |
| 16 | 87250519 | rs28655023 | G | A | -0,050 | 0,010 | 0,000 | |
| 10 | 105459116 | rs4630220 | A | G | -0,053 | 0,011 | 0,000 | SH3PXD2A |
| 2 | 43098013 | rs7594680 | C | T | -0,058 | 0,012 | 0,000 | |
| 19 | 45411941 | rs429358 | C | T | 0,066 | 0,013 | 0,000 | APOE |
| 20 | 56559939 | rs6070327 | C | T | -0,049 | 0,010 | 0,000 | |
| 6 | 97050265 | rs1855533 | C | T | -0,050 | 0,010 | 0,000 | FHL5 |
| 5 | 82857870 | rs7733216 | C | T | 0,059 | 0,012 | 0,000 | VCAN |
| 8 | 11154358 | rs7836248 | A | G | -0,064 | 0,013 | 0,000 | MTMR9 |
| 1 | 230821062 | rs16852286 | C | T | -0,091 | 0,019 | 0,000 | COG2 |
| 12 | 46595684 | rs7977873 | T | C | 0,078 | 0,017 | 0,000 | SLC38A1 |
| 10 | 25192123 | rs10764509 | A | G | 0,046 | 0,010 | 0,000 | PRTFDC1 |
| 6 | 80191532 | rs9294147 | G | A | -0,055 | 0,012 | 0,000 | LOC100506851 |
| 2 | 48759845 | rs10184783 | T | C | 0,045 | 0,010 | 0,000 | STON1 |
| 16 | 87593783 | rs117428453 | A | G | -0,161 | 0,035 | 0,000 | |
| 8 | 74452451 | rs34207938 | C | A | -0,109 | 0,024 | 0,000 | STAU2 |
| 11 | 120212601 | rs61528732 | C | T | -0,100 | 0,022 | 0,000 | ARHGEF12 |

**Supplementary Table 3.** Summary statistics of the SNPs included in the PRS-WMH calculation under the threshold $5 \cdot 10^{-6}$.

| Ensembl gene name | SNP | CHR | BP | P | Entrezgene id | Gene symbol |
|---|---|---|---|---|---|---|
| ENSG00000141569 | rs34974290 | 17 | 73888354 | 4,03E-19 | 201292 | TRIM65 |
| ENSG00000115380 | rs7596872 | 2 | 56128091 | 2,34E-14 | 2202 | EFEMP1 |
| ENSG00000172992 | rs12950988 | 17 | 43127708 | 1,18E-11 | 79877 | DCAKD |
| ENSG00000120278 | rs6940540 | 6 | 151018909 | 1,58E-09 | 57480 | PLEKHG1 |
| | rs12615761 | 2 | 43103440 | 7,32E-09 | | |
| ENSG00000146839 | rs2293767 | 7 | 100361675 | 2,52E-08 | 7455 | ZAN |
| ENSG00000260456 | rs12928520 | 16 | 87237568 | 2,60E-08 | 100506581 | C16orf95 |
| ENSG00000107957 | rs11191829 | 10 | 105614452 | 6,49E-08 | 9644 | SH3PXD2A |
| | rs1948948 | 16 | 51442679 | 6,94E-08 | | |
| ENSG00000260456 | rs28655023 | 16 | 87250519 | 4,11E-07 | 100506581 | C16orf95 |
| ENSG00000261095 | rs28655023 | 16 | 87250519 | 4,11E-07 | 101928708 | |
| ENSG00000107957 | rs4630220 | 10 | 105459116 | 7,63E-07 | 9644 | SH3PXD2A |
| | rs7594680 | 2 | 43098013 | 8,00E-07 | | |
| ENSG00000130203 | rs429358 | 19 | 45411941 | 8,04E-07 | 348 | APOE |
| | rs6070327 | 20 | 56559939 | 8,26E-07 | | |
| ENSG00000112214 | rs1855533 | 6 | 97050265 | 9,88E-07 | 9457 | FHL5 |
| ENSG00000038427 | rs7733216 | 5 | 82857870 | 1,21E-06 | 1462 | VCAN |
| ENSG00000249835 | rs7733216 | 5 | 82857870 | 1,21E-06 | | VCAN-AS1 |
| ENSG00000104643 | rs7836248 | 8 | 11154358 | 1,37E-06 | 66036 | MTMR9 |
| ENSG00000135775 | rs16852286 | 1 | 230821062 | 1,61E-06 | 22796 | COG2 |
| ENSG00000111371 | rs7977873 | 12 | 46595684 | 2,47E-06 | 81539 | SLC38A1 |
| ENSG00000099256 | rs10764509 | 10 | 25192123 | 3,16E-06 | 56952 | PRTFDC1 |
| | rs9294147 | 6 | 80191532 | 3,22E-06 | | |
| ENSG00000068781 | rs10184783 | 2 | 48759845 | 3,50E-06 | 286749 | STON1-GTF2A1L |
| ENSG00000068781 | rs10184783 | 2 | 48759845 | 3,50E-06 | 11036 | STON1-GTF2A1L |
| ENSG00000243244 | rs10184783 | 2 | 48759845 | 3,50E-06 | 11037 | STON1 |

| | | | | | | |
|---|---|---|---|---|---|---|
| | rs117428453 | 16 | 87593783 | 4,03E-06 | | |
| ENSG00000040341 | rs34207938 | 8 | 74452451 | 4,23E-06 | 27067 | STAU2 |
| ENSG00000196914 | rs61528732 | 11 | 120212601 | 4,49E-06 | 23365 | ARHGEF12 |

**Supplementary Table 4.** Annotation of the genetic variants included in the PRS-WMH to their nearest genes. *Legend: we used the biomaRt R package for annotation.*

| ID | Description | GeneRatio | BgRatio | p-value | FDR | q-value | geneID | Count |
|---|---|---|---|---|---|---|---|---|
| GO:0033700 | phospholipid efflux | 2/38 | 11/11590 | 0,001 | 0,374 | 0,353 | APOE/APOC1 | 2 |
| GO:0034375 | high-density lipoprotein particle remodeling | 2/38 | 16/11590 | 0,001 | 0,374 | 0,353 | APOE/APOC1 | 2 |
| GO:0034377 | plasma lipoprotein particle assembly | 2/38 | 22/11590 | 0,002 | 0,374 | 0,353 | APOE/APOC1 | 2 |
| GO:0045940 | positive regulation of steroid metabolic process | 2/38 | 24/11590 | 0,003 | 0,374 | 0,353 | APOE/APOC1 | 2 |
| GO:0065005 | protein-lipid complex assembly | 2/38 | 24/11590 | 0,003 | 0,374 | 0,353 | APOE/APOC1 | 2 |
| GO:0034368 | protein-lipid complex remodeling | 2/38 | 27/11590 | 0,003 | 0,374 | 0,353 | APOE/APOC1 | 2 |
| GO:0034369 | plasma lipoprotein particle remodeling | 2/38 | 27/11590 | 0,003 | 0,374 | 0,353 | APOE/APOC1 | 2 |
| GO:0034367 | protein-containing complex remodeling | 2/38 | 28/11590 | 0,004 | 0,374 | 0,353 | APOE/APOC1 | 2 |
| GO:0042157 | lipoprotein metabolic process | 3/38 | 99/11590 | 0,004 | 0,374 | 0,353 | APOE/APOC1/ NMT1 | 3 |
| GO:0060999 | positive regulation of dendritic spine development | 2/38 | 30/11590 | 0,004 | 0,374 | 0,353 | STAU2/APOE | 2 |
| GO:0033146 | regulation of intracellular estrogen receptor signaling pathway | 2/38 | 31/11590 | 0,005 | 0,374 | 0,353 | UFL1/WBP2 | 2 |
| GO:1903725 | regulation of phospholipid metabolic process | 2/38 | 31/11590 | 0,005 | 0,374 | 0,353 | MTMR9/APO C1 | 2 |
| GO:0034381 | plasma lipoprotein particle clearance | 2/38 | 34/11590 | 0,005 | 0,414 | 0,391 | APOE/APOC1 | 2 |
| GO:0071827 | plasma lipoprotein particle organization | 2/38 | 39/11590 | 0,007 | 0,439 | 0,414 | APOE/APOC1 | 2 |
| GO:0071825 | protein-lipid complex subunit organization | 2/38 | 41/11590 | 0,008 | 0,439 | 0,414 | APOE/APOC1 | 2 |
| GO:0033344 | cholesterol efflux | 2/38 | 44/11590 | 0,009 | 0,439 | 0,414 | APOE/APOC1 | 2 |
| GO:0060998 | regulation of dendritic spine development | 2/38 | 44/11590 | 0,009 | 0,439 | 0,414 | STAU2/APOE | 2 |
| GO:0030520 | intracellular estrogen receptor signaling pathway | 2/38 | 45/11590 | 0,009 | 0,439 | 0,414 | UFL1/WBP2 | 2 |
| GO:0032371 | regulation of sterol transport | 2/38 | 45/11590 | 0,009 | 0,439 | 0,414 | APOE/APOC1 | 2 |
| GO:0032374 | regulation of cholesterol transport | 2/38 | 45/11590 | 0,009 | 0,439 | 0,414 | APOE/APOC1 | 2 |
| GO:0051055 | negative regulation of lipid biosynthetic process | 2/38 | 46/11590 | 0,010 | 0,439 | 0,414 | APOE/APOC1 | 2 |
| GO:0048167 | regulation of synaptic plasticity | 3/38 | 158/11590 | 0,015 | 0,439 | 0,414 | STAU2/SLC38 A1/APOE | 3 |
| GO:0030100 | regulation of endocytosis | 3/38 | 164/11590 | 0,016 | 0,439 | 0,414 | APOE/APOC1/ STON1 | 3 |

| | | | | | | | | |
|---|---|---|---|---|---|---|---|---|
| GO:0097006 | regulation of plasma lipoprotein particle levels | 2/38 | 60/11590 | 0,016 | 0,439 | 0,414 | APOE/APOC1 | 2 |
| GO:0032787 | monocarboxylic acid metabolic process | 5/38 | 472/11590 | 0,018 | 0,439 | 0,414 | GALK1/SLC38A1/APOC1/ACOX1/ACBD4 | 5 |
| GO:0030258 | lipid modification | 3/38 | 173/11590 | 0,019 | 0,439 | 0,414 | MTMR9/APOC1/ACOX1 | 3 |
| GO:0097061 | dendritic spine organization | 2/38 | 66/11590 | 0,020 | 0,439 | 0,414 | STAU2/APOE | 2 |
| GO:0033143 | regulation of intracellular steroid hormone receptor signaling pathway | 2/38 | 67/11590 | 0,020 | 0,439 | 0,414 | UFL1/WBP2 | 2 |
| GO:0099175 | regulation of postsynapse organization | 2/38 | 68/11590 | 0,021 | 0,439 | 0,414 | STAU2/APOE | 2 |
| GO:0042158 | lipoprotein biosynthetic process | 2/38 | 70/11590 | 0,022 | 0,439 | 0,414 | APOE/NMT1 | 2 |
| GO:0006641 | triglyceride metabolic process | 2/38 | 72/11590 | 0,023 | 0,439 | 0,414 | APOE/APOC1 | 2 |
| GO:0015914 | phospholipid transport | 2/38 | 73/11590 | 0,024 | 0,439 | 0,414 | APOE/APOC1 | 2 |
| GO:0060996 | dendritic spine development | 2/38 | 76/11590 | 0,026 | 0,439 | 0,414 | STAU2/APOE | 2 |
| GO:0106027 | neuron projection organization | 2/38 | 76/11590 | 0,026 | 0,439 | 0,414 | STAU2/APOE | 2 |
| GO:0001649 | osteoblast differentiation | 3/38 | 195/11590 | 0,026 | 0,439 | 0,414 | UFL1/VCAN/H3-3A | 3 |
| GO:0010232 | vascular transport | 2/38 | 77/11590 | 0,026 | 0,439 | 0,414 | SLC38A1/APOE | 2 |
| GO:0150104 | transport across blood-brain barrier | 2/38 | 77/11590 | 0,026 | 0,439 | 0,414 | SLC38A1/APOE | 2 |
| GO:0030301 | cholesterol transport | 2/38 | 78/11590 | 0,027 | 0,439 | 0,414 | APOE/APOC1 | 2 |
| GO:0045833 | negative regulation of lipid metabolic process | 2/38 | 79/11590 | 0,028 | 0,439 | 0,414 | APOE/APOC1 | 2 |
| GO:0019218 | regulation of steroid metabolic process | 2/38 | 81/11590 | 0,029 | 0,439 | 0,414 | APOE/APOC1 | 2 |
| GO:2001251 | negative regulation of chromosome organization | 2/38 | 81/11590 | 0,029 | 0,439 | 0,414 | STN1/H3-3A | 2 |
| GO:0062014 | negative regulation of small molecule metabolic process | 2/38 | 84/11590 | 0,031 | 0,439 | 0,414 | APOE/APOC1 | 2 |
| GO:0006707 | cholesterol catabolic process | 1/38 | 10/11590 | 0,032 | 0,439 | 0,414 | APOE | 1 |
| GO:0008298 | intracellular mRNA localization | 1/38 | 10/11590 | 0,032 | 0,439 | 0,414 | STAU2 | 1 |
| GO:0015937 | coenzyme A biosynthetic process | 1/38 | 10/11590 | 0,032 | 0,439 | 0,414 | DCAKD | 1 |
| GO:0016127 | sterol catabolic process | 1/38 | 10/11590 | 0,032 | 0,439 | 0,414 | APOE | 1 |

| | | | | | | | | |
|---|---|---|---|---|---|---|---|---|
| GO:0033148 | positive regulation of intracellular estrogen receptor signaling pathway | 1/38 | 10/11590 | 0,032 | 0,439 | 0,414 | WBP2 | 1 |
| GO:0034370 | triglyceride-rich lipoprotein particle remodeling | 1/38 | 10/11590 | 0,032 | 0,439 | 0,414 | APOE | 1 |
| GO:0034372 | very-low-density lipoprotein particle remodeling | 1/38 | 10/11590 | 0,032 | 0,439 | 0,414 | APOE | 1 |
| GO:0034384 | high-density lipoprotein particle clearance | 1/38 | 10/11590 | 0,032 | 0,439 | 0,414 | APOE | 1 |
| GO:0060623 | regulation of chromosome condensation | 1/38 | 10/11590 | 0,032 | 0,439 | 0,414 | H3-3A | 1 |
| GO:1901030 | positive regulation of mitochondrial outer membrane permeabilization involved in apoptotic signaling pathway | 1/38 | 10/11590 | 0,032 | 0,439 | 0,414 | NMT1 | 1 |
| GO:1903365 | regulation of fear response | 1/38 | 10/11590 | 0,032 | 0,439 | 0,414 | APOE | 1 |
| GO:1903961 | positive regulation of anion transmembrane transport | 1/38 | 10/11590 | 0,032 | 0,439 | 0,414 | SLC38A1 | 1 |
| GO:0015918 | sterol transport | 2/38 | 90/11590 | 0,035 | 0,439 | 0,414 | APOE/APOC1 | 2 |
| GO:0033145 | positive regulation of intracellular steroid hormone receptor signaling pathway | 1/38 | 11/11590 | 0,035 | 0,439 | 0,414 | WBP2 | 1 |
| GO:0042159 | lipoprotein catabolic process | 1/38 | 11/11590 | 0,035 | 0,439 | 0,414 | APOE | 1 |
| GO:0045815 | epigenetic maintenance of chromatin in transcription-competent conformation | 1/38 | 11/11590 | 0,035 | 0,439 | 0,414 | WBP2 | 1 |
| GO:0090205 | positive regulation of cholesterol metabolic process | 1/38 | 11/11590 | 0,035 | 0,439 | 0,414 | APOE | 1 |
| GO:1900272 | negative regulation of long-term synaptic potentiation | 1/38 | 11/11590 | 0,035 | 0,439 | 0,414 | APOE | 1 |
| GO:1902065 | response to L-glutamate | 1/38 | 11/11590 | 0,035 | 0,439 | 0,414 | UFL1 | 1 |
| GO:1902950 | regulation of dendritic spine maintenance | 1/38 | 11/11590 | 0,035 | 0,439 | 0,414 | APOE | 1 |
| GO:0006639 | acylglycerol metabolic process | 2/38 | 93/11590 | 0,037 | 0,439 | 0,414 | APOE/APOC1 | 2 |
| GO:0006638 | neutral lipid metabolic process | 2/38 | 94/11590 | 0,038 | 0,439 | 0,414 | APOE/APOC1 | 2 |
| GO:0071901 | negative regulation of protein serine/threonine kinase activity | 2/38 | 95/11590 | 0,039 | 0,439 | 0,414 | APOE/HEXIM 1 | 2 |
| GO:0009886 | post-embryonic animal morphogenesis | 1/38 | 12/11590 | 0,039 | 0,439 | 0,414 | EFEMP1 | 1 |
| GO:0032328 | alanine transport | 1/38 | 12/11590 | 0,039 | 0,439 | 0,414 | SLC38A1 | 1 |

| | | | | | | | | |
|---|---|---|---|---|---|---|---|---|
| GO:0032488 | Cdc42 protein signal transduction | 1/38 | 12/11590 | 0,039 | 0,439 | 0,414 | APOE | 1 |
| GO:0033540 | fatty acid beta-oxidation using acyl-CoA oxidase | 1/38 | 12/11590 | 0,039 | 0,439 | 0,414 | ACOX1 | 1 |
| GO:0034380 | high-density lipoprotein particle assembly | 1/38 | 12/11590 | 0,039 | 0,439 | 0,414 | APOE | 1 |
| GO:0034638 | phosphatidylcholine catabolic process | 1/38 | 12/11590 | 0,039 | 0,439 | 0,414 | APOC1 | 1 |
| GO:0050665 | hydrogen peroxide biosynthetic process | 1/38 | 12/11590 | 0,039 | 0,439 | 0,414 | ACOX1 | 1 |
| GO:0051044 | positive regulation of membrane protein ectodomain proteolysis | 1/38 | 12/11590 | 0,039 | 0,439 | 0,414 | APOE | 1 |
| GO:0071391 | cellular response to estrogen stimulus | 1/38 | 12/11590 | 0,039 | 0,439 | 0,414 | WBP2 | 1 |
| GO:1900102 | negative regulation of endoplasmic reticulum unfolded protein response | 1/38 | 12/11590 | 0,039 | 0,439 | 0,414 | UFL1 | 1 |
| GO:1900452 | regulation of long-term synaptic depression | 1/38 | 12/11590 | 0,039 | 0,439 | 0,414 | STAU2 | 1 |
| GO:1905907 | negative regulation of amyloid fibril formation | 1/38 | 12/11590 | 0,039 | 0,439 | 0,414 | APOE | 1 |
| GO:2001140 | positive regulation of phospholipid transport | 1/38 | 12/11590 | 0,039 | 0,439 | 0,414 | APOE | 1 |
| GO:0016056 | rhodopsin mediated signaling pathway | 1/38 | 13/11590 | 0,042 | 0,439 | 0,414 | NMT1 | 1 |
| GO:0035641 | locomotory exploration behavior | 1/38 | 13/11590 | 0,042 | 0,439 | 0,414 | APOE | 1 |
| GO:0055089 | fatty acid homeostasis | 1/38 | 13/11590 | 0,042 | 0,439 | 0,414 | APOE | 1 |
| GO:0060192 | negative regulation of lipase activity | 1/38 | 13/11590 | 0,042 | 0,439 | 0,414 | APOC1 | 1 |
| GO:0098935 | dendritic transport | 1/38 | 13/11590 | 0,042 | 0,439 | 0,414 | STAU2 | 1 |
| GO:1902931 | negative regulation of alcohol biosynthetic process | 1/38 | 13/11590 | 0,042 | 0,439 | 0,414 | APOE | 1 |
| GO:1903894 | regulation of IRE1-mediated unfolded protein response | 1/38 | 13/11590 | 0,042 | 0,439 | 0,414 | UFL1 | 1 |
| GO:2001138 | regulation of phospholipid transport | 1/38 | 13/11590 | 0,042 | 0,439 | 0,414 | APOE | 1 |
| GO:0015748 | organophosphate ester transport | 2/38 | 101/11590 | 0,043 | 0,439 | 0,414 | APOE/APOC1 | 2 |
| GO:0030518 | intracellular steroid hormone receptor signaling pathway | 2/38 | 103/11590 | 0,045 | 0,439 | 0,414 | UFL1/WBP2 | 2 |
| GO:0032897 | negative regulation of viral transcription | 1/38 | 14/11590 | 0,045 | 0,439 | 0,414 | HEXIM1 | 1 |
| GO:0044794 | positive regulation by host of viral process | 1/38 | 14/11590 | 0,045 | 0,439 | 0,414 | APOE | 1 |
| GO:1905906 | regulation of amyloid fibril formation | 1/38 | 14/11590 | 0,045 | 0,439 | 0,414 | APOE | 1 |

| | | | | | | | | |
|---|---|---|---|---|---|---|---|---|
| GO:0016042 | lipid catabolic process | 3/38 | 246/11590 | 0,046 | 0,439 | 0,414 | APOE/APOC1/ ACOX1 | 3 |
| GO:0010958 | regulation of amino acid import across plasma membrane | 1/38 | 15/11590 | 0,048 | 0,439 | 0,414 | SLC38A1 | 1 |
| GO:0015936 | coenzyme A metabolic process | 1/38 | 15/11590 | 0,048 | 0,439 | 0,414 | DCAKD | 1 |
| GO:0032211 | negative regulation of telomere maintenance via telomerase | 1/38 | 15/11590 | 0,048 | 0,439 | 0,414 | STN1 | 1 |
| GO:0034244 | negative regulation of transcription elongation by RNA polymerase II | 1/38 | 15/11590 | 0,048 | 0,439 | 0,414 | HEXIM1 | 1 |
| GO:0034374 | low-density lipoprotein particle remodeling | 1/38 | 15/11590 | 0,048 | 0,439 | 0,414 | APOE | 1 |
| GO:0043691 | reverse cholesterol transport | 1/38 | 15/11590 | 0,048 | 0,439 | 0,414 | APOE | 1 |
| GO:0051004 | regulation of lipoprotein lipase activity | 1/38 | 15/11590 | 0,048 | 0,439 | 0,414 | APOC1 | 1 |
| GO:0061003 | positive regulation of dendritic spine morphogenesis | 1/38 | 15/11590 | 0,048 | 0,439 | 0,414 | STAU2 | 1 |
| GO:0061709 | reticulophagy | 1/38 | 15/11590 | 0,048 | 0,439 | 0,414 | UFL1 | 1 |
| GO:1903789 | regulation of amino acid transmembrane transport | 1/38 | 15/11590 | 0,048 | 0,439 | 0,414 | SLC38A1 | 1 |
| GO:0032368 | regulation of lipid transport | 2/38 | 110/11590 | 0,050 | 0,439 | 0,414 | APOE/APOC1 | 2 |
| GO:0045717 | negative regulation of fatty acid biosynthetic process | 1/38 | 16/11590 | 0,051 | 0,439 | 0,414 | APOC1 | 1 |
| GO:0048569 | post-embryonic animal organ development | 1/38 | 16/11590 | 0,051 | 0,439 | 0,414 | EFEMP1 | 1 |
| GO:1900221 | regulation of amyloid-beta clearance | 1/38 | 16/11590 | 0,051 | 0,439 | 0,414 | APOE | 1 |
| GO:0010544 | negative regulation of platelet activation | 1/38 | 17/11590 | 0,054 | 0,439 | 0,414 | APOE | 1 |
| GO:0010984 | regulation of lipoprotein particle clearance | 1/38 | 17/11590 | 0,054 | 0,439 | 0,414 | APOC1 | 1 |
| GO:0043117 | positive regulation of vascular permeability | 1/38 | 17/11590 | 0,054 | 0,439 | 0,414 | APOE | 1 |
| GO:0061620 | glycolytic process through glucose-6-phosphate | 1/38 | 17/11590 | 0,054 | 0,439 | 0,414 | GALK1 | 1 |
| GO:1902430 | negative regulation of amyloid-beta formation | 1/38 | 17/11590 | 0,054 | 0,439 | 0,414 | APOE | 1 |
| GO:0007266 | Rho protein signal transduction | 2/38 | 115/11590 | 0,054 | 0,439 | 0,414 | APOE/ARHGE F12 | 2 |
| GO:0008203 | cholesterol metabolic process | 2/38 | 118/11590 | 0,057 | 0,439 | 0,414 | APOE/APOC1 | 2 |
| GO:0007603 | phototransduction, visible light | 1/38 | 18/11590 | 0,057 | 0,439 | 0,414 | NMT1 | 1 |

| | | | | | | | | |
|---|---|---|---|---|---|---|---|---|
| GO:0019400 | alditol metabolic process | 1/38 | 18/11590 | 0,057 | 0,439 | 0,414 | GALK1 | 1 |
| GO:0032785 | negative regulation of DNA-templated transcription, elongation | 1/38 | 18/11590 | 0,057 | 0,439 | 0,414 | HEXIM1 | 1 |
| GO:0044154 | histone H3-K14 acetylation | 1/38 | 18/11590 | 0,057 | 0,439 | 0,414 | WBP2 | 1 |
| GO:0045540 | regulation of cholesterol biosynthetic process | 1/38 | 18/11590 | 0,057 | 0,439 | 0,414 | APOE | 1 |
| GO:0046475 | glycerophospholipid catabolic process | 1/38 | 18/11590 | 0,057 | 0,439 | 0,414 | APOC1 | 1 |
| GO:0046782 | regulation of viral transcription | 1/38 | 18/11590 | 0,057 | 0,439 | 0,414 | HEXIM1 | 1 |
| GO:0060252 | positive regulation of glial cell proliferation | 1/38 | 18/11590 | 0,057 | 0,439 | 0,414 | UFL1 | 1 |
| GO:0061615 | glycolytic process through fructose-6-phosphate | 1/38 | 18/11590 | 0,057 | 0,439 | 0,414 | GALK1 | 1 |
| GO:0106118 | regulation of sterol biosynthetic process | 1/38 | 18/11590 | 0,057 | 0,439 | 0,414 | APOE | 1 |
| GO:0043401 | steroid hormone mediated signaling pathway | 2/38 | 120/11590 | 0,059 | 0,439 | 0,414 | UFL1/WBP2 | 2 |
| GO:0055088 | lipid homeostasis | 2/38 | 121/11590 | 0,060 | 0,439 | 0,414 | APOE/ACOX1 | 2 |
| GO:0036498 | IRE1-mediated unfolded protein response | 1/38 | 19/11590 | 0,061 | 0,439 | 0,414 | UFL1 | 1 |
| GO:0050995 | negative regulation of lipid catabolic process | 1/38 | 19/11590 | 0,061 | 0,439 | 0,414 | APOC1 | 1 |
| GO:0051000 | positive regulation of nitric-oxide synthase activity | 1/38 | 19/11590 | 0,061 | 0,439 | 0,414 | APOE | 1 |
| GO:0097062 | dendritic spine maintenance | 1/38 | 19/11590 | 0,061 | 0,439 | 0,414 | APOE | 1 |
| GO:1904357 | negative regulation of telomere maintenance via telomere lengthening | 1/38 | 19/11590 | 0,061 | 0,439 | 0,414 | STN1 | 1 |
| GO:0019216 | regulation of lipid metabolic process | 3/38 | 276/11590 | 0,061 | 0,439 | 0,414 | MTMR9/APOE/APOC1 | 3 |
| GO:0040029 | epigenetic regulation of gene expression | 2/38 | 123/11590 | 0,061 | 0,439 | 0,414 | WBP2/H3-3A | 2 |
| GO:0032331 | negative regulation of chondrocyte differentiation | 1/38 | 20/11590 | 0,064 | 0,439 | 0,414 | EFEMP1 | 1 |
| GO:0051043 | regulation of membrane protein ectodomain proteolysis | 1/38 | 20/11590 | 0,064 | 0,439 | 0,414 | APOE | 1 |
| GO:1901028 | regulation of mitochondrial outer membrane permeabilization involved in apoptotic signaling pathway | 1/38 | 20/11590 | 0,064 | 0,439 | 0,414 | NMT1 | 1 |
| GO:1902992 | negative regulation of amyloid precursor protein catabolic process | 1/38 | 20/11590 | 0,064 | 0,439 | 0,414 | APOE | 1 |

| | | | | | | | | |
|---|---|---|---|---|---|---|---|---|
| GO:1902652 | secondary alcohol metabolic process | 2/38 | 126/11590 | 0,064 | 0,439 | 0,414 | APOE/APOC1 | 2 |
| GO:0032200 | telomere organization | 2/38 | 128/11590 | 0,066 | 0,439 | 0,414 | STN1/H3-3A | 2 |
| GO:0045834 | positive regulation of lipid metabolic process | 2/38 | 128/11590 | 0,066 | 0,439 | 0,414 | APOE/APOC1 | 2 |
| GO:0016125 | sterol metabolic process | 2/38 | 129/11590 | 0,067 | 0,439 | 0,414 | APOE/APOC1 | 2 |
| GO:1903959 | regulation of anion transmembrane transport | 1/38 | 21/11590 | 0,067 | 0,439 | 0,414 | SLC38A1 | 1 |
| GO:0099173 | postsynapse organization | 2/38 | 131/11590 | 0,068 | 0,439 | 0,414 | STAU2/APOE | 2 |
| GO:0006541 | glutamine metabolic process | 1/38 | 22/11590 | 0,070 | 0,439 | 0,414 | SLC38A1 | 1 |
| GO:0010894 | negative regulation of steroid biosynthetic process | 1/38 | 22/11590 | 0,070 | 0,439 | 0,414 | APOE | 1 |
| GO:0031365 | N-terminal protein amino acid modification | 1/38 | 22/11590 | 0,070 | 0,439 | 0,414 | NMT1 | 1 |
| GO:0032801 | receptor catabolic process | 1/38 | 22/11590 | 0,070 | 0,439 | 0,414 | APOE | 1 |
| GO:0046835 | carbohydrate phosphorylation | 1/38 | 22/11590 | 0,070 | 0,439 | 0,414 | GALK1 | 1 |
| GO:0048261 | negative regulation of receptor-mediated endocytosis | 1/38 | 22/11590 | 0,070 | 0,439 | 0,414 | APOC1 | 1 |
| GO:0051957 | positive regulation of amino acid transport | 1/38 | 22/11590 | 0,070 | 0,439 | 0,414 | SLC38A1 | 1 |
| GO:0150146 | cell junction disassembly | 1/38 | 22/11590 | 0,070 | 0,439 | 0,414 | STON1 | 1 |
| GO:1905952 | regulation of lipid localization | 2/38 | 133/11590 | 0,070 | 0,439 | 0,414 | APOE/APOC1 | 2 |
| GO:0006897 | endocytosis | 4/38 | 480/11590 | 0,071 | 0,439 | 0,414 | MTMR9/APOE/APOC1/STON1 | 4 |
| GO:0010976 | positive regulation of neuron projection development | 2/38 | 134/11590 | 0,071 | 0,439 | 0,414 | STAU2/APOE | 2 |
| GO:0006929 | substrate-dependent cell migration | 1/38 | 23/11590 | 0,073 | 0,439 | 0,414 | STON1 | 1 |
| GO:0010875 | positive regulation of cholesterol efflux | 1/38 | 23/11590 | 0,073 | 0,439 | 0,414 | APOE | 1 |
| GO:0032369 | negative regulation of lipid transport | 1/38 | 23/11590 | 0,073 | 0,439 | 0,414 | APOC1 | 1 |
| GO:0070199 | establishment of protein localization to chromosome | 1/38 | 23/11590 | 0,073 | 0,439 | 0,414 | WBP2 | 1 |
| GO:0046486 | glycerolipid metabolic process | 3/38 | 297/11590 | 0,073 | 0,439 | 0,414 | MTMR9/APOE/APOC1 | 3 |
| GO:0048592 | eye morphogenesis | 2/38 | 136/11590 | 0,073 | 0,439 | 0,414 | STAU2/EFEMP1 | 2 |

| | | | | | | | | |
|---|---|---|---|---|---|---|---|---|
| GO:1905475 | regulation of protein localization to membrane | 2/38 | 136/11590 | 0,073 | 0,439 | 0,414 | NMT1/GPC5 | 2 |
| GO:0006066 | alcohol metabolic process | 3/38 | 302/11590 | 0,076 | 0,439 | 0,414 | GALK1/APOE/ APOC1 | 3 |
| GO:0006891 | intra-Golgi vesicle-mediated transport | 1/38 | 24/11590 | 0,076 | 0,439 | 0,414 | COG2 | 1 |
| GO:0035640 | exploration behavior | 1/38 | 24/11590 | 0,076 | 0,439 | 0,414 | APOE | 1 |
| GO:0045939 | negative regulation of steroid metabolic process | 1/38 | 24/11590 | 0,076 | 0,439 | 0,414 | APOE | 1 |
| GO:1900101 | regulation of endoplasmic reticulum unfolded protein response | 1/38 | 24/11590 | 0,076 | 0,439 | 0,414 | UFL1 | 1 |
| GO:0006631 | fatty acid metabolic process | 3/38 | 303/11590 | 0,076 | 0,439 | 0,414 | APOC1/ACOX 1/ACBD4 | 3 |
| GO:0046890 | regulation of lipid biosynthetic process | 2/38 | 141/11590 | 0,078 | 0,439 | 0,414 | APOE/APOC1 | 2 |
| GO:0019068 | virion assembly | 1/38 | 25/11590 | 0,079 | 0,439 | 0,414 | APOE | 1 |
| GO:0034508 | centromere complex assembly | 1/38 | 25/11590 | 0,079 | 0,439 | 0,414 | H3-3A | 1 |
| GO:0045736 | negative regulation of cyclin-dependent protein serine/threonine kinase activity | 1/38 | 25/11590 | 0,079 | 0,439 | 0,414 | HEXIM1 | 1 |
| GO:0055090 | acylglycerol homeostasis | 1/38 | 25/11590 | 0,079 | 0,439 | 0,414 | APOE | 1 |
| GO:0060292 | long-term synaptic depression | 1/38 | 25/11590 | 0,079 | 0,439 | 0,414 | STAU2 | 1 |
| GO:0061037 | negative regulation of cartilage development | 1/38 | 25/11590 | 0,079 | 0,439 | 0,414 | EFEMP1 | 1 |
| GO:0070328 | triglyceride homeostasis | 1/38 | 25/11590 | 0,079 | 0,439 | 0,414 | APOE | 1 |
| GO:1901798 | positive regulation of signal transduction by p53 class mediator | 1/38 | 25/11590 | 0,079 | 0,439 | 0,414 | HEXIM1 | 1 |
| GO:1904030 | negative regulation of cyclin-dependent protein kinase activity | 1/38 | 25/11590 | 0,079 | 0,439 | 0,414 | HEXIM1 | 1 |
| GO:0000038 | very long-chain fatty acid metabolic process | 1/38 | 26/11590 | 0,082 | 0,440 | 0,415 | ACOX1 | 1 |
| GO:0002021 | response to dietary excess | 1/38 | 26/11590 | 0,082 | 0,440 | 0,415 | APOE | 1 |
| GO:0006706 | steroid catabolic process | 1/38 | 26/11590 | 0,082 | 0,440 | 0,415 | APOE | 1 |
| GO:0007263 | nitric oxide mediated signal transduction | 1/38 | 26/11590 | 0,082 | 0,440 | 0,415 | APOE | 1 |
| GO:0051204 | protein insertion into mitochondrial membrane | 1/38 | 26/11590 | 0,082 | 0,440 | 0,415 | NMT1 | 1 |
| GO:0044282 | small molecule catabolic process | 3/38 | 313/11590 | 0,082 | 0,440 | 0,415 | GALK1/APOE/ ACOX1 | 3 |

| | | | | | | | | |
|---|---|---|---|---|---|---|---|---|
| GO:0001556 | oocyte maturation | 1/38 | 27/11590 | 0,085 | 0,440 | 0,415 | H3-3A | 1 |
| GO:0007271 | synaptic transmission, cholinergic | 1/38 | 27/11590 | 0,085 | 0,440 | 0,415 | APOE | 1 |
| GO:0010922 | positive regulation of phosphatase activity | 1/38 | 27/11590 | 0,085 | 0,440 | 0,415 | MTMR9 | 1 |
| GO:0032205 | negative regulation of telomere maintenance | 1/38 | 27/11590 | 0,085 | 0,440 | 0,415 | STN1 | 1 |
| GO:0001941 | postsynaptic membrane organization | 1/38 | 28/11590 | 0,088 | 0,440 | 0,415 | APOE | 1 |
| GO:0046856 | phosphatidylinositol dephosphorylation | 1/38 | 28/11590 | 0,088 | 0,440 | 0,415 | MTMR9 | 1 |
| GO:0055094 | response to lipoprotein particle | 1/38 | 28/11590 | 0,088 | 0,440 | 0,415 | APOE | 1 |
| GO:0090181 | regulation of cholesterol metabolic process | 1/38 | 28/11590 | 0,088 | 0,440 | 0,415 | APOE | 1 |
| GO:0090207 | regulation of triglyceride metabolic process | 1/38 | 28/11590 | 0,088 | 0,440 | 0,415 | APOE | 1 |
| GO:0061136 | regulation of proteasomal protein catabolic process | 2/38 | 152/11590 | 0,088 | 0,440 | 0,415 | UFL1/APOE | 2 |
| GO:0010874 | regulation of cholesterol efflux | 1/38 | 29/11590 | 0,091 | 0,440 | 0,415 | APOE | 1 |
| GO:0032770 | positive regulation of monooxygenase activity | 1/38 | 29/11590 | 0,091 | 0,440 | 0,415 | APOE | 1 |
| GO:0045922 | negative regulation of fatty acid metabolic process | 1/38 | 29/11590 | 0,091 | 0,440 | 0,415 | APOC1 | 1 |
| GO:0050775 | positive regulation of dendrite morphogenesis | 1/38 | 29/11590 | 0,091 | 0,440 | 0,415 | STAU2 | 1 |
| GO:0090151 | establishment of protein localization to mitochondrial membrane | 1/38 | 29/11590 | 0,091 | 0,440 | 0,415 | NMT1 | 1 |
| GO:0007616 | long-term memory | 1/38 | 30/11590 | 0,094 | 0,440 | 0,415 | APOE | 1 |
| GO:0032228 | regulation of synaptic transmission, GABAergic | 1/38 | 30/11590 | 0,094 | 0,440 | 0,415 | SLC38A1 | 1 |
| GO:0097345 | mitochondrial outer membrane permeabilization | 1/38 | 30/11590 | 0,094 | 0,440 | 0,415 | NMT1 | 1 |
| GO:2000279 | negative regulation of DNA biosynthetic process | 1/38 | 30/11590 | 0,094 | 0,440 | 0,415 | STN1 | 1 |
| GO:0050807 | regulation of synapse organization | 2/38 | 158/11590 | 0,094 | 0,440 | 0,415 | STAU2/APOE | 2 |
| GO:0032373 | positive regulation of sterol transport | 1/38 | 31/11590 | 0,097 | 0,440 | 0,415 | APOE | 1 |
| GO:0032376 | positive regulation of cholesterol transport | 1/38 | 31/11590 | 0,097 | 0,440 | 0,415 | APOE | 1 |
| GO:0071168 | protein localization to chromatin | 1/38 | 31/11590 | 0,097 | 0,440 | 0,415 | WBP2 | 1 |
| GO:0071402 | cellular response to lipoprotein particle stimulus | 1/38 | 31/11590 | 0,097 | 0,440 | 0,415 | APOE | 1 |

| | | | | | | | | |
|---|---|---|---|---|---|---|---|---|
| GO:1903749 | positive regulation of establishment of protein localization to mitochondrion | 1/38 | 31/11590 | 0,097 | 0,440 | 0,415 | NMT1 | 1 |
| GO:0050803 | regulation of synapse structure or activity | 2/38 | 161/11590 | 0,097 | 0,440 | 0,415 | STAU2/APOE | 2 |
| GO:0044242 | cellular lipid catabolic process | 2/38 | 163/11590 | 0,100 | 0,440 | 0,415 | APOC1/ACOX 1 | 2 |
| GO:0018149 | peptide cross-linking | 1/38 | 32/11590 | 0,100 | 0,440 | 0,415 | EVPL | 1 |
| GO:1902110 | positive regulation of mitochondrial membrane permeability involved in apoptotic process | 1/38 | 32/11590 | 0,100 | 0,440 | 0,415 | NMT1 | 1 |
| GO:0009755 | hormone-mediated signaling pathway | 2/38 | 164/11590 | 0,101 | 0,440 | 0,415 | UFL1/WBP2 | 2 |
| GO:0045936 | negative regulation of phosphate metabolic process | 3/38 | 342/11590 | 0,101 | 0,440 | 0,415 | APOE/APOC1/ HEXIM1 | 3 |
| GO:0010563 | negative regulation of phosphorus metabolic process | 3/38 | 343/11590 | 0,102 | 0,440 | 0,415 | APOE/APOC1/ HEXIM1 | 3 |
| GO:0009395 | phospholipid catabolic process | 1/38 | 33/11590 | 0,103 | 0,440 | 0,415 | APOC1 | 1 |
| GO:0019934 | cGMP-mediated signaling | 1/38 | 33/11590 | 0,103 | 0,440 | 0,415 | APOE | 1 |
| GO:0035066 | positive regulation of histone acetylation | 1/38 | 33/11590 | 0,103 | 0,440 | 0,415 | WBP2 | 1 |
| GO:0043537 | negative regulation of blood vessel endothelial cell migration | 1/38 | 33/11590 | 0,103 | 0,440 | 0,415 | APOE | 1 |
| GO:0048048 | embryonic eye morphogenesis | 1/38 | 33/11590 | 0,103 | 0,440 | 0,415 | EFEMP1 | 1 |
| GO:0050999 | regulation of nitric-oxide synthase activity | 1/38 | 33/11590 | 0,103 | 0,440 | 0,415 | APOE | 1 |
| GO:0060218 | hematopoietic stem cell differentiation | 1/38 | 33/11590 | 0,103 | 0,440 | 0,415 | UFL1 | 1 |
| GO:0061001 | regulation of dendritic spine morphogenesis | 1/38 | 33/11590 | 0,103 | 0,440 | 0,415 | STAU2 | 1 |
| GO:1902003 | regulation of amyloid-beta formation | 1/38 | 33/11590 | 0,103 | 0,440 | 0,415 | APOE | 1 |
| GO:0006469 | negative regulation of protein kinase activity | 2/38 | 167/11590 | 0,104 | 0,440 | 0,415 | APOE/HEXIM 1 | 2 |
| GO:0007565 | female pregnancy | 2/38 | 168/11590 | 0,105 | 0,440 | 0,415 | SLC38A1/H3-3A | 2 |
| GO:0042554 | superoxide anion generation | 1/38 | 34/11590 | 0,106 | 0,440 | 0,415 | SH3PXD2A | 1 |
| GO:0060251 | regulation of glial cell proliferation | 1/38 | 34/11590 | 0,106 | 0,440 | 0,415 | UFL1 | 1 |

| | | | | | | | | |
|---|---|---|---|---|---|---|---|---|
| GO:1902686 | mitochondrial outer membrane permeabilization involved in programmed cell death | 1/38 | 34/11590 | 0,106 | 0,440 | 0,415 | NMT1 | 1 |
| GO:1903573 | negative regulation of response to endoplasmic reticulum stress | 1/38 | 34/11590 | 0,106 | 0,440 | 0,415 | UFL1 | 1 |
| GO:1990000 | amyloid fibril formation | 1/38 | 34/11590 | 0,106 | 0,440 | 0,415 | APOE | 1 |
| GO:0001503 | ossification | 3/38 | 352/11590 | 0,108 | 0,440 | 0,415 | UFL1/VCAN/H3-3A | 3 |
| GO:0001662 | behavioral fear response | 1/38 | 35/11590 | 0,109 | 0,440 | 0,415 | APOE | 1 |
| GO:0045740 | positive regulation of DNA replication | 1/38 | 35/11590 | 0,109 | 0,440 | 0,415 | STN1 | 1 |
| GO:0097242 | amyloid-beta clearance | 1/38 | 35/11590 | 0,109 | 0,440 | 0,415 | APOE | 1 |
| GO:1902108 | regulation of mitochondrial membrane permeability involved in apoptotic process | 1/38 | 35/11590 | 0,109 | 0,440 | 0,415 | NMT1 | 1 |
| GO:0050804 | modulation of chemical synaptic transmission | 3/38 | 354/11590 | 0,109 | 0,440 | 0,415 | STAU2/SLC38A1/APOE | 3 |
| GO:0099177 | regulation of trans-synaptic signaling | 3/38 | 355/11590 | 0,110 | 0,440 | 0,415 | STAU2/SLC38A1/APOE | 3 |
| GO:0002209 | behavioral defense response | 1/38 | 36/11590 | 0,112 | 0,440 | 0,415 | APOE | 1 |
| GO:0031424 | keratinization | 1/38 | 36/11590 | 0,112 | 0,440 | 0,415 | EVPL | 1 |
| GO:0032570 | response to progesterone | 1/38 | 36/11590 | 0,112 | 0,440 | 0,415 | WBP2 | 1 |
| GO:0046839 | phospholipid dephosphorylation | 1/38 | 36/11590 | 0,112 | 0,440 | 0,415 | MTMR9 | 1 |
| GO:0042180 | cellular ketone metabolic process | 2/38 | 176/11590 | 0,113 | 0,440 | 0,415 | APOC1/NMT1 | 2 |
| GO:0001937 | negative regulation of endothelial cell proliferation | 1/38 | 37/11590 | 0,115 | 0,440 | 0,415 | APOE | 1 |
| GO:0007602 | phototransduction | 1/38 | 37/11590 | 0,115 | 0,440 | 0,415 | NMT1 | 1 |
| GO:0016233 | telomere capping | 1/38 | 37/11590 | 0,115 | 0,440 | 0,415 | STN1 | 1 |
| GO:0019320 | hexose catabolic process | 1/38 | 37/11590 | 0,115 | 0,440 | 0,415 | GALK1 | 1 |
| GO:0035794 | positive regulation of mitochondrial membrane permeability | 1/38 | 37/11590 | 0,115 | 0,440 | 0,415 | NMT1 | 1 |
| GO:0044788 | modulation by host of viral process | 1/38 | 37/11590 | 0,115 | 0,440 | 0,415 | APOE | 1 |
| GO:0045806 | negative regulation of endocytosis | 1/38 | 37/11590 | 0,115 | 0,440 | 0,415 | APOC1 | 1 |

| | | | | | | | | |
|---|---|---|---|---|---|---|---|---|
| GO:1903050 | regulation of proteolysis involved in protein catabolic process | 2/38 | 178/11590 | 0,115 | 0,440 | 0,415 | UFL1/APOE | 2 |
| GO:0071383 | cellular response to steroid hormone stimulus | 2/38 | 179/11590 | 0,116 | 0,440 | 0,415 | UFL1/WBP2 | 2 |
| GO:0006509 | membrane protein ectodomain proteolysis | 1/38 | 38/11590 | 0,117 | 0,440 | 0,415 | APOE | 1 |
| GO:0006692 | prostanoid metabolic process | 1/38 | 38/11590 | 0,117 | 0,440 | 0,415 | ACOX1 | 1 |
| GO:0006693 | prostaglandin metabolic process | 1/38 | 38/11590 | 0,117 | 0,440 | 0,415 | ACOX1 | 1 |
| GO:0009584 | detection of visible light | 1/38 | 38/11590 | 0,117 | 0,440 | 0,415 | NMT1 | 1 |
| GO:0097178 | ruffle assembly | 1/38 | 38/11590 | 0,117 | 0,440 | 0,415 | STON1 | 1 |
| GO:1900271 | regulation of long-term synaptic potentiation | 1/38 | 38/11590 | 0,117 | 0,440 | 0,415 | APOE | 1 |
| GO:1905953 | negative regulation of lipid localization | 1/38 | 38/11590 | 0,117 | 0,440 | 0,415 | APOC1 | 1 |
| GO:2000758 | positive regulation of peptidyl-lysine acetylation | 1/38 | 38/11590 | 0,117 | 0,440 | 0,415 | WBP2 | 1 |
| GO:0042304 | regulation of fatty acid biosynthetic process | 1/38 | 39/11590 | 0,120 | 0,440 | 0,415 | APOC1 | 1 |
| GO:0046365 | monosaccharide catabolic process | 1/38 | 39/11590 | 0,120 | 0,440 | 0,415 | GALK1 | 1 |
| GO:0051954 | positive regulation of amine transport | 1/38 | 39/11590 | 0,120 | 0,440 | 0,415 | SLC38A1 | 1 |
| GO:0120009 | intermembrane lipid transfer | 1/38 | 39/11590 | 0,120 | 0,440 | 0,415 | APOE | 1 |
| GO:1902991 | regulation of amyloid precursor protein catabolic process | 1/38 | 39/11590 | 0,120 | 0,440 | 0,415 | APOE | 1 |
| GO:0042743 | hydrogen peroxide metabolic process | 1/38 | 40/11590 | 0,123 | 0,440 | 0,415 | ACOX1 | 1 |
| GO:0051955 | regulation of amino acid transport | 1/38 | 40/11590 | 0,123 | 0,440 | 0,415 | SLC38A1 | 1 |
| GO:0033673 | negative regulation of kinase activity | 2/38 | 186/11590 | 0,124 | 0,440 | 0,415 | APOE/HEXIM1 | 2 |
| GO:0044703 | multi-organism reproductive process | 2/38 | 186/11590 | 0,124 | 0,440 | 0,415 | SLC38A1/H3-3A | 2 |
| GO:0060828 | regulation of canonical Wnt signaling pathway | 2/38 | 188/11590 | 0,126 | 0,440 | 0,415 | APOE/GPC5 | 2 |
| GO:0015804 | neutral amino acid transport | 1/38 | 41/11590 | 0,126 | 0,440 | 0,415 | SLC38A1 | 1 |
| GO:0030261 | chromosome condensation | 1/38 | 41/11590 | 0,126 | 0,440 | 0,415 | H3-3A | 1 |
| GO:0032892 | positive regulation of organic acid transport | 1/38 | 41/11590 | 0,126 | 0,440 | 0,415 | SLC38A1 | 1 |
| GO:0034205 | amyloid-beta formation | 1/38 | 41/11590 | 0,126 | 0,440 | 0,415 | APOE | 1 |
| GO:0042596 | fear response | 1/38 | 41/11590 | 0,126 | 0,440 | 0,415 | APOE | 1 |

| | | | | | | | | |
|---|---|---|---|---|---|---|---|---|
| GO:1905710 | positive regulation of membrane permeability | 1/38 | 41/11590 | 0,126 | 0,440 | 0,415 | NMT1 | 1 |
| GO:0072593 | reactive oxygen species metabolic process | 2/38 | 190/11590 | 0,128 | 0,440 | 0,415 | SH3PXD2A/A COX1 | 2 |
| GO:0001504 | neurotransmitter uptake | 1/38 | 42/11590 | 0,129 | 0,440 | 0,415 | SLC38A1 | 1 |
| GO:1903747 | regulation of establishment of protein localization to mitochondrion | 1/38 | 42/11590 | 0,129 | 0,440 | 0,415 | NMT1 | 1 |
| GO:0031330 | negative regulation of cellular catabolic process | 2/38 | 192/11590 | 0,130 | 0,440 | 0,415 | MTMR9/APO C1 | 2 |
| GO:0044706 | multi-multicellular organism process | 2/38 | 192/11590 | 0,130 | 0,440 | 0,415 | SLC38A1/H3-3A | 2 |
| GO:0045088 | regulation of innate immune response | 2/38 | 193/11590 | 0,132 | 0,440 | 0,415 | APOE/HEXIM 1 | 2 |
| GO:0032210 | regulation of telomere maintenance via telomerase | 1/38 | 43/11590 | 0,132 | 0,440 | 0,415 | STN1 | 1 |
| GO:0032330 | regulation of chondrocyte differentiation | 1/38 | 43/11590 | 0,132 | 0,440 | 0,415 | EFEMP1 | 1 |
| GO:0046503 | glycerolipid catabolic process | 1/38 | 43/11590 | 0,132 | 0,440 | 0,415 | APOC1 | 1 |
| GO:0051965 | positive regulation of synapse assembly | 1/38 | 43/11590 | 0,132 | 0,440 | 0,415 | STAU2 | 1 |
| GO:0016358 | dendrite development | 2/38 | 194/11590 | 0,133 | 0,440 | 0,415 | STAU2/APOE | 2 |
| GO:0007281 | germ cell development | 2/38 | 195/11590 | 0,134 | 0,440 | 0,415 | STAU2/H3-3A | 2 |
| GO:0051489 | regulation of filopodium assembly | 1/38 | 44/11590 | 0,135 | 0,440 | 0,415 | STAU2 | 1 |
| GO:1903793 | positive regulation of anion transport | 1/38 | 44/11590 | 0,135 | 0,440 | 0,415 | SLC38A1 | 1 |
| GO:0032768 | regulation of monooxygenase activity | 1/38 | 45/11590 | 0,138 | 0,440 | 0,415 | APOE | 1 |
| GO:0043114 | regulation of vascular permeability | 1/38 | 45/11590 | 0,138 | 0,440 | 0,415 | APOE | 1 |
| GO:0048168 | regulation of neuronal synaptic plasticity | 1/38 | 45/11590 | 0,138 | 0,440 | 0,415 | APOE | 1 |
| GO:0089718 | amino acid import across plasma membrane | 1/38 | 45/11590 | 0,138 | 0,440 | 0,415 | SLC38A1 | 1 |
| GO:0050769 | positive regulation of neurogenesis | 2/38 | 199/11590 | 0,138 | 0,440 | 0,415 | UFL1/STAU2 | 2 |
| GO:0033044 | regulation of chromosome organization | 2/38 | 200/11590 | 0,139 | 0,440 | 0,415 | STN1/H3-3A | 2 |
| GO:0007566 | embryo implantation | 1/38 | 46/11590 | 0,140 | 0,440 | 0,415 | H3-3A | 1 |
| GO:0010543 | regulation of platelet activation | 1/38 | 46/11590 | 0,140 | 0,440 | 0,415 | APOE | 1 |
| GO:0014009 | glial cell proliferation | 1/38 | 46/11590 | 0,140 | 0,440 | 0,415 | UFL1 | 1 |
| GO:0042311 | vasodilation | 1/38 | 46/11590 | 0,140 | 0,440 | 0,415 | APOE | 1 |

| | | | | | | | | |
|---|---|---|---|---|---|---|---|---|
| GO:0043113 | receptor clustering | 1/38 | 46/11590 | 0,140 | 0,440 | 0,415 | APOE | 1 |
| GO:0046902 | regulation of mitochondrial membrane permeability | 1/38 | 46/11590 | 0,140 | 0,440 | 0,415 | NMT1 | 1 |
| GO:0048599 | oocyte development | 1/38 | 46/11590 | 0,140 | 0,440 | 0,415 | H3-3A | 1 |
| GO:1901985 | positive regulation of protein acetylation | 1/38 | 46/11590 | 0,140 | 0,440 | 0,415 | WBP2 | 1 |
| GO:1903409 | reactive oxygen species biosynthetic process | 1/38 | 46/11590 | 0,140 | 0,440 | 0,415 | ACOX1 | 1 |
| GO:0031400 | negative regulation of protein modification process | 3/38 | 398/11590 | 0,141 | 0,440 | 0,415 | UFL1/APOE/HEXIM1 | 3 |
| GO:0006354 | DNA-templated transcription elongation | 2/38 | 202/11590 | 0,142 | 0,440 | 0,415 | FHL5/HEXIM1 | 2 |
| GO:0006695 | cholesterol biosynthetic process | 1/38 | 47/11590 | 0,143 | 0,440 | 0,415 | APOE | 1 |
| GO:0009994 | oocyte differentiation | 1/38 | 47/11590 | 0,143 | 0,440 | 0,415 | H3-3A | 1 |
| GO:0030195 | negative regulation of blood coagulation | 1/38 | 47/11590 | 0,143 | 0,440 | 0,415 | APOE | 1 |
| GO:0033619 | membrane protein proteolysis | 1/38 | 47/11590 | 0,143 | 0,440 | 0,415 | APOE | 1 |
| GO:0046164 | alcohol catabolic process | 1/38 | 47/11590 | 0,143 | 0,440 | 0,415 | APOE | 1 |
| GO:0060997 | dendritic spine morphogenesis | 1/38 | 47/11590 | 0,143 | 0,440 | 0,415 | STAU2 | 1 |
| GO:1902653 | secondary alcohol biosynthetic process | 1/38 | 47/11590 | 0,143 | 0,440 | 0,415 | APOE | 1 |
| GO:1902930 | regulation of alcohol biosynthetic process | 1/38 | 47/11590 | 0,143 | 0,440 | 0,415 | APOE | 1 |
| GO:1904356 | regulation of telomere maintenance via telomere lengthening | 1/38 | 47/11590 | 0,143 | 0,440 | 0,415 | STN1 | 1 |
| GO:0010596 | negative regulation of endothelial cell migration | 1/38 | 48/11590 | 0,146 | 0,440 | 0,415 | APOE | 1 |
| GO:0019083 | viral transcription | 1/38 | 48/11590 | 0,146 | 0,440 | 0,415 | HEXIM1 | 1 |
| GO:0035306 | positive regulation of dephosphorylation | 1/38 | 48/11590 | 0,146 | 0,440 | 0,415 | MTMR9 | 1 |
| GO:0048662 | negative regulation of smooth muscle cell proliferation | 1/38 | 48/11590 | 0,146 | 0,440 | 0,415 | APOE | 1 |
| GO:0050709 | negative regulation of protein secretion | 1/38 | 48/11590 | 0,146 | 0,440 | 0,415 | APOE | 1 |
| GO:0050994 | regulation of lipid catabolic process | 1/38 | 48/11590 | 0,146 | 0,440 | 0,415 | APOC1 | 1 |
| GO:0051932 | synaptic transmission, GABAergic | 1/38 | 48/11590 | 0,146 | 0,440 | 0,415 | SLC38A1 | 1 |
| GO:1900047 | negative regulation of hemostasis | 1/38 | 48/11590 | 0,146 | 0,440 | 0,415 | APOE | 1 |
| GO:0043543 | protein acylation | 2/38 | 208/11590 | 0,148 | 0,440 | 0,415 | WBP2/NMT1 | 2 |

| | | | | | | | | |
|---|---|---|---|---|---|---|---|---|
| GO:0051348 | negative regulation of transferase activity | 2/38 | 208/11590 | 0,148 | 0,440 | 0,415 | APOE/HEXIM 1 | 2 |
| GO:0009583 | detection of light stimulus | 1/38 | 49/11590 | 0,149 | 0,440 | 0,415 | NMT1 | 1 |
| GO:0043112 | receptor metabolic process | 1/38 | 49/11590 | 0,149 | 0,440 | 0,415 | APOE | 1 |
| GO:0043407 | negative regulation of MAP kinase activity | 1/38 | 49/11590 | 0,149 | 0,440 | 0,415 | APOE | 1 |
| GO:0050819 | negative regulation of coagulation | 1/38 | 49/11590 | 0,149 | 0,440 | 0,415 | APOE | 1 |
| GO:0000768 | syncytium formation by plasma membrane fusion | 1/38 | 50/11590 | 0,152 | 0,440 | 0,415 | SH3PXD2A | 1 |
| GO:0002218 | activation of innate immune response | 1/38 | 50/11590 | 0,152 | 0,440 | 0,415 | HEXIM1 | 1 |
| GO:0031529 | ruffle organization | 1/38 | 50/11590 | 0,152 | 0,440 | 0,415 | STON1 | 1 |
| GO:0033866 | nucleoside bisphosphate biosynthetic process | 1/38 | 50/11590 | 0,152 | 0,440 | 0,415 | DCAKD | 1 |
| GO:0034030 | ribonucleoside bisphosphate biosynthetic process | 1/38 | 50/11590 | 0,152 | 0,440 | 0,415 | DCAKD | 1 |
| GO:0034033 | purine nucleoside bisphosphate biosynthetic process | 1/38 | 50/11590 | 0,152 | 0,440 | 0,415 | DCAKD | 1 |
| GO:0050435 | amyloid-beta metabolic process | 1/38 | 50/11590 | 0,152 | 0,440 | 0,415 | APOE | 1 |
| GO:0051205 | protein insertion into membrane | 1/38 | 50/11590 | 0,152 | 0,440 | 0,415 | NMT1 | 1 |
| GO:0140253 | cell-cell fusion | 1/38 | 50/11590 | 0,152 | 0,440 | 0,415 | SH3PXD2A | 1 |
| GO:0051353 | positive regulation of oxidoreductase activity | 1/38 | 51/11590 | 0,155 | 0,446 | 0,421 | APOE | 1 |
| GO:0006898 | receptor-mediated endocytosis | 2/38 | 214/11590 | 0,155 | 0,446 | 0,421 | APOE/APOC1 | 2 |
| GO:0015850 | organic hydroxy compound transport | 2/38 | 214/11590 | 0,155 | 0,446 | 0,421 | APOE/APOC1 | 2 |
| GO:0090596 | sensory organ morphogenesis | 2/38 | 215/11590 | 0,156 | 0,447 | 0,421 | STAU2/EFEM P1 | 2 |
| GO:0042987 | amyloid precursor protein catabolic process | 1/38 | 52/11590 | 0,157 | 0,447 | 0,421 | APOE | 1 |
| GO:0043954 | cellular component maintenance | 1/38 | 52/11590 | 0,157 | 0,447 | 0,421 | APOE | 1 |
| GO:0048008 | platelet-derived growth factor receptor signaling pathway | 1/38 | 52/11590 | 0,157 | 0,447 | 0,421 | STON1 | 1 |
| GO:0006949 | syncytium formation | 1/38 | 53/11590 | 0,160 | 0,451 | 0,425 | SH3PXD2A | 1 |
| GO:0048488 | synaptic vesicle endocytosis | 1/38 | 53/11590 | 0,160 | 0,451 | 0,425 | STON1 | 1 |
| GO:0140238 | presynaptic endocytosis | 1/38 | 53/11590 | 0,160 | 0,451 | 0,425 | STON1 | 1 |
| GO:0016126 | sterol biosynthetic process | 1/38 | 54/11590 | 0,163 | 0,457 | 0,431 | APOE | 1 |

| | | | | | | | | |
|---|---|---|---|---|---|---|---|---|
| GO:0015909 | long-chain fatty acid transport | 1/38 | 55/11590 | 0,166 | 0,464 | 0,437 | APOE | 1 |
| GO:0006801 | superoxide metabolic process | 1/38 | 56/11590 | 0,168 | 0,466 | 0,440 | SH3PXD2A | 1 |
| GO:0035065 | regulation of histone acetylation | 1/38 | 56/11590 | 0,168 | 0,466 | 0,440 | WBP2 | 1 |
| GO:0046847 | filopodium assembly | 1/38 | 56/11590 | 0,168 | 0,466 | 0,440 | STAU2 | 1 |
| GO:0048814 | regulation of dendrite morphogenesis | 1/38 | 56/11590 | 0,168 | 0,466 | 0,440 | STAU2 | 1 |
| GO:0007004 | telomere maintenance via telomerase | 1/38 | 57/11590 | 0,171 | 0,470 | 0,443 | STN1 | 1 |
| GO:0035418 | protein localization to synapse | 1/38 | 57/11590 | 0,171 | 0,470 | 0,443 | STAU2 | 1 |
| GO:0006278 | RNA-templated DNA biosynthetic process | 1/38 | 58/11590 | 0,174 | 0,470 | 0,443 | STN1 | 1 |
| GO:0014015 | positive regulation of gliogenesis | 1/38 | 58/11590 | 0,174 | 0,470 | 0,443 | UFL1 | 1 |
| GO:0015807 | L-amino acid transport | 1/38 | 58/11590 | 0,174 | 0,470 | 0,443 | SLC38A1 | 1 |
| GO:0043627 | response to estrogen | 1/38 | 58/11590 | 0,174 | 0,470 | 0,443 | WBP2 | 1 |
| GO:0090559 | regulation of membrane permeability | 1/38 | 58/11590 | 0,174 | 0,470 | 0,443 | NMT1 | 1 |
| GO:0060627 | regulation of vesicle-mediated transport | 3/38 | 440/11590 | 0,174 | 0,470 | 0,443 | APOE/APOC1/ STON1 | 3 |
| GO:0051962 | positive regulation of nervous system development | 2/38 | 231/11590 | 0,175 | 0,470 | 0,443 | UFL1/STAU2 | 2 |
| GO:0030522 | intracellular receptor signaling pathway | 2/38 | 232/11590 | 0,176 | 0,470 | 0,443 | UFL1/WBP2 | 2 |
| GO:0060070 | canonical Wnt signaling pathway | 2/38 | 232/11590 | 0,176 | 0,470 | 0,443 | APOE/GPC5 | 2 |
| GO:0050805 | negative regulation of synaptic transmission | 1/38 | 59/11590 | 0,177 | 0,470 | 0,443 | STAU2 | 1 |
| GO:0061035 | regulation of cartilage development | 1/38 | 59/11590 | 0,177 | 0,470 | 0,443 | EFEMP1 | 1 |
| GO:1902475 | L-alpha-amino acid transmembrane transport | 1/38 | 59/11590 | 0,177 | 0,470 | 0,443 | SLC38A1 | 1 |
| GO:0006650 | glycerophospholipid metabolic process | 2/38 | 234/11590 | 0,178 | 0,473 | 0,446 | MTMR9/APO C1 | 2 |
| GO:0008088 | axo-dendritic transport | 1/38 | 60/11590 | 0,179 | 0,473 | 0,446 | STAU2 | 1 |
| GO:0050795 | regulation of behavior | 1/38 | 60/11590 | 0,179 | 0,473 | 0,446 | APOE | 1 |
| GO:0072657 | protein localization to membrane | 3/38 | 447/11590 | 0,180 | 0,473 | 0,446 | APOE/NMT1/ GPC5 | 3 |
| GO:0010633 | negative regulation of epithelial cell migration | 1/38 | 61/11590 | 0,182 | 0,474 | 0,447 | APOE | 1 |
| GO:0030968 | endoplasmic reticulum unfolded protein response | 1/38 | 61/11590 | 0,182 | 0,474 | 0,447 | UFL1 | 1 |

| | | | | | | | | |
|---|---|---|---|---|---|---|---|---|
| GO:0046470 | phosphatidylcholine metabolic process | 1/38 | 61/11590 | 0,182 | 0,474 | 0,447 | APOC1 | 1 |
| GO:0050810 | regulation of steroid biosynthetic process | 1/38 | 61/11590 | 0,182 | 0,474 | 0,447 | APOE | 1 |
| GO:0006334 | nucleosome assembly | 1/38 | 62/11590 | 0,185 | 0,476 | 0,449 | H3-3A | 1 |
| GO:0009064 | glutamine family amino acid metabolic process | 1/38 | 62/11590 | 0,185 | 0,476 | 0,449 | SLC38A1 | 1 |
| GO:0010822 | positive regulation of mitochondrion organization | 1/38 | 62/11590 | 0,185 | 0,476 | 0,449 | NMT1 | 1 |
| GO:0036465 | synaptic vesicle recycling | 1/38 | 62/11590 | 0,185 | 0,476 | 0,449 | STON1 | 1 |
| GO:0051056 | regulation of small GTPase mediated signal transduction | 2/38 | 240/11590 | 0,186 | 0,477 | 0,450 | APOE/ARHGEF12 | 2 |
| GO:0010921 | regulation of phosphatase activity | 1/38 | 63/11590 | 0,187 | 0,477 | 0,450 | MTMR9 | 1 |
| GO:2000756 | regulation of peptidyl-lysine acetylation | 1/38 | 63/11590 | 0,187 | 0,477 | 0,450 | WBP2 | 1 |
| GO:0030111 | regulation of Wnt signaling pathway | 2/38 | 243/11590 | 0,189 | 0,477 | 0,450 | APOE/GPC5 | 2 |
| GO:0010833 | telomere maintenance via telomere lengthening | 1/38 | 64/11590 | 0,190 | 0,477 | 0,450 | STN1 | 1 |
| GO:0031507 | heterochromatin formation | 1/38 | 64/11590 | 0,190 | 0,477 | 0,450 | H3-3A | 1 |
| GO:0003018 | vascular process in circulatory system | 2/38 | 244/11590 | 0,190 | 0,477 | 0,450 | SLC38A1/APOE | 2 |
| GO:0006497 | protein lipidation | 1/38 | 65/11590 | 0,193 | 0,477 | 0,450 | NMT1 | 1 |
| GO:0006635 | fatty acid beta-oxidation | 1/38 | 65/11590 | 0,193 | 0,477 | 0,450 | ACOX1 | 1 |
| GO:0048844 | artery morphogenesis | 1/38 | 65/11590 | 0,193 | 0,477 | 0,450 | APOE | 1 |
| GO:1901615 | organic hydroxy compound metabolic process | 3/38 | 463/11590 | 0,193 | 0,477 | 0,450 | GALK1/APOE/APOC1 | 3 |
| GO:0015849 | organic acid transport | 2/38 | 248/11590 | 0,195 | 0,477 | 0,450 | SLC38A1/APOE | 2 |
| GO:0008652 | cellular amino acid biosynthetic process | 1/38 | 66/11590 | 0,195 | 0,477 | 0,450 | SLC38A1 | 1 |
| GO:0019915 | lipid storage | 1/38 | 66/11590 | 0,195 | 0,477 | 0,450 | APOE | 1 |
| GO:0030193 | regulation of blood coagulation | 1/38 | 66/11590 | 0,195 | 0,477 | 0,450 | APOE | 1 |
| GO:0031397 | negative regulation of protein ubiquitination | 1/38 | 66/11590 | 0,195 | 0,477 | 0,450 | UFL1 | 1 |
| GO:0032088 | negative regulation of NF-kappaB transcription factor activity | 1/38 | 66/11590 | 0,195 | 0,477 | 0,450 | UFL1 | 1 |

| | | | | | | | | |
|---|---|---|---|---|---|---|---|---|
| GO:0043966 | histone H3 acetylation | 1/38 | 66/11590 | 0,195 | 0,477 | 0,450 | WBP2 | 1 |
| GO:0061045 | negative regulation of wound healing | 1/38 | 66/11590 | 0,195 | 0,477 | 0,450 | APOE | 1 |
| GO:1901616 | organic hydroxy compound catabolic process | 1/38 | 66/11590 | 0,195 | 0,477 | 0,450 | APOE | 1 |
| GO:1905897 | regulation of response to endoplasmic reticulum stress | 1/38 | 66/11590 | 0,195 | 0,477 | 0,450 | UFL1 | 1 |
| GO:0046394 | carboxylic acid biosynthetic process | 2/38 | 250/11590 | 0,197 | 0,479 | 0,452 | SLC38A1/APOC1 | 2 |
| GO:0032890 | regulation of organic acid transport | 1/38 | 67/11590 | 0,198 | 0,479 | 0,452 | SLC38A1 | 1 |
| GO:0042982 | amyloid precursor protein metabolic process | 1/38 | 67/11590 | 0,198 | 0,479 | 0,452 | APOE | 1 |
| GO:0061912 | selective autophagy | 1/38 | 67/11590 | 0,198 | 0,479 | 0,452 | UFL1 | 1 |
| GO:0044283 | small molecule biosynthetic process | 3/38 | 470/11590 | 0,199 | 0,480 | 0,453 | SLC38A1/APOE/APOC1 | 3 |
| GO:0016053 | organic acid biosynthetic process | 2/38 | 252/11590 | 0,200 | 0,481 | 0,453 | SLC38A1/APOC1 | 2 |
| GO:0051851 | modulation by host of symbiont process | 1/38 | 68/11590 | 0,201 | 0,481 | 0,453 | APOE | 1 |
| GO:1900046 | regulation of hemostasis | 1/38 | 68/11590 | 0,201 | 0,481 | 0,453 | APOE | 1 |
| GO:0034243 | regulation of transcription elongation by RNA polymerase II | 1/38 | 69/11590 | 0,203 | 0,484 | 0,456 | HEXIM1 | 1 |
| GO:0046889 | positive regulation of lipid biosynthetic process | 1/38 | 69/11590 | 0,203 | 0,484 | 0,456 | APOE | 1 |
| GO:0050818 | regulation of coagulation | 1/38 | 69/11590 | 0,203 | 0,484 | 0,456 | APOE | 1 |
| GO:0010507 | negative regulation of autophagy | 1/38 | 70/11590 | 0,206 | 0,484 | 0,457 | MTMR9 | 1 |
| GO:0042632 | cholesterol homeostasis | 1/38 | 70/11590 | 0,206 | 0,484 | 0,457 | APOE | 1 |
| GO:0045104 | intermediate filament cytoskeleton organization | 1/38 | 70/11590 | 0,206 | 0,484 | 0,457 | EVPL | 1 |
| GO:0048477 | oogenesis | 1/38 | 70/11590 | 0,206 | 0,484 | 0,457 | H3-3A | 1 |
| GO:0001933 | negative regulation of protein phosphorylation | 2/38 | 259/11590 | 0,208 | 0,484 | 0,457 | APOE/HEXIM1 | 2 |
| GO:0009895 | negative regulation of catabolic process | 2/38 | 259/11590 | 0,208 | 0,484 | 0,457 | MTMR9/APOC1 | 2 |
| GO:0045103 | intermediate filament-based process | 1/38 | 71/11590 | 0,209 | 0,484 | 0,457 | EVPL | 1 |
| GO:0055092 | sterol homeostasis | 1/38 | 71/11590 | 0,209 | 0,484 | 0,457 | APOE | 1 |

| | | | | | | | | |
|---|---|---|---|---|---|---|---|---|
| GO:0070828 | heterochromatin organization | 1/38 | 71/11590 | 0,209 | 0,484 | 0,457 | H3-3A | 1 |
| GO:0098869 | cellular oxidant detoxification | 1/38 | 71/11590 | 0,209 | 0,484 | 0,457 | APOE | 1 |
| GO:0010770 | positive regulation of cell morphogenesis involved in differentiation | 1/38 | 72/11590 | 0,211 | 0,486 | 0,458 | STAU2 | 1 |
| GO:0045814 | negative regulation of gene expression, epigenetic | 1/38 | 72/11590 | 0,211 | 0,486 | 0,458 | H3-3A | 1 |
| GO:0051963 | regulation of synapse assembly | 1/38 | 72/11590 | 0,211 | 0,486 | 0,458 | STAU2 | 1 |
| GO:1901983 | regulation of protein acetylation | 1/38 | 72/11590 | 0,211 | 0,486 | 0,458 | WBP2 | 1 |
| GO:0033555 | multicellular organismal response to stress | 1/38 | 73/11590 | 0,214 | 0,489 | 0,462 | APOE | 1 |
| GO:1903321 | negative regulation of protein modification by small protein conjugation or removal | 1/38 | 73/11590 | 0,214 | 0,489 | 0,462 | UFL1 | 1 |
| GO:0032370 | positive regulation of lipid transport | 1/38 | 74/11590 | 0,216 | 0,492 | 0,464 | APOE | 1 |
| GO:0035023 | regulation of Rho protein signal transduction | 1/38 | 74/11590 | 0,216 | 0,492 | 0,464 | APOE | 1 |
| GO:0060291 | long-term synaptic potentiation | 1/38 | 74/11590 | 0,216 | 0,492 | 0,464 | APOE | 1 |
| GO:0007276 | gamete generation | 3/38 | 491/11590 | 0,217 | 0,492 | 0,464 | STAU2/H3-3A/ACOX1 | 3 |
| GO:0006338 | chromatin remodeling | 2/38 | 268/11590 | 0,219 | 0,495 | 0,467 | WBP2/H3-3A | 2 |
| GO:0008202 | steroid metabolic process | 2/38 | 268/11590 | 0,219 | 0,495 | 0,467 | APOE/APOC1 | 2 |
| GO:0010720 | positive regulation of cell development | 2/38 | 269/11590 | 0,220 | 0,496 | 0,467 | UFL1/STAU2 | 2 |
| GO:0006096 | glycolytic process | 1/38 | 76/11590 | 0,222 | 0,496 | 0,467 | GALK1 | 1 |
| GO:0006757 | ATP generation from ADP | 1/38 | 76/11590 | 0,222 | 0,496 | 0,467 | GALK1 | 1 |
| GO:0062012 | regulation of small molecule metabolic process | 2/38 | 271/11590 | 0,223 | 0,496 | 0,467 | APOE/APOC1 | 2 |
| GO:0019217 | regulation of fatty acid metabolic process | 1/38 | 77/11590 | 0,224 | 0,496 | 0,467 | APOC1 | 1 |
| GO:0019935 | cyclic-nucleotide-mediated signaling | 1/38 | 77/11590 | 0,224 | 0,496 | 0,467 | APOE | 1 |
| GO:0032204 | regulation of telomere maintenance | 1/38 | 77/11590 | 0,224 | 0,496 | 0,467 | STN1 | 1 |
| GO:0048041 | focal adhesion assembly | 1/38 | 77/11590 | 0,224 | 0,496 | 0,467 | STON1 | 1 |
| GO:1903035 | negative regulation of response to wounding | 1/38 | 77/11590 | 0,224 | 0,496 | 0,467 | APOE | 1 |
| GO:0003333 | amino acid transmembrane transport | 1/38 | 78/11590 | 0,227 | 0,497 | 0,469 | SLC38A1 | 1 |
| GO:0034620 | cellular response to unfolded protein | 1/38 | 78/11590 | 0,227 | 0,497 | 0,469 | UFL1 | 1 |

| | | | | | | | | |
|---|---|---|---|---|---|---|---|---|
| GO:0060191 | regulation of lipase activity | 1/38 | 78/11590 | 0,227 | 0,497 | 0,469 | APOC1 | 1 |
| GO:0022412 | cellular process involved in reproduction in multicellular organism | 2/38 | 275/11590 | 0,228 | 0,497 | 0,469 | STAU2/H3-3A | 2 |
| GO:0010506 | regulation of autophagy | 2/38 | 276/11590 | 0,229 | 0,497 | 0,469 | UFL1/MTMR9 | 2 |
| GO:0015908 | fatty acid transport | 1/38 | 79/11590 | 0,229 | 0,497 | 0,469 | APOE | 1 |
| GO:0030516 | regulation of axon extension | 1/38 | 79/11590 | 0,229 | 0,497 | 0,469 | APOE | 1 |
| GO:0032092 | positive regulation of protein binding | 1/38 | 79/11590 | 0,229 | 0,497 | 0,469 | APOE | 1 |
| GO:0044070 | regulation of anion transport | 1/38 | 79/11590 | 0,229 | 0,497 | 0,469 | SLC38A1 | 1 |
| GO:0009791 | post-embryonic development | 1/38 | 80/11590 | 0,232 | 0,500 | 0,472 | EFEMP1 | 1 |
| GO:0048525 | negative regulation of viral process | 1/38 | 80/11590 | 0,232 | 0,500 | 0,472 | HEXIM1 | 1 |
| GO:0009062 | fatty acid catabolic process | 1/38 | 81/11590 | 0,234 | 0,501 | 0,473 | ACOX1 | 1 |
| GO:0034502 | protein localization to chromosome | 1/38 | 81/11590 | 0,234 | 0,501 | 0,473 | WBP2 | 1 |
| GO:0045807 | positive regulation of endocytosis | 1/38 | 81/11590 | 0,234 | 0,501 | 0,473 | APOE | 1 |
| GO:1905477 | positive regulation of protein localization to membrane | 1/38 | 81/11590 | 0,234 | 0,501 | 0,473 | NMT1 | 1 |
| GO:0046031 | ADP metabolic process | 1/38 | 82/11590 | 0,237 | 0,503 | 0,474 | GALK1 | 1 |
| GO:1901890 | positive regulation of cell junction assembly | 1/38 | 82/11590 | 0,237 | 0,503 | 0,474 | STAU2 | 1 |
| GO:1902414 | protein localization to cell junction | 1/38 | 82/11590 | 0,237 | 0,503 | 0,474 | STAU2 | 1 |
| GO:0000079 | regulation of cyclin-dependent protein serine/threonine kinase activity | 1/38 | 83/11590 | 0,239 | 0,503 | 0,474 | HEXIM1 | 1 |
| GO:0030316 | osteoclast differentiation | 1/38 | 83/11590 | 0,239 | 0,503 | 0,474 | SH3PXD2A | 1 |
| GO:0050773 | regulation of dendrite development | 1/38 | 83/11590 | 0,239 | 0,503 | 0,474 | STAU2 | 1 |
| GO:0090263 | positive regulation of canonical Wnt signaling pathway | 1/38 | 83/11590 | 0,239 | 0,503 | 0,474 | GPC5 | 1 |
| GO:0002831 | regulation of response to biotic stimulus | 2/38 | 285/11590 | 0,240 | 0,503 | 0,474 | APOE/HEXIM 1 | 2 |
| GO:0007265 | Ras protein signal transduction | 2/38 | 285/11590 | 0,240 | 0,503 | 0,474 | APOE/ARHGE F12 | 2 |
| GO:0032784 | regulation of DNA-templated transcription elongation | 1/38 | 84/11590 | 0,242 | 0,503 | 0,475 | HEXIM1 | 1 |
| GO:0051341 | regulation of oxidoreductase activity | 1/38 | 84/11590 | 0,242 | 0,503 | 0,475 | APOE | 1 |

| | | | | | | | | |
|---|---|---|---|---|---|---|---|---|
| GO:1904029 | regulation of cyclin-dependent protein kinase activity | 1/38 | 84/11590 | 0,242 | 0,503 | 0,475 | HEXIM1 | 1 |
| GO:1990748 | cellular detoxification | 1/38 | 84/11590 | 0,242 | 0,503 | 0,475 | APOE | 1 |
| GO:0006165 | nucleoside diphosphate phosphorylation | 1/38 | 85/11590 | 0,244 | 0,504 | 0,475 | GALK1 | 1 |
| GO:0043535 | regulation of blood vessel endothelial cell migration | 1/38 | 85/11590 | 0,244 | 0,504 | 0,475 | APOE | 1 |
| GO:0051952 | regulation of amine transport | 1/38 | 85/11590 | 0,244 | 0,504 | 0,475 | SLC38A1 | 1 |
| GO:0010639 | negative regulation of organelle organization | 2/38 | 289/11590 | 0,245 | 0,504 | 0,475 | STN1/H3-3A | 2 |
| GO:0042176 | regulation of protein catabolic process | 2/38 | 289/11590 | 0,245 | 0,504 | 0,475 | UFL1/APOE | 2 |
| GO:0006644 | phospholipid metabolic process | 2/38 | 290/11590 | 0,246 | 0,505 | 0,476 | MTMR9/APOC1 | 2 |
| GO:0031058 | positive regulation of histone modification | 1/38 | 86/11590 | 0,247 | 0,505 | 0,476 | WBP2 | 1 |
| GO:0048259 | regulation of receptor-mediated endocytosis | 1/38 | 86/11590 | 0,247 | 0,505 | 0,476 | APOC1 | 1 |
| GO:0007044 | cell-substrate junction assembly | 1/38 | 87/11590 | 0,249 | 0,506 | 0,477 | STON1 | 1 |
| GO:0046939 | nucleotide phosphorylation | 1/38 | 87/11590 | 0,249 | 0,506 | 0,477 | GALK1 | 1 |
| GO:0007006 | mitochondrial membrane organization | 1/38 | 88/11590 | 0,252 | 0,506 | 0,477 | NMT1 | 1 |
| GO:0010769 | regulation of cell morphogenesis involved in differentiation | 1/38 | 88/11590 | 0,252 | 0,506 | 0,477 | STAU2 | 1 |
| GO:0014013 | regulation of gliogenesis | 1/38 | 88/11590 | 0,252 | 0,506 | 0,477 | UFL1 | 1 |
| GO:1901796 | regulation of signal transduction by p53 class mediator | 1/38 | 88/11590 | 0,252 | 0,506 | 0,477 | HEXIM1 | 1 |
| GO:0050727 | regulation of inflammatory response | 2/38 | 295/11590 | 0,252 | 0,506 | 0,477 | UFL1/APOE | 2 |
| GO:0006368 | transcription elongation by RNA polymerase II promoter | 1/38 | 89/11590 | 0,254 | 0,506 | 0,477 | HEXIM1 | 1 |
| GO:0008637 | apoptotic mitochondrial changes | 1/38 | 89/11590 | 0,254 | 0,506 | 0,477 | NMT1 | 1 |
| GO:0051224 | negative regulation of protein transport | 1/38 | 89/11590 | 0,254 | 0,506 | 0,477 | APOE | 1 |
| GO:0060840 | artery development | 1/38 | 89/11590 | 0,254 | 0,506 | 0,477 | APOE | 1 |
| GO:0042326 | negative regulation of phosphorylation | 2/38 | 297/11590 | 0,254 | 0,506 | 0,477 | APOE/HEXIM1 | 2 |
| GO:0033559 | unsaturated fatty acid metabolic process | 1/38 | 90/11590 | 0,257 | 0,506 | 0,477 | ACOX1 | 1 |
| GO:0048545 | response to steroid hormone | 2/38 | 299/11590 | 0,257 | 0,506 | 0,477 | UFL1/WBP2 | 2 |

| | | | | | | | | |
|---|---|---|---|---|---|---|---|---|
| GO:0031346 | positive regulation of cell projection organization | 2/38 | 300/11590 | 0,258 | 0,506 | 0,477 | STAU2/APOE | 2 |
| GO:0009135 | purine nucleoside diphosphate metabolic process | 1/38 | 91/11590 | 0,259 | 0,506 | 0,477 | GALK1 | 1 |
| GO:0009179 | purine ribonucleoside diphosphate metabolic process | 1/38 | 91/11590 | 0,259 | 0,506 | 0,477 | GALK1 | 1 |
| GO:0019080 | viral gene expression | 1/38 | 91/11590 | 0,259 | 0,506 | 0,477 | HEXIM1 | 1 |
| GO:0033865 | nucleoside bisphosphate metabolic process | 1/38 | 91/11590 | 0,259 | 0,506 | 0,477 | DCAKD | 1 |
| GO:0033875 | ribonucleoside bisphosphate metabolic process | 1/38 | 91/11590 | 0,259 | 0,506 | 0,477 | DCAKD | 1 |
| GO:0034032 | purine nucleoside bisphosphate metabolic process | 1/38 | 91/11590 | 0,259 | 0,506 | 0,477 | DCAKD | 1 |
| GO:0097237 | cellular response to toxic substance | 1/38 | 91/11590 | 0,259 | 0,506 | 0,477 | APOE | 1 |
| GO:1905954 | positive regulation of lipid localization | 1/38 | 91/11590 | 0,259 | 0,506 | 0,477 | APOE | 1 |
| GO:0015837 | amine transport | 1/38 | 92/11590 | 0,262 | 0,508 | 0,480 | SLC38A1 | 1 |
| GO:0006690 | icosanoid metabolic process | 1/38 | 93/11590 | 0,264 | 0,508 | 0,480 | ACOX1 | 1 |
| GO:0007173 | epidermal growth factor receptor signaling pathway | 1/38 | 93/11590 | 0,264 | 0,508 | 0,480 | EFEMP1 | 1 |
| GO:0019395 | fatty acid oxidation | 1/38 | 93/11590 | 0,264 | 0,508 | 0,480 | ACOX1 | 1 |
| GO:0034728 | nucleosome organization | 1/38 | 93/11590 | 0,264 | 0,508 | 0,480 | H3-3A | 1 |
| GO:0150115 | cell-substrate junction organization | 1/38 | 93/11590 | 0,264 | 0,508 | 0,480 | STON1 | 1 |
| GO:1904950 | negative regulation of establishment of protein localization | 1/38 | 93/11590 | 0,264 | 0,508 | 0,480 | APOE | 1 |
| GO:0035967 | cellular response to topologically incorrect protein | 1/38 | 94/11590 | 0,267 | 0,511 | 0,482 | UFL1 | 1 |
| GO:0061387 | regulation of extent of cell growth | 1/38 | 94/11590 | 0,267 | 0,511 | 0,482 | APOE | 1 |
| GO:0051702 | biological process involved in interaction with symbiont | 1/38 | 95/11590 | 0,269 | 0,515 | 0,485 | APOE | 1 |
| GO:0002062 | chondrocyte differentiation | 1/38 | 96/11590 | 0,271 | 0,516 | 0,487 | EFEMP1 | 1 |
| GO:0043200 | response to amino acid | 1/38 | 96/11590 | 0,271 | 0,516 | 0,487 | UFL1 | 1 |
| GO:0006869 | lipid transport | 2/38 | 312/11590 | 0,273 | 0,516 | 0,487 | APOE/APOC1 | 2 |
| GO:0019751 | polyol metabolic process | 1/38 | 97/11590 | 0,274 | 0,516 | 0,487 | GALK1 | 1 |

| | | | | | | | | |
|---|---|---|---|---|---|---|---|---|
| GO:0072655 | establishment of protein localization to mitochondrion | 1/38 | 97/11590 | 0,274 | 0,516 | 0,487 | NMT1 | 1 |
| GO:0031331 | positive regulation of cellular catabolic process | 2/38 | 313/11590 | 0,274 | 0,516 | 0,487 | UFL1/APOE | 2 |
| GO:0050767 | regulation of neurogenesis | 2/38 | 313/11590 | 0,274 | 0,516 | 0,487 | UFL1/STAU2 | 2 |
| GO:0071900 | regulation of protein serine/threonine kinase activity | 2/38 | 313/11590 | 0,274 | 0,516 | 0,487 | APOE/HEXIM1 | 2 |
| GO:0007338 | single fertilization | 1/38 | 98/11590 | 0,276 | 0,517 | 0,488 | H3-3A | 1 |
| GO:0034440 | lipid oxidation | 1/38 | 98/11590 | 0,276 | 0,517 | 0,488 | ACOX1 | 1 |
| GO:0071482 | cellular response to light stimulus | 1/38 | 98/11590 | 0,276 | 0,517 | 0,488 | NMT1 | 1 |
| GO:0001654 | eye development | 2/38 | 316/11590 | 0,278 | 0,519 | 0,489 | STAU2/EFEMP1 | 2 |
| GO:0007613 | memory | 1/38 | 99/11590 | 0,279 | 0,519 | 0,489 | APOE | 1 |
| GO:0009185 | ribonucleoside diphosphate metabolic process | 1/38 | 99/11590 | 0,279 | 0,519 | 0,489 | GALK1 | 1 |
| GO:0006090 | pyruvate metabolic process | 1/38 | 100/11590 | 0,281 | 0,520 | 0,491 | GALK1 | 1 |
| GO:0072329 | monocarboxylic acid catabolic process | 1/38 | 100/11590 | 0,281 | 0,520 | 0,491 | ACOX1 | 1 |
| GO:0090090 | negative regulation of canonical Wnt signaling pathway | 1/38 | 100/11590 | 0,281 | 0,520 | 0,491 | APOE | 1 |
| GO:0150063 | visual system development | 2/38 | 319/11590 | 0,281 | 0,520 | 0,491 | STAU2/EFEMP1 | 2 |
| GO:0001701 | in utero embryonic development | 2/38 | 321/11590 | 0,284 | 0,524 | 0,494 | SH3PXD2A/NMT1 | 2 |
| GO:0070585 | protein localization to mitochondrion | 1/38 | 102/11590 | 0,286 | 0,526 | 0,496 | NMT1 | 1 |
| GO:0048880 | sensory system development | 2/38 | 324/11590 | 0,287 | 0,529 | 0,498 | STAU2/EFEMP1 | 2 |
| GO:0035303 | regulation of dephosphorylation | 1/38 | 103/11590 | 0,288 | 0,529 | 0,499 | MTMR9 | 1 |
| GO:0043254 | regulation of protein-containing complex assembly | 2/38 | 325/11590 | 0,289 | 0,529 | 0,499 | APOE/H3-3A | 2 |
| GO:0007286 | spermatid development | 1/38 | 104/11590 | 0,290 | 0,530 | 0,500 | H3-3A | 1 |
| GO:0009581 | detection of external stimulus | 1/38 | 104/11590 | 0,290 | 0,530 | 0,500 | NMT1 | 1 |
| GO:0030099 | myeloid cell differentiation | 2/38 | 328/11590 | 0,292 | 0,531 | 0,501 | UFL1/SH3PXD2A | 2 |

| | | | | | | | | |
|---|---|---|---|---|---|---|---|---|
| GO:0098754 | detoxification | 1/38 | 105/11590 | 0,293 | 0,531 | 0,501 | APOE | 1 |
| GO:2000278 | regulation of DNA biosynthetic process | 1/38 | 105/11590 | 0,293 | 0,531 | 0,501 | STN1 | 1 |
| GO:0002244 | hematopoietic progenitor cell differentiation | 1/38 | 106/11590 | 0,295 | 0,532 | 0,501 | UFL1 | 1 |
| GO:0009582 | detection of abiotic stimulus | 1/38 | 106/11590 | 0,295 | 0,532 | 0,501 | NMT1 | 1 |
| GO:0038127 | ERBB signaling pathway | 1/38 | 106/11590 | 0,295 | 0,532 | 0,501 | EFEMP1 | 1 |
| GO:0048675 | axon extension | 1/38 | 106/11590 | 0,295 | 0,532 | 0,501 | APOE | 1 |
| GO:0050808 | synapse organization | 2/38 | 331/11590 | 0,296 | 0,532 | 0,502 | STAU2/APOE | 2 |
| GO:0010565 | regulation of cellular ketone metabolic process | 1/38 | 107/11590 | 0,297 | 0,534 | 0,503 | APOC1 | 1 |
| GO:0051051 | negative regulation of transport | 2/38 | 335/11590 | 0,301 | 0,537 | 0,507 | APOE/APOC1 | 2 |
| GO:0001101 | response to acid chemical | 1/38 | 109/11590 | 0,302 | 0,537 | 0,507 | UFL1 | 1 |
| GO:0032434 | regulation of proteasomal ubiquitin-dependent protein catabolic process | 1/38 | 109/11590 | 0,302 | 0,537 | 0,507 | UFL1 | 1 |
| GO:0048515 | spermatid differentiation | 1/38 | 109/11590 | 0,302 | 0,537 | 0,507 | H3-3A | 1 |
| GO:0006986 | response to unfolded protein | 1/38 | 110/11590 | 0,304 | 0,537 | 0,507 | UFL1 | 1 |
| GO:0009132 | nucleoside diphosphate metabolic process | 1/38 | 110/11590 | 0,304 | 0,537 | 0,507 | GALK1 | 1 |
| GO:0030216 | keratinocyte differentiation | 1/38 | 110/11590 | 0,304 | 0,537 | 0,507 | EVPL | 1 |
| GO:0030218 | erythrocyte differentiation | 1/38 | 110/11590 | 0,304 | 0,537 | 0,507 | UFL1 | 1 |
| GO:0043534 | blood vessel endothelial cell migration | 1/38 | 110/11590 | 0,304 | 0,537 | 0,507 | APOE | 1 |
| GO:0034329 | cell junction assembly | 2/38 | 339/11590 | 0,306 | 0,538 | 0,507 | STAU2/STON1 | 2 |
| GO:0007030 | Golgi organization | 1/38 | 111/11590 | 0,307 | 0,538 | 0,507 | COG2 | 1 |
| GO:0030177 | positive regulation of Wnt signaling pathway | 1/38 | 111/11590 | 0,307 | 0,538 | 0,507 | GPC5 | 1 |
| GO:0045089 | positive regulation of innate immune response | 1/38 | 111/11590 | 0,307 | 0,538 | 0,507 | HEXIM1 | 1 |
| GO:0006275 | regulation of DNA replication | 1/38 | 112/11590 | 0,309 | 0,540 | 0,509 | STN1 | 1 |
| GO:0051053 | negative regulation of DNA metabolic process | 1/38 | 112/11590 | 0,309 | 0,540 | 0,509 | STN1 | 1 |
| GO:0016055 | Wnt signaling pathway | 2/38 | 343/11590 | 0,311 | 0,541 | 0,510 | APOE/GPC5 | 2 |
| GO:0010508 | positive regulation of autophagy | 1/38 | 113/11590 | 0,311 | 0,541 | 0,510 | UFL1 | 1 |
| GO:0001558 | regulation of cell growth | 2/38 | 344/11590 | 0,312 | 0,541 | 0,510 | APOE/H3-3A | 2 |

| | | | | | | | | |
|---|---|---|---|---|---|---|---|---|
| GO:0198738 | cell-cell signaling by wnt | 2/38 | 344/11590 | 0,312 | 0,541 | 0,510 | APOE/GPC5 | 2 |
| GO:0006865 | amino acid transport | 1/38 | 115/11590 | 0,316 | 0,545 | 0,514 | SLC38A1 | 1 |
| GO:0006997 | nucleus organization | 1/38 | 115/11590 | 0,316 | 0,545 | 0,514 | H3-3A | 1 |
| GO:0007292 | female gamete generation | 1/38 | 115/11590 | 0,316 | 0,545 | 0,514 | H3-3A | 1 |
| GO:0010876 | lipid localization | 2/38 | 348/11590 | 0,317 | 0,546 | 0,515 | APOE/APOC1 | 2 |
| GO:0009150 | purine ribonucleotide metabolic process | 2/38 | 349/11590 | 0,318 | 0,547 | 0,516 | GALK1/DCAKD | 2 |
| GO:0010977 | negative regulation of neuron projection development | 1/38 | 117/11590 | 0,320 | 0,548 | 0,517 | APOE | 1 |
| GO:1903008 | organelle disassembly | 1/38 | 117/11590 | 0,320 | 0,548 | 0,517 | UFL1 | 1 |
| GO:0006979 | response to oxidative stress | 2/38 | 352/11590 | 0,322 | 0,548 | 0,517 | STAU2/APOE | 2 |
| GO:0030168 | platelet activation | 1/38 | 118/11590 | 0,323 | 0,548 | 0,517 | APOE | 1 |
| GO:0034101 | erythrocyte homeostasis | 1/38 | 118/11590 | 0,323 | 0,548 | 0,517 | UFL1 | 1 |
| GO:1903825 | organic acid transmembrane transport | 1/38 | 118/11590 | 0,323 | 0,548 | 0,517 | SLC38A1 | 1 |
| GO:1905039 | carboxylic acid transmembrane transport | 1/38 | 118/11590 | 0,323 | 0,548 | 0,517 | SLC38A1 | 1 |
| GO:0046165 | alcohol biosynthetic process | 1/38 | 119/11590 | 0,325 | 0,548 | 0,517 | APOE | 1 |
| GO:0046434 | organophosphate catabolic process | 1/38 | 119/11590 | 0,325 | 0,548 | 0,517 | APOC1 | 1 |
| GO:0048813 | dendrite morphogenesis | 1/38 | 119/11590 | 0,325 | 0,548 | 0,517 | STAU2 | 1 |
| GO:0061041 | regulation of wound healing | 1/38 | 119/11590 | 0,325 | 0,548 | 0,517 | APOE | 1 |
| GO:0001936 | regulation of endothelial cell proliferation | 1/38 | 120/11590 | 0,327 | 0,550 | 0,518 | APOE | 1 |
| GO:0072594 | establishment of protein localization to organelle | 2/38 | 357/11590 | 0,328 | 0,550 | 0,518 | WBP2/NMT1 | 2 |
| GO:0000723 | telomere maintenance | 1/38 | 121/11590 | 0,329 | 0,550 | 0,518 | STN1 | 1 |
| GO:0008277 | regulation of G protein-coupled receptor signaling pathway | 1/38 | 121/11590 | 0,329 | 0,550 | 0,518 | NMT1 | 1 |
| GO:0030178 | negative regulation of Wnt signaling pathway | 1/38 | 121/11590 | 0,329 | 0,550 | 0,518 | APOE | 1 |
| GO:1903531 | negative regulation of secretion by cell | 1/38 | 121/11590 | 0,329 | 0,550 | 0,518 | APOE | 1 |
| GO:0007283 | spermatogenesis | 2/38 | 360/11590 | 0,331 | 0,552 | 0,520 | H3-3A/ACOX1 | 2 |
| GO:0050728 | negative regulation of inflammatory response | 1/38 | 122/11590 | 0,332 | 0,552 | 0,520 | APOE | 1 |

| | | | | | | | | |
|---|---|---|---|---|---|---|---|---|
| GO:0062013 | positive regulation of small molecule metabolic process | 1/38 | 123/11590 | 0,334 | 0,553 | 0,522 | APOE | 1 |
| GO:0016032 | viral process | 2/38 | 362/11590 | 0,334 | 0,553 | 0,522 | APOE/HEXIM1 | 2 |
| GO:0009566 | fertilization | 1/38 | 124/11590 | 0,336 | 0,555 | 0,524 | H3-3A | 1 |
| GO:0050680 | negative regulation of epithelial cell proliferation | 1/38 | 124/11590 | 0,336 | 0,555 | 0,524 | APOE | 1 |
| GO:0009259 | ribonucleotide metabolic process | 2/38 | 365/11590 | 0,337 | 0,556 | 0,524 | GALK1/DCAKD | 2 |
| GO:0008037 | cell recognition | 1/38 | 125/11590 | 0,338 | 0,556 | 0,524 | VCAN | 1 |
| GO:0010821 | regulation of mitochondrion organization | 1/38 | 125/11590 | 0,338 | 0,556 | 0,524 | NMT1 | 1 |
| GO:0006633 | fatty acid biosynthetic process | 1/38 | 126/11590 | 0,340 | 0,558 | 0,526 | APOC1 | 1 |
| GO:0035264 | multicellular organism growth | 1/38 | 126/11590 | 0,340 | 0,558 | 0,526 | H3-3A | 1 |
| GO:0006163 | purine nucleotide metabolic process | 2/38 | 371/11590 | 0,345 | 0,560 | 0,528 | GALK1/DCAKD | 2 |
| GO:0010975 | regulation of neuron projection development | 2/38 | 371/11590 | 0,345 | 0,560 | 0,528 | STAU2/APOE | 2 |
| GO:0010970 | transport along microtubule | 1/38 | 128/11590 | 0,345 | 0,560 | 0,528 | STAU2 | 1 |
| GO:0035966 | response to topologically incorrect protein | 1/38 | 128/11590 | 0,345 | 0,560 | 0,528 | UFL1 | 1 |
| GO:0046488 | phosphatidylinositol metabolic process | 1/38 | 128/11590 | 0,345 | 0,560 | 0,528 | MTMR9 | 1 |
| GO:0051960 | regulation of nervous system development | 2/38 | 372/11590 | 0,346 | 0,560 | 0,528 | UFL1/STAU2 | 2 |
| GO:0008584 | male gonad development | 1/38 | 129/11590 | 0,347 | 0,560 | 0,528 | H3-3A | 1 |
| GO:0019693 | ribose phosphate metabolic process | 2/38 | 373/11590 | 0,347 | 0,560 | 0,528 | GALK1/DCAKD | 2 |
| GO:0048232 | male gamete generation | 2/38 | 373/11590 | 0,347 | 0,560 | 0,528 | H3-3A/ACOX1 | 2 |
| GO:0046546 | development of primary male sexual characteristics | 1/38 | 130/11590 | 0,349 | 0,562 | 0,530 | H3-3A | 1 |
| GO:0042060 | wound healing | 2/38 | 375/11590 | 0,349 | 0,562 | 0,530 | APOE/EVPL | 2 |
| GO:0007416 | synapse assembly | 1/38 | 131/11590 | 0,351 | 0,562 | 0,530 | STAU2 | 1 |
| GO:1902904 | negative regulation of supramolecular fiber organization | 1/38 | 131/11590 | 0,351 | 0,562 | 0,530 | APOE | 1 |
| GO:2000058 | regulation of ubiquitin-dependent protein catabolic process | 1/38 | 131/11590 | 0,351 | 0,562 | 0,530 | UFL1 | 1 |

| | | | | | | | | |
|---|---|---|---|---|---|---|---|---|
| GO:0043524 | negative regulation of neuron apoptotic process | 1/38 | 132/11590 | 0,353 | 0,563 | 0,531 | APOE | 1 |
| GO:0048660 | regulation of smooth muscle cell proliferation | 1/38 | 132/11590 | 0,353 | 0,563 | 0,531 | APOE | 1 |
| GO:0050806 | positive regulation of synaptic transmission | 1/38 | 132/11590 | 0,353 | 0,563 | 0,531 | APOE | 1 |
| GO:0016570 | histone modification | 2/38 | 381/11590 | 0,357 | 0,566 | 0,534 | UFL1/WBP2 | 2 |
| GO:0001935 | endothelial cell proliferation | 1/38 | 134/11590 | 0,358 | 0,566 | 0,534 | APOE | 1 |
| GO:0016052 | carbohydrate catabolic process | 1/38 | 134/11590 | 0,358 | 0,566 | 0,534 | GALK1 | 1 |
| GO:0030307 | positive regulation of cell growth | 1/38 | 135/11590 | 0,360 | 0,566 | 0,534 | H3-3A | 1 |
| GO:0035150 | regulation of tube size | 1/38 | 135/11590 | 0,360 | 0,566 | 0,534 | APOE | 1 |
| GO:0035296 | regulation of tube diameter | 1/38 | 135/11590 | 0,360 | 0,566 | 0,534 | APOE | 1 |
| GO:0043433 | negative regulation of DNA-binding transcription factor activity | 1/38 | 135/11590 | 0,360 | 0,566 | 0,534 | UFL1 | 1 |
| GO:0097746 | blood vessel diameter maintenance | 1/38 | 135/11590 | 0,360 | 0,566 | 0,534 | APOE | 1 |
| GO:0048659 | smooth muscle cell proliferation | 1/38 | 136/11590 | 0,362 | 0,568 | 0,536 | APOE | 1 |
| GO:0010498 | proteasomal protein catabolic process | 2/38 | 386/11590 | 0,363 | 0,569 | 0,536 | UFL1/APOE | 2 |
| GO:0043409 | negative regulation of MAPK cascade | 1/38 | 137/11590 | 0,364 | 0,570 | 0,537 | APOE | 1 |
| GO:0031503 | protein-containing complex localization | 1/38 | 138/11590 | 0,366 | 0,572 | 0,540 | STAU2 | 1 |
| GO:0051048 | negative regulation of secretion | 1/38 | 139/11590 | 0,368 | 0,574 | 0,541 | APOE | 1 |
| GO:0090316 | positive regulation of intracellular protein transport | 1/38 | 139/11590 | 0,368 | 0,574 | 0,541 | NMT1 | 1 |
| GO:0120031 | plasma membrane bounded cell projection assembly | 2/38 | 393/11590 | 0,371 | 0,577 | 0,544 | STAU2/STON1 | 2 |
| GO:0050792 | regulation of viral process | 1/38 | 141/11590 | 0,372 | 0,577 | 0,545 | HEXIM1 | 1 |
| GO:1904064 | positive regulation of cation transmembrane transport | 1/38 | 141/11590 | 0,372 | 0,577 | 0,545 | SLC38A1 | 1 |
| GO:0072521 | purine-containing compound metabolic process | 2/38 | 395/11590 | 0,373 | 0,578 | 0,545 | GALK1/DCAKD | 2 |
| GO:0009896 | positive regulation of catabolic process | 2/38 | 397/11590 | 0,376 | 0,578 | 0,546 | UFL1/APOE | 2 |
| GO:0022411 | cellular component disassembly | 2/38 | 397/11590 | 0,376 | 0,578 | 0,546 | UFL1/STON1 | 2 |
| GO:0006839 | mitochondrial transport | 1/38 | 143/11590 | 0,377 | 0,578 | 0,546 | NMT1 | 1 |
| GO:0031056 | regulation of histone modification | 1/38 | 143/11590 | 0,377 | 0,578 | 0,546 | WBP2 | 1 |

| | | | | | | | | |
|---|---|---|---|---|---|---|---|---|
| GO:0048469 | cell maturation | 1/38 | 143/11590 | 0,377 | 0,578 | 0,546 | H3-3A | 1 |
| GO:0002833 | positive regulation of response to biotic stimulus | 1/38 | 144/11590 | 0,379 | 0,580 | 0,547 | HEXIM1 | 1 |
| GO:1990138 | neuron projection extension | 1/38 | 144/11590 | 0,379 | 0,580 | 0,547 | APOE | 1 |
| GO:0030031 | cell projection assembly | 2/38 | 401/11590 | 0,381 | 0,581 | 0,548 | STAU2/STON1 | 2 |
| GO:0072331 | signal transduction by p53 class mediator | 1/38 | 145/11590 | 0,381 | 0,581 | 0,548 | HEXIM1 | 1 |
| GO:0006091 | generation of precursor metabolites and energy | 2/38 | 402/11590 | 0,382 | 0,581 | 0,548 | GALK1/ACOX1 | 2 |
| GO:0007264 | small GTPase mediated signal transduction | 2/38 | 402/11590 | 0,382 | 0,581 | 0,548 | APOE/ARHGEF12 | 2 |
| GO:1903034 | regulation of response to wounding | 1/38 | 146/11590 | 0,383 | 0,582 | 0,548 | APOE | 1 |
| GO:1903828 | negative regulation of protein localization | 1/38 | 147/11590 | 0,385 | 0,584 | 0,551 | APOE | 1 |
| GO:0002262 | myeloid cell homeostasis | 1/38 | 148/11590 | 0,387 | 0,585 | 0,552 | UFL1 | 1 |
| GO:0046661 | male sex differentiation | 1/38 | 148/11590 | 0,387 | 0,585 | 0,552 | H3-3A | 1 |
| GO:0016049 | cell growth | 2/38 | 407/11590 | 0,388 | 0,585 | 0,552 | APOE/H3-3A | 2 |
| GO:0016573 | histone acetylation | 1/38 | 150/11590 | 0,391 | 0,588 | 0,554 | WBP2 | 1 |
| GO:0034249 | negative regulation of cellular amide metabolic process | 1/38 | 150/11590 | 0,391 | 0,588 | 0,554 | APOE | 1 |
| GO:0099111 | microtubule-based transport | 1/38 | 150/11590 | 0,391 | 0,588 | 0,554 | STAU2 | 1 |
| GO:0006694 | steroid biosynthetic process | 1/38 | 151/11590 | 0,393 | 0,589 | 0,555 | APOE | 1 |
| GO:0010594 | regulation of endothelial cell migration | 1/38 | 151/11590 | 0,393 | 0,589 | 0,555 | APOE | 1 |
| GO:0051223 | regulation of protein transport | 2/38 | 412/11590 | 0,393 | 0,589 | 0,555 | APOE/NMT1 | 2 |
| GO:0099504 | synaptic vesicle cycle | 1/38 | 152/11590 | 0,395 | 0,589 | 0,556 | STON1 | 1 |
| GO:1902905 | positive regulation of supramolecular fiber organization | 1/38 | 152/11590 | 0,395 | 0,589 | 0,556 | APOE | 1 |
| GO:0034767 | positive regulation of ion transmembrane transport | 1/38 | 153/11590 | 0,397 | 0,591 | 0,558 | SLC38A1 | 1 |
| GO:0065004 | protein-DNA complex assembly | 1/38 | 154/11590 | 0,399 | 0,593 | 0,560 | H3-3A | 1 |
| GO:0018393 | internal peptidyl-lysine acetylation | 1/38 | 155/11590 | 0,401 | 0,594 | 0,560 | WBP2 | 1 |

| | | | | | | | | |
|---|---|---|---|---|---|---|---|---|
| GO:0031345 | negative regulation of cell projection organization | 1/38 | 155/11590 | 0,401 | 0,594 | 0,560 | APOE | 1 |
| GO:0051099 | positive regulation of binding | 1/38 | 155/11590 | 0,401 | 0,594 | 0,560 | APOE | 1 |
| GO:0006475 | internal protein amino acid acetylation | 1/38 | 157/11590 | 0,405 | 0,597 | 0,563 | WBP2 | 1 |
| GO:0071478 | cellular response to radiation | 1/38 | 157/11590 | 0,405 | 0,597 | 0,563 | NMT1 | 1 |
| GO:0008361 | regulation of cell size | 1/38 | 158/11590 | 0,407 | 0,597 | 0,563 | APOE | 1 |
| GO:0030705 | cytoskeleton-dependent intracellular transport | 1/38 | 158/11590 | 0,407 | 0,597 | 0,563 | STAU2 | 1 |
| GO:0120032 | regulation of plasma membrane bounded cell projection assembly | 1/38 | 158/11590 | 0,407 | 0,597 | 0,563 | STAU2 | 1 |
| GO:0006914 | autophagy | 2/38 | 424/11590 | 0,407 | 0,597 | 0,563 | UFL1/MTMR9 | 2 |
| GO:0061919 | process utilizing autophagic mechanism | 2/38 | 424/11590 | 0,407 | 0,597 | 0,563 | UFL1/MTMR9 | 2 |
| GO:0046578 | regulation of Ras protein signal transduction | 1/38 | 159/11590 | 0,409 | 0,597 | 0,563 | APOE | 1 |
| GO:0060491 | regulation of cell projection assembly | 1/38 | 159/11590 | 0,409 | 0,597 | 0,563 | STAU2 | 1 |
| GO:0009913 | epidermal cell differentiation | 1/38 | 160/11590 | 0,411 | 0,598 | 0,564 | EVPL | 1 |
| GO:0031396 | regulation of protein ubiquitination | 1/38 | 160/11590 | 0,411 | 0,598 | 0,564 | UFL1 | 1 |
| GO:1901888 | regulation of cell junction assembly | 1/38 | 161/11590 | 0,413 | 0,600 | 0,566 | STAU2 | 1 |
| GO:0071897 | DNA biosynthetic process | 1/38 | 162/11590 | 0,415 | 0,602 | 0,568 | STN1 | 1 |
| GO:0018394 | peptidyl-lysine acetylation | 1/38 | 163/11590 | 0,417 | 0,603 | 0,569 | WBP2 | 1 |
| GO:0070201 | regulation of establishment of protein localization | 2/38 | 433/11590 | 0,418 | 0,603 | 0,569 | APOE/NMT1 | 2 |
| GO:0043405 | regulation of MAP kinase activity | 1/38 | 164/11590 | 0,419 | 0,603 | 0,569 | APOE | 1 |
| GO:0045732 | positive regulation of protein catabolic process | 1/38 | 164/11590 | 0,419 | 0,603 | 0,569 | APOE | 1 |
| GO:0070085 | glycosylation | 1/38 | 164/11590 | 0,419 | 0,603 | 0,569 | COG2 | 1 |
| GO:0060284 | regulation of cell development | 2/38 | 434/11590 | 0,419 | 0,603 | 0,569 | UFL1/STAU2 | 2 |
| GO:0098739 | import across plasma membrane | 1/38 | 165/11590 | 0,421 | 0,603 | 0,569 | SLC38A1 | 1 |
| GO:1901605 | alpha-amino acid metabolic process | 1/38 | 165/11590 | 0,421 | 0,603 | 0,569 | SLC38A1 | 1 |
| GO:0007601 | visual perception | 1/38 | 166/11590 | 0,423 | 0,605 | 0,571 | EFEMP1 | 1 |
| GO:0006403 | RNA localization | 1/38 | 167/11590 | 0,424 | 0,606 | 0,572 | STAU2 | 1 |
| GO:0072330 | monocarboxylic acid biosynthetic process | 1/38 | 167/11590 | 0,424 | 0,606 | 0,572 | APOC1 | 1 |

| | | | | | | | | |
|---|---|---|---|---|---|---|---|---|
| GO:0050953 | sensory perception of light stimulus | 1/38 | 168/11590 | 0,426 | 0,608 | 0,574 | EFEMP1 | 1 |
| GO:0007626 | locomotory behavior | 1/38 | 169/11590 | 0,428 | 0,609 | 0,574 | APOE | 1 |
| GO:0099003 | vesicle-mediated transport in synapse | 1/38 | 169/11590 | 0,428 | 0,609 | 0,574 | STON1 | 1 |
| GO:1905114 | cell surface receptor signaling pathway involved in cell-cell signaling | 2/38 | 443/11590 | 0,429 | 0,609 | 0,575 | APOE/GPC5 | 2 |
| GO:0000302 | response to reactive oxygen species | 1/38 | 170/11590 | 0,430 | 0,609 | 0,575 | APOE | 1 |
| GO:0043393 | regulation of protein binding | 1/38 | 170/11590 | 0,430 | 0,609 | 0,575 | APOE | 1 |
| GO:0051216 | cartilage development | 1/38 | 172/11590 | 0,434 | 0,614 | 0,579 | EFEMP1 | 1 |
| GO:0046034 | ATP metabolic process | 1/38 | 173/11590 | 0,436 | 0,615 | 0,580 | GALK1 | 1 |
| GO:0001501 | skeletal system development | 2/38 | 450/11590 | 0,437 | 0,616 | 0,581 | VCAN/EFEMP1 | 2 |
| GO:0032388 | positive regulation of intracellular transport | 1/38 | 174/11590 | 0,438 | 0,616 | 0,581 | NMT1 | 1 |
| GO:0009152 | purine ribonucleotide biosynthetic process | 1/38 | 175/11590 | 0,440 | 0,618 | 0,583 | DCAKD | 1 |
| GO:0007423 | sensory organ development | 2/38 | 453/11590 | 0,441 | 0,619 | 0,584 | STAU2/EFEMP1 | 2 |
| GO:0051276 | chromosome organization | 2/38 | 455/11590 | 0,443 | 0,621 | 0,585 | STN1/H3-3A | 2 |
| GO:0050821 | protein stabilization | 1/38 | 177/11590 | 0,443 | 0,621 | 0,585 | MTMR9 | 1 |
| GO:0050866 | negative regulation of cell activation | 1/38 | 178/11590 | 0,445 | 0,621 | 0,586 | APOE | 1 |
| GO:1901654 | response to ketone | 1/38 | 178/11590 | 0,445 | 0,621 | 0,586 | WBP2 | 1 |
| GO:0006836 | neurotransmitter transport | 1/38 | 179/11590 | 0,447 | 0,622 | 0,587 | SLC38A1 | 1 |
| GO:0070374 | positive regulation of ERK1 and ERK2 cascade | 1/38 | 179/11590 | 0,447 | 0,622 | 0,587 | APOE | 1 |
| GO:0009117 | nucleotide metabolic process | 2/38 | 461/11590 | 0,450 | 0,624 | 0,588 | GALK1/DCAKD | 2 |
| GO:0048667 | cell morphogenesis involved in neuron differentiation | 2/38 | 461/11590 | 0,450 | 0,624 | 0,588 | STAU2/APOE | 2 |
| GO:0002573 | myeloid leukocyte differentiation | 1/38 | 181/11590 | 0,451 | 0,624 | 0,588 | SH3PXD2A | 1 |
| GO:1901215 | negative regulation of neuron death | 1/38 | 181/11590 | 0,451 | 0,624 | 0,588 | APOE | 1 |
| GO:0006325 | chromatin organization | 2/38 | 466/11590 | 0,455 | 0,629 | 0,594 | WBP2/H3-3A | 2 |
| GO:0006753 | nucleoside phosphate metabolic process | 2/38 | 467/11590 | 0,456 | 0,629 | 0,594 | GALK1/DCAKD | 2 |

| | | | | | | | | |
|---|---|---|---|---|---|---|---|---|
| GO:0001505 | regulation of neurotransmitter levels | 1/38 | 185/11590 | 0,458 | 0,629 | 0,594 | SLC38A1 | 1 |
| GO:0006473 | protein acetylation | 1/38 | 185/11590 | 0,458 | 0,629 | 0,594 | WBP2 | 1 |
| GO:0071824 | protein-DNA complex subunit organization | 1/38 | 185/11590 | 0,458 | 0,629 | 0,594 | H3-3A | 1 |
| GO:0009260 | ribonucleotide biosynthetic process | 1/38 | 186/11590 | 0,460 | 0,630 | 0,594 | DCAKD | 1 |
| GO:0051651 | maintenance of location in cell | 1/38 | 186/11590 | 0,460 | 0,630 | 0,594 | APOE | 1 |
| GO:0033002 | muscle cell proliferation | 1/38 | 187/11590 | 0,462 | 0,632 | 0,596 | APOE | 1 |
| GO:0006814 | sodium ion transport | 1/38 | 189/11590 | 0,465 | 0,635 | 0,599 | SLC38A1 | 1 |
| GO:0043523 | regulation of neuron apoptotic process | 1/38 | 189/11590 | 0,465 | 0,635 | 0,599 | APOE | 1 |
| GO:0048588 | developmental cell growth | 1/38 | 191/11590 | 0,469 | 0,639 | 0,602 | APOE | 1 |
| GO:0009205 | purine ribonucleoside triphosphate metabolic process | 1/38 | 192/11590 | 0,470 | 0,639 | 0,602 | GALK1 | 1 |
| GO:0006164 | purine nucleotide biosynthetic process | 1/38 | 193/11590 | 0,472 | 0,639 | 0,602 | DCAKD | 1 |
| GO:0046390 | ribose phosphate biosynthetic process | 1/38 | 193/11590 | 0,472 | 0,639 | 0,602 | DCAKD | 1 |
| GO:0046395 | carboxylic acid catabolic process | 1/38 | 193/11590 | 0,472 | 0,639 | 0,602 | ACOX1 | 1 |
| GO:0009636 | response to toxic substance | 1/38 | 194/11590 | 0,474 | 0,639 | 0,602 | APOE | 1 |
| GO:0019318 | hexose metabolic process | 1/38 | 194/11590 | 0,474 | 0,639 | 0,602 | GALK1 | 1 |
| GO:0033157 | regulation of intracellular protein transport | 1/38 | 194/11590 | 0,474 | 0,639 | 0,602 | NMT1 | 1 |
| GO:0090150 | establishment of protein localization to membrane | 1/38 | 195/11590 | 0,476 | 0,639 | 0,602 | NMT1 | 1 |
| GO:0009144 | purine nucleoside triphosphate metabolic process | 1/38 | 196/11590 | 0,478 | 0,639 | 0,602 | GALK1 | 1 |
| GO:0060560 | developmental growth involved in morphogenesis | 1/38 | 196/11590 | 0,478 | 0,639 | 0,602 | APOE | 1 |
| GO:0008406 | gonad development | 1/38 | 197/11590 | 0,479 | 0,639 | 0,602 | H3-3A | 1 |
| GO:0009199 | ribonucleoside triphosphate metabolic process | 1/38 | 197/11590 | 0,479 | 0,639 | 0,602 | GALK1 | 1 |
| GO:0016054 | organic acid catabolic process | 1/38 | 197/11590 | 0,479 | 0,639 | 0,602 | ACOX1 | 1 |
| GO:0071695 | anatomical structure maturation | 1/38 | 197/11590 | 0,479 | 0,639 | 0,602 | H3-3A | 1 |
| GO:0071407 | cellular response to organic cyclic compound | 2/38 | 489/11590 | 0,481 | 0,639 | 0,602 | UFL1/WBP2 | 2 |
| GO:0034764 | positive regulation of transmembrane transport | 1/38 | 198/11590 | 0,481 | 0,639 | 0,602 | SLC38A1 | 1 |

| | | | | | | | | |
|---|---|---|---|---|---|---|---|---|
| GO:0043122 | regulation of I-kappaB kinase/NF-kappaB signaling | 1/38 | 198/11590 | 0,481 | 0,639 | 0,602 | UFL1 | 1 |
| GO:0043542 | endothelial cell migration | 1/38 | 198/11590 | 0,481 | 0,639 | 0,602 | APOE | 1 |
| GO:1903320 | regulation of protein modification by small protein conjugation or removal | 1/38 | 198/11590 | 0,481 | 0,639 | 0,602 | UFL1 | 1 |
| GO:0031348 | negative regulation of defense response | 1/38 | 199/11590 | 0,483 | 0,639 | 0,603 | APOE | 1 |
| GO:0072522 | purine-containing compound biosynthetic process | 1/38 | 199/11590 | 0,483 | 0,639 | 0,603 | DCAKD | 1 |
| GO:0007596 | blood coagulation | 1/38 | 200/11590 | 0,484 | 0,641 | 0,604 | APOE | 1 |
| GO:0009611 | response to wounding | 2/38 | 493/11590 | 0,485 | 0,641 | 0,604 | APOE/EVPL | 2 |
| GO:0045137 | development of primary sexual characteristics | 1/38 | 201/11590 | 0,486 | 0,641 | 0,605 | H3-3A | 1 |
| GO:0098656 | anion transmembrane transport | 1/38 | 202/11590 | 0,488 | 0,643 | 0,606 | SLC38A1 | 1 |
| GO:0034976 | response to endoplasmic reticulum stress | 1/38 | 203/11590 | 0,490 | 0,643 | 0,607 | UFL1 | 1 |
| GO:0050817 | coagulation | 1/38 | 203/11590 | 0,490 | 0,643 | 0,607 | APOE | 1 |
| GO:0048863 | stem cell differentiation | 1/38 | 204/11590 | 0,491 | 0,645 | 0,608 | UFL1 | 1 |
| GO:0007599 | hemostasis | 1/38 | 205/11590 | 0,493 | 0,646 | 0,609 | APOE | 1 |
| GO:0005996 | monosaccharide metabolic process | 1/38 | 206/11590 | 0,495 | 0,646 | 0,610 | GALK1 | 1 |
| GO:0007160 | cell-matrix adhesion | 1/38 | 206/11590 | 0,495 | 0,646 | 0,610 | STON1 | 1 |
| GO:0010632 | regulation of epithelial cell migration | 1/38 | 207/11590 | 0,496 | 0,648 | 0,611 | APOE | 1 |
| GO:0009141 | nucleoside triphosphate metabolic process | 1/38 | 209/11590 | 0,500 | 0,651 | 0,614 | GALK1 | 1 |
| GO:0050708 | regulation of protein secretion | 1/38 | 210/11590 | 0,501 | 0,651 | 0,614 | APOE | 1 |
| GO:0098657 | import into cell | 1/38 | 210/11590 | 0,501 | 0,651 | 0,614 | SLC38A1 | 1 |
| GO:1901617 | organic hydroxy compound biosynthetic process | 1/38 | 210/11590 | 0,501 | 0,651 | 0,614 | APOE | 1 |
| GO:0045927 | positive regulation of growth | 1/38 | 214/11590 | 0,508 | 0,659 | 0,621 | H3-3A | 1 |
| GO:0043588 | skin development | 1/38 | 217/11590 | 0,513 | 0,663 | 0,625 | EVPL | 1 |
| GO:0051606 | detection of stimulus | 1/38 | 217/11590 | 0,513 | 0,663 | 0,625 | NMT1 | 1 |
| GO:0046942 | carboxylic acid transport | 1/38 | 220/11590 | 0,518 | 0,669 | 0,631 | SLC38A1 | 1 |
| GO:0051402 | neuron apoptotic process | 1/38 | 223/11590 | 0,523 | 0,674 | 0,636 | APOE | 1 |
| GO:0007249 | I-kappaB kinase/NF-kappaB signaling | 1/38 | 224/11590 | 0,524 | 0,675 | 0,637 | UFL1 | 1 |

| | | | | | | | | |
|---|---|---|---|---|---|---|---|---|
| GO:0048562 | embryonic organ morphogenesis | 1/38 | 228/11590 | 0,531 | 0,682 | 0,644 | EFEMP1 | 1 |
| GO:0048193 | Golgi vesicle transport | 1/38 | 229/11590 | 0,532 | 0,683 | 0,645 | COG2 | 1 |
| GO:0006260 | DNA replication | 1/38 | 231/11590 | 0,535 | 0,685 | 0,646 | STN1 | 1 |
| GO:0006520 | cellular amino acid metabolic process | 1/38 | 231/11590 | 0,535 | 0,685 | 0,646 | SLC38A1 | 1 |
| GO:0034599 | cellular response to oxidative stress | 1/38 | 231/11590 | 0,535 | 0,685 | 0,646 | STAU2 | 1 |
| GO:0007611 | learning or memory | 1/38 | 233/11590 | 0,538 | 0,686 | 0,647 | APOE | 1 |
| GO:0009165 | nucleotide biosynthetic process | 1/38 | 233/11590 | 0,538 | 0,686 | 0,647 | DCAKD | 1 |
| GO:0061448 | connective tissue development | 1/38 | 233/11590 | 0,538 | 0,686 | 0,647 | EFEMP1 | 1 |
| GO:1901293 | nucleoside phosphate biosynthetic process | 1/38 | 234/11590 | 0,540 | 0,687 | 0,648 | DCAKD | 1 |
| GO:0006874 | cellular calcium ion homeostasis | 1/38 | 235/11590 | 0,541 | 0,688 | 0,649 | APOE | 1 |
| GO:0031349 | positive regulation of defense response | 1/38 | 237/11590 | 0,545 | 0,691 | 0,652 | HEXIM1 | 1 |
| GO:0016236 | macroautophagy | 1/38 | 238/11590 | 0,546 | 0,692 | 0,653 | UFL1 | 1 |
| GO:0007548 | sex differentiation | 1/38 | 239/11590 | 0,548 | 0,692 | 0,653 | H3-3A | 1 |
| GO:0021700 | developmental maturation | 1/38 | 239/11590 | 0,548 | 0,692 | 0,653 | H3-3A | 1 |
| GO:0043270 | positive regulation of ion transport | 1/38 | 243/11590 | 0,554 | 0,698 | 0,658 | SLC38A1 | 1 |
| GO:0070372 | regulation of ERK1 and ERK2 cascade | 1/38 | 243/11590 | 0,554 | 0,698 | 0,658 | APOE | 1 |
| GO:0044262 | cellular carbohydrate metabolic process | 1/38 | 244/11590 | 0,555 | 0,699 | 0,659 | GALK1 | 1 |
| GO:0048872 | homeostasis of number of cells | 1/38 | 245/11590 | 0,557 | 0,700 | 0,660 | UFL1 | 1 |
| GO:0030336 | negative regulation of cell migration | 1/38 | 247/11590 | 0,560 | 0,703 | 0,663 | APOE | 1 |
| GO:0022604 | regulation of cell morphogenesis | 1/38 | 252/11590 | 0,567 | 0,710 | 0,670 | STAU2 | 1 |
| GO:0051054 | positive regulation of DNA metabolic process | 1/38 | 252/11590 | 0,567 | 0,710 | 0,670 | STN1 | 1 |
| GO:0055074 | calcium ion homeostasis | 1/38 | 256/11590 | 0,573 | 0,717 | 0,676 | APOE | 1 |
| GO:0007018 | microtubule-based movement | 1/38 | 257/11590 | 0,574 | 0,717 | 0,676 | STAU2 | 1 |
| GO:0044403 | biological process involved in symbiotic interaction | 1/38 | 257/11590 | 0,574 | 0,717 | 0,676 | APOE | 1 |
| GO:0009416 | response to light stimulus | 1/38 | 259/11590 | 0,577 | 0,717 | 0,677 | NMT1 | 1 |
| GO:0051222 | positive regulation of protein transport | 1/38 | 259/11590 | 0,577 | 0,717 | 0,677 | NMT1 | 1 |
| GO:2000146 | negative regulation of cell motility | 1/38 | 259/11590 | 0,577 | 0,717 | 0,677 | APOE | 1 |
| GO:0048608 | reproductive structure development | 1/38 | 260/11590 | 0,578 | 0,718 | 0,677 | H3-3A | 1 |

| | | | | | | | | |
|---|---|---|---|---|---|---|---|---|
| GO:0050890 | cognition | 1/38 | 262/11590 | 0,581 | 0,719 | 0,678 | APOE | 1 |
| GO:0061458 | reproductive system development | 1/38 | 262/11590 | 0,581 | 0,719 | 0,678 | H3-3A | 1 |
| GO:0072503 | cellular divalent inorganic cation homeostasis | 1/38 | 262/11590 | 0,581 | 0,719 | 0,678 | APOE | 1 |
| GO:0008544 | epidermis development | 1/38 | 263/11590 | 0,583 | 0,720 | 0,679 | EVPL | 1 |
| GO:0031647 | regulation of protein stability | 1/38 | 264/11590 | 0,584 | 0,720 | 0,679 | MTMR9 | 1 |
| GO:0070371 | ERK1 and ERK2 cascade | 1/38 | 264/11590 | 0,584 | 0,720 | 0,679 | APOE | 1 |
| GO:0042063 | gliogenesis | 1/38 | 266/11590 | 0,587 | 0,722 | 0,681 | UFL1 | 1 |
| GO:0016311 | dephosphorylation | 1/38 | 269/11590 | 0,591 | 0,726 | 0,684 | MTMR9 | 1 |
| GO:0019932 | second-messenger-mediated signaling | 1/38 | 269/11590 | 0,591 | 0,726 | 0,684 | APOE | 1 |
| GO:0048638 | regulation of developmental growth | 1/38 | 271/11590 | 0,594 | 0,727 | 0,685 | APOE | 1 |
| GO:0010631 | epithelial cell migration | 1/38 | 272/11590 | 0,595 | 0,727 | 0,685 | APOE | 1 |
| GO:0019058 | viral life cycle | 1/38 | 272/11590 | 0,595 | 0,727 | 0,685 | APOE | 1 |
| GO:1904951 | positive regulation of establishment of protein localization | 1/38 | 272/11590 | 0,595 | 0,727 | 0,685 | NMT1 | 1 |
| GO:0002253 | activation of immune response | 1/38 | 273/11590 | 0,596 | 0,727 | 0,685 | HEXIM1 | 1 |
| GO:0051235 | maintenance of location | 1/38 | 274/11590 | 0,598 | 0,727 | 0,685 | APOE | 1 |
| GO:0051346 | negative regulation of hydrolase activity | 1/38 | 274/11590 | 0,598 | 0,727 | 0,685 | APOC1 | 1 |
| GO:0090132 | epithelium migration | 1/38 | 274/11590 | 0,598 | 0,727 | 0,685 | APOE | 1 |
| GO:0015711 | organic anion transport | 1/38 | 276/11590 | 0,600 | 0,729 | 0,688 | SLC38A1 | 1 |
| GO:1901214 | regulation of neuron death | 1/38 | 278/11590 | 0,603 | 0,731 | 0,690 | APOE | 1 |
| GO:0032386 | regulation of intracellular transport | 1/38 | 279/11590 | 0,604 | 0,731 | 0,690 | NMT1 | 1 |
| GO:0090130 | tissue migration | 1/38 | 279/11590 | 0,604 | 0,731 | 0,690 | APOE | 1 |
| GO:0062197 | cellular response to chemical stress | 1/38 | 282/11590 | 0,608 | 0,735 | 0,693 | STAU2 | 1 |
| GO:0009306 | protein secretion | 1/38 | 283/11590 | 0,610 | 0,735 | 0,693 | APOE | 1 |
| GO:0043010 | camera-type eye development | 1/38 | 283/11590 | 0,610 | 0,735 | 0,693 | EFEMP1 | 1 |
| GO:0035592 | establishment of protein localization to extracellular region | 1/38 | 284/11590 | 0,611 | 0,736 | 0,694 | APOE | 1 |
| GO:0071214 | cellular response to abiotic stimulus | 1/38 | 287/11590 | 0,615 | 0,739 | 0,696 | NMT1 | 1 |
| GO:0104004 | cellular response to environmental stimulus | 1/38 | 287/11590 | 0,615 | 0,739 | 0,696 | NMT1 | 1 |

| | | | | | | | | |
|---|---|---|---|---|---|---|---|---|
| GO:0040013 | negative regulation of locomotion | 1/38 | 288/11590 | 0,616 | 0,739 | 0,697 | APOE | 1 |
| GO:0071692 | protein localization to extracellular region | 1/38 | 289/11590 | 0,618 | 0,740 | 0,698 | APOE | 1 |
| GO:0072507 | divalent inorganic cation homeostasis | 1/38 | 290/11590 | 0,619 | 0,740 | 0,698 | APOE | 1 |
| GO:0032956 | regulation of actin cytoskeleton organization | 1/38 | 291/11590 | 0,620 | 0,741 | 0,699 | STAU2 | 1 |
| GO:1904062 | regulation of cation transmembrane transport | 1/38 | 293/11590 | 0,623 | 0,743 | 0,701 | SLC38A1 | 1 |
| GO:0032535 | regulation of cellular component size | 1/38 | 297/11590 | 0,628 | 0,748 | 0,706 | APOE | 1 |
| GO:0045862 | positive regulation of proteolysis | 1/38 | 303/11590 | 0,635 | 0,756 | 0,713 | APOE | 1 |
| GO:0042692 | muscle cell differentiation | 1/38 | 307/11590 | 0,640 | 0,760 | 0,717 | H3-3A | 1 |
| GO:1902903 | regulation of supramolecular fiber organization | 1/38 | 307/11590 | 0,640 | 0,760 | 0,717 | APOE | 1 |
| GO:0050678 | regulation of epithelial cell proliferation | 1/38 | 309/11590 | 0,642 | 0,762 | 0,719 | APOE | 1 |
| GO:0045786 | negative regulation of cell cycle | 1/38 | 316/11590 | 0,651 | 0,771 | 0,727 | HEXIM1 | 1 |
| GO:0051098 | regulation of binding | 1/38 | 317/11590 | 0,652 | 0,771 | 0,727 | APOE | 1 |
| GO:0070997 | neuron death | 1/38 | 317/11590 | 0,652 | 0,771 | 0,727 | APOE | 1 |
| GO:0031589 | cell-substrate adhesion | 1/38 | 318/11590 | 0,653 | 0,771 | 0,727 | STON1 | 1 |
| GO:0018108 | peptidyl-tyrosine phosphorylation | 1/38 | 325/11590 | 0,661 | 0,780 | 0,735 | EFEMP1 | 1 |
| GO:0018212 | peptidyl-tyrosine modification | 1/38 | 327/11590 | 0,664 | 0,781 | 0,736 | EFEMP1 | 1 |
| GO:0032970 | regulation of actin filament-based process | 1/38 | 327/11590 | 0,664 | 0,781 | 0,736 | STAU2 | 1 |
| GO:0018205 | peptidyl-lysine modification | 1/38 | 332/11590 | 0,669 | 0,786 | 0,741 | WBP2 | 1 |
| GO:0006875 | cellular metal ion homeostasis | 1/38 | 333/11590 | 0,670 | 0,786 | 0,741 | APOE | 1 |
| GO:0032102 | negative regulation of response to external stimulus | 1/38 | 333/11590 | 0,670 | 0,786 | 0,741 | APOE | 1 |
| GO:0043161 | proteasome-mediated ubiquitin-dependent protein catabolic process | 1/38 | 335/11590 | 0,673 | 0,787 | 0,743 | UFL1 | 1 |
| GO:0050878 | regulation of body fluid levels | 1/38 | 341/11590 | 0,679 | 0,794 | 0,749 | APOE | 1 |
| GO:0051090 | regulation of DNA-binding transcription factor activity | 1/38 | 357/11590 | 0,696 | 0,813 | 0,767 | UFL1 | 1 |
| GO:0034248 | regulation of cellular amide metabolic process | 1/38 | 358/11590 | 0,697 | 0,813 | 0,767 | APOE | 1 |
| GO:0048568 | embryonic organ development | 1/38 | 361/11590 | 0,700 | 0,816 | 0,769 | EFEMP1 | 1 |
| GO:0007409 | axonogenesis | 1/38 | 365/11590 | 0,704 | 0,820 | 0,773 | APOE | 1 |

| | | | | | | | | |
|---|---|---|---|---|---|---|---|---|
| GO:0050673 | epithelial cell proliferation | 1/38 | 369/11590 | 0,708 | 0,823 | 0,776 | APOE | 1 |
| GO:0009314 | response to radiation | 1/38 | 371/11590 | 0,710 | 0,825 | 0,778 | NMT1 | 1 |
| GO:0032103 | positive regulation of response to external stimulus | 1/38 | 374/11590 | 0,713 | 0,827 | 0,780 | HEXIM1 | 1 |
| GO:0001667 | ameboidal-type cell migration | 1/38 | 379/11590 | 0,718 | 0,832 | 0,784 | APOE | 1 |
| GO:0034765 | regulation of ion transmembrane transport | 1/38 | 382/11590 | 0,721 | 0,834 | 0,786 | SLC38A1 | 1 |
| GO:0006820 | anion transport | 1/38 | 386/11590 | 0,725 | 0,837 | 0,790 | SLC38A1 | 1 |
| GO:1903829 | positive regulation of protein localization | 1/38 | 391/11590 | 0,729 | 0,842 | 0,794 | NMT1 | 1 |
| GO:0030003 | cellular cation homeostasis | 1/38 | 392/11590 | 0,730 | 0,842 | 0,794 | APOE | 1 |
| GO:0007005 | mitochondrion organization | 1/38 | 394/11590 | 0,732 | 0,843 | 0,795 | NMT1 | 1 |
| GO:1901361 | organic cyclic compound catabolic process | 1/38 | 397/11590 | 0,735 | 0,845 | 0,797 | APOE | 1 |
| GO:0031667 | response to nutrient levels | 1/38 | 403/11590 | 0,740 | 0,850 | 0,802 | APOE | 1 |
| GO:0061564 | axon development | 1/38 | 404/11590 | 0,741 | 0,850 | 0,802 | APOE | 1 |
| GO:0006873 | cellular ion homeostasis | 1/38 | 406/11590 | 0,743 | 0,851 | 0,803 | APOE | 1 |
| GO:0044089 | positive regulation of cellular component biogenesis | 1/38 | 407/11590 | 0,744 | 0,851 | 0,803 | STAU2 | 1 |
| GO:0043410 | positive regulation of MAPK cascade | 1/38 | 413/11590 | 0,749 | 0,856 | 0,807 | APOE | 1 |
| GO:0010256 | endomembrane system organization | 1/38 | 414/11590 | 0,750 | 0,856 | 0,807 | COG2 | 1 |
| GO:1902532 | negative regulation of intracellular signal transduction | 1/38 | 415/11590 | 0,750 | 0,856 | 0,807 | APOE | 1 |
| GO:0055065 | metal ion homeostasis | 1/38 | 420/11590 | 0,755 | 0,860 | 0,811 | APOE | 1 |
| GO:0010638 | positive regulation of organelle organization | 1/38 | 423/11590 | 0,757 | 0,862 | 0,813 | NMT1 | 1 |
| GO:0090066 | regulation of anatomical structure size | 1/38 | 425/11590 | 0,759 | 0,863 | 0,814 | APOE | 1 |
| GO:0009991 | response to extracellular stimulus | 1/38 | 429/11590 | 0,762 | 0,865 | 0,816 | APOE | 1 |
| GO:0051052 | regulation of DNA metabolic process | 1/38 | 430/11590 | 0,763 | 0,865 | 0,816 | STN1 | 1 |
| GO:0051493 | regulation of cytoskeleton organization | 1/38 | 438/11590 | 0,769 | 0,871 | 0,822 | STAU2 | 1 |
| GO:0050778 | positive regulation of immune response | 1/38 | 441/11590 | 0,772 | 0,873 | 0,823 | HEXIM1 | 1 |
| GO:0006281 | DNA repair | 1/38 | 451/11590 | 0,779 | 0,881 | 0,831 | UFL1 | 1 |
| GO:0008015 | blood circulation | 1/38 | 452/11590 | 0,780 | 0,881 | 0,831 | APOE | 1 |

| | | | | | | | | |
|---|---|---|---|---|---|---|---|---|
| GO:0034762 | regulation of transmembrane transport | 1/38 | 454/11590 | 0,781 | 0,881 | 0,831 | SLC38A1 | 1 |
| GO:0051345 | positive regulation of hydrolase activity | 1/38 | 461/11590 | 0,787 | 0,886 | 0,836 | MTMR9 | 1 |
| GO:0007600 | sensory perception | 1/38 | 464/11590 | 0,789 | 0,888 | 0,837 | EFEMP1 | 1 |
| GO:0090407 | organophosphate biosynthetic process | 1/38 | 474/11590 | 0,796 | 0,894 | 0,843 | DCAKD | 1 |
| GO:0006511 | ubiquitin-dependent protein catabolic process | 1/38 | 475/11590 | 0,797 | 0,894 | 0,843 | UFL1 | 1 |
| GO:0044057 | regulation of system process | 1/38 | 479/11590 | 0,799 | 0,894 | 0,843 | APOE | 1 |
| GO:0005975 | carbohydrate metabolic process | 1/38 | 480/11590 | 0,800 | 0,894 | 0,843 | GALK1 | 1 |
| GO:0051347 | positive regulation of transferase activity | 1/38 | 480/11590 | 0,800 | 0,894 | 0,843 | APOE | 1 |
| GO:1903530 | regulation of secretion by cell | 1/38 | 480/11590 | 0,800 | 0,894 | 0,843 | APOE | 1 |
| GO:0007507 | heart development | 1/38 | 481/11590 | 0,801 | 0,894 | 0,843 | HEXIM1 | 1 |
| GO:0055080 | cation homeostasis | 1/38 | 483/11590 | 0,802 | 0,894 | 0,843 | APOE | 1 |
| GO:0019941 | modification-dependent protein catabolic process | 1/38 | 485/11590 | 0,803 | 0,895 | 0,844 | UFL1 | 1 |
| GO:0048598 | embryonic morphogenesis | 1/38 | 490/11590 | 0,807 | 0,897 | 0,846 | EFEMP1 | 1 |
| GO:1901137 | carbohydrate derivative biosynthetic process | 1/38 | 491/11590 | 0,807 | 0,897 | 0,846 | DCAKD | 1 |
| GO:0043632 | modification-dependent macromolecule catabolic process | 1/38 | 492/11590 | 0,808 | 0,897 | 0,846 | UFL1 | 1 |
| GO:0098771 | inorganic ion homeostasis | 1/38 | 496/11590 | 0,811 | 0,899 | 0,848 | APOE | 1 |
| GO:0001525 | angiogenesis | 0/38 | 464/11590 | 1,000 | 1,000 | 0,943 | | 0 |
| GO:0001764 | neuron migration | 0/38 | 142/11590 | 1,000 | 1,000 | 0,943 | | 0 |
| GO:0001890 | placenta development | 0/38 | 132/11590 | 1,000 | 1,000 | 0,943 | | 0 |
| GO:0001892 | embryonic placenta development | 0/38 | 78/11590 | 1,000 | 1,000 | 0,943 | | 0 |
| GO:0001906 | cell killing | 0/38 | 150/11590 | 1,000 | 1,000 | 0,943 | | 0 |
| GO:0001909 | leukocyte mediated cytotoxicity | 0/38 | 112/11590 | 1,000 | 1,000 | 0,943 | | 0 |
| GO:0002181 | cytoplasmic translation | 0/38 | 125/11590 | 1,000 | 1,000 | 0,943 | | 0 |
| GO:0002228 | natural killer cell mediated immunity | 0/38 | 60/11590 | 1,000 | 1,000 | 0,943 | | 0 |
| GO:0002250 | adaptive immune response | 0/38 | 389/11590 | 1,000 | 1,000 | 0,943 | | 0 |
| GO:0002263 | cell activation involved in immune response | 0/38 | 246/11590 | 1,000 | 1,000 | 0,943 | | 0 |
| GO:0002274 | myeloid leukocyte activation | 0/38 | 198/11590 | 1,000 | 1,000 | 0,943 | | 0 |

| | | | | | | | |
|---|---|---|---|---|---|---|---|
| GO:0002275 | myeloid cell activation involved in immune response | 0/38 | 86/11590 | 1,000 | 1,000 | 0,943 | 0 |
| GO:0002279 | mast cell activation involved in immune response | 0/38 | 49/11590 | 1,000 | 1,000 | 0,943 | 0 |
| GO:0002285 | lymphocyte activation involved in immune response | 0/38 | 165/11590 | 1,000 | 1,000 | 0,943 | 0 |
| GO:0002323 | natural killer cell activation involved in immune response | 0/38 | 24/11590 | 1,000 | 1,000 | 0,943 | 0 |
| GO:0002366 | leukocyte activation involved in immune response | 0/38 | 242/11590 | 1,000 | 1,000 | 0,943 | 0 |
| GO:0002443 | leukocyte mediated immunity | 0/38 | 336/11590 | 1,000 | 1,000 | 0,943 | 0 |
| GO:0002444 | myeloid leukocyte mediated immunity | 0/38 | 94/11590 | 1,000 | 1,000 | 0,943 | 0 |
| GO:0002448 | mast cell mediated immunity | 0/38 | 49/11590 | 1,000 | 1,000 | 0,943 | 0 |
| GO:0002449 | lymphocyte mediated immunity | 0/38 | 253/11590 | 1,000 | 1,000 | 0,943 | 0 |
| GO:0002460 | adaptive immune response based on somatic recombination of immune receptors built from immunoglobulin superfamily domains | 0/38 | 261/11590 | 1,000 | 1,000 | 0,943 | 0 |
| GO:0002467 | germinal center formation | 0/38 | 11/11590 | 1,000 | 1,000 | 0,943 | 0 |
| GO:0002544 | chronic inflammatory response | 0/38 | 18/11590 | 1,000 | 1,000 | 0,943 | 0 |
| GO:0002694 | regulation of leukocyte activation | 0/38 | 492/11590 | 1,000 | 1,000 | 0,943 | 0 |
| GO:0002697 | regulation of immune effector process | 0/38 | 319/11590 | 1,000 | 1,000 | 0,943 | 0 |
| GO:0002699 | positive regulation of immune effector process | 0/38 | 224/11590 | 1,000 | 1,000 | 0,943 | 0 |
| GO:0002703 | regulation of leukocyte mediated immunity | 0/38 | 203/11590 | 1,000 | 1,000 | 0,943 | 0 |
| GO:0002886 | regulation of myeloid leukocyte mediated immunity | 0/38 | 56/11590 | 1,000 | 1,000 | 0,943 | 0 |
| GO:0006352 | DNA-templated transcription initiation | 0/38 | 115/11590 | 1,000 | 1,000 | 0,943 | 0 |
| GO:0006367 | transcription initiation at RNA polymerase II promoter | 0/38 | 80/11590 | 1,000 | 1,000 | 0,943 | 0 |
| GO:0006417 | regulation of translation | 0/38 | 307/11590 | 1,000 | 1,000 | 0,943 | 0 |
| GO:0006887 | exocytosis | 0/38 | 288/11590 | 1,000 | 1,000 | 0,943 | 0 |
| GO:0006909 | phagocytosis | 0/38 | 211/11590 | 1,000 | 1,000 | 0,943 | 0 |
| GO:0007043 | cell-cell junction assembly | 0/38 | 124/11590 | 1,000 | 1,000 | 0,943 | 0 |

| | | | | | | | |
|---|---|---|---|---|---|---|---|
| GO:0007163 | establishment or maintenance of cell polarity | 0/38 | 196/11590 | 1,000 | 1,000 | 0,943 | 0 |
| GO:0007339 | binding of sperm to zona pellucida | 0/38 | 26/11590 | 1,000 | 1,000 | 0,943 | 0 |
| GO:0009116 | nucleoside metabolic process | 0/38 | 44/11590 | 1,000 | 1,000 | 0,943 | 0 |
| GO:0009119 | ribonucleoside metabolic process | 0/38 | 28/11590 | 1,000 | 1,000 | 0,943 | 0 |
| GO:0009163 | nucleoside biosynthetic process | 0/38 | 11/11590 | 1,000 | 1,000 | 0,943 | 0 |
| GO:0009615 | response to virus | 0/38 | 329/11590 | 1,000 | 1,000 | 0,943 | 0 |
| GO:0009988 | cell-cell recognition | 0/38 | 47/11590 | 1,000 | 1,000 | 0,943 | 0 |
| GO:0010608 | post-transcriptional regulation of gene expression | 0/38 | 393/11590 | 1,000 | 1,000 | 0,943 | 0 |
| GO:0010810 | regulation of cell-substrate adhesion | 0/38 | 194/11590 | 1,000 | 1,000 | 0,943 | 0 |
| GO:0010811 | positive regulation of cell-substrate adhesion | 0/38 | 113/11590 | 1,000 | 1,000 | 0,943 | 0 |
| GO:0017148 | negative regulation of translation | 0/38 | 131/11590 | 1,000 | 1,000 | 0,943 | 0 |
| GO:0017157 | regulation of exocytosis | 0/38 | 177/11590 | 1,000 | 1,000 | 0,943 | 0 |
| GO:0030010 | establishment of cell polarity | 0/38 | 125/11590 | 1,000 | 1,000 | 0,943 | 0 |
| GO:0030101 | natural killer cell activation | 0/38 | 78/11590 | 1,000 | 1,000 | 0,943 | 0 |
| GO:0032418 | lysosome localization | 0/38 | 70/11590 | 1,000 | 1,000 | 0,943 | 0 |
| GO:0032543 | mitochondrial translation | 0/38 | 88/11590 | 1,000 | 1,000 | 0,943 | 0 |
| GO:0033003 | regulation of mast cell activation | 0/38 | 41/11590 | 1,000 | 1,000 | 0,943 | 0 |
| GO:0033006 | regulation of mast cell activation involved in immune response | 0/38 | 32/11590 | 1,000 | 1,000 | 0,943 | 0 |
| GO:0034404 | nucleobase-containing small molecule biosynthetic process | 0/38 | 11/11590 | 1,000 | 1,000 | 0,943 | 0 |
| GO:0034446 | substrate adhesion-dependent cell spreading | 0/38 | 98/11590 | 1,000 | 1,000 | 0,943 | 0 |
| GO:0035036 | sperm-egg recognition | 0/38 | 30/11590 | 1,000 | 1,000 | 0,943 | 0 |
| GO:0042267 | natural killer cell mediated cytotoxicity | 0/38 | 59/11590 | 1,000 | 1,000 | 0,943 | 0 |
| GO:0042278 | purine nucleoside metabolic process | 0/38 | 20/11590 | 1,000 | 1,000 | 0,943 | 0 |
| GO:0043094 | cellular metabolic compound salvage | 0/38 | 22/11590 | 1,000 | 1,000 | 0,943 | 0 |
| GO:0043101 | purine-containing compound salvage | 0/38 | 12/11590 | 1,000 | 1,000 | 0,943 | 0 |
| GO:0043297 | apical junction assembly | 0/38 | 58/11590 | 1,000 | 1,000 | 0,943 | 0 |

| | | | | | | | |
|---|---|---|---|---|---|---|---|
| GO:0043299 | leukocyte degranulation | 0/38 | 69/11590 | 1,000 | 1,000 | 0,943 | 0 |
| GO:0043300 | regulation of leukocyte degranulation | 0/38 | 45/11590 | 1,000 | 1,000 | 0,943 | 0 |
| GO:0043303 | mast cell degranulation | 0/38 | 47/11590 | 1,000 | 1,000 | 0,943 | 0 |
| GO:0043304 | regulation of mast cell degranulation | 0/38 | 30/11590 | 1,000 | 1,000 | 0,943 | 0 |
| GO:0044782 | cilium organization | 0/38 | 236/11590 | 1,000 | 1,000 | 0,943 | 0 |
| GO:0045055 | regulated exocytosis | 0/38 | 195/11590 | 1,000 | 1,000 | 0,943 | 0 |
| GO:0045216 | cell-cell junction organization | 0/38 | 170/11590 | 1,000 | 1,000 | 0,943 | 0 |
| GO:0045576 | mast cell activation | 0/38 | 60/11590 | 1,000 | 1,000 | 0,943 | 0 |
| GO:0045785 | positive regulation of cell adhesion | 0/38 | 420/11590 | 1,000 | 1,000 | 0,943 | 0 |
| GO:0045921 | positive regulation of exocytosis | 0/38 | 76/11590 | 1,000 | 1,000 | 0,943 | 0 |
| GO:0046128 | purine ribonucleoside metabolic process | 0/38 | 17/11590 | 1,000 | 1,000 | 0,943 | 0 |
| GO:0048015 | phosphatidylinositol-mediated signaling | 0/38 | 151/11590 | 1,000 | 1,000 | 0,943 | 0 |
| GO:0048017 | inositol lipid-mediated signaling | 0/38 | 154/11590 | 1,000 | 1,000 | 0,943 | 0 |
| GO:0051047 | positive regulation of secretion | 0/38 | 271/11590 | 1,000 | 1,000 | 0,943 | 0 |
| GO:0051607 | defense response to virus | 0/38 | 239/11590 | 1,000 | 1,000 | 0,943 | 0 |
| GO:0051640 | organelle localization | 0/38 | 450/11590 | 1,000 | 1,000 | 0,943 | 0 |
| GO:0051656 | establishment of organelle localization | 0/38 | 347/11590 | 1,000 | 1,000 | 0,943 | 0 |
| GO:0051972 | regulation of telomerase activity | 0/38 | 38/11590 | 1,000 | 1,000 | 0,943 | 0 |
| GO:0060271 | cilium assembly | 0/38 | 225/11590 | 1,000 | 1,000 | 0,943 | 0 |
| GO:0060674 | placenta blood vessel development | 0/38 | 28/11590 | 1,000 | 1,000 | 0,943 | 0 |
| GO:0060711 | labyrinthine layer development | 0/38 | 43/11590 | 1,000 | 1,000 | 0,943 | 0 |
| GO:0060716 | labyrinthine layer blood vessel development | 0/38 | 18/11590 | 1,000 | 1,000 | 0,943 | 0 |
| GO:0090162 | establishment of epithelial cell polarity | 0/38 | 28/11590 | 1,000 | 1,000 | 0,943 | 0 |
| GO:0140053 | mitochondrial gene expression | 0/38 | 113/11590 | 1,000 | 1,000 | 0,943 | 0 |
| GO:0140546 | defense response to symbiont | 0/38 | 240/11590 | 1,000 | 1,000 | 0,943 | 0 |
| GO:1900024 | regulation of substrate adhesion-dependent cell spreading | 0/38 | 56/11590 | 1,000 | 1,000 | 0,943 | 0 |
| GO:1900026 | positive regulation of substrate adhesion-dependent cell spreading | 0/38 | 41/11590 | 1,000 | 1,000 | 0,943 | 0 |

| | | | | | | | |
|---|---|---|---|---|---|---|---|
| GO:1901657 | glycosyl compound metabolic process | 0/38 | 62/11590 | 1,000 | 1,000 | 0,943 | 0 |
| GO:1901659 | glycosyl compound biosynthetic process | 0/38 | 12/11590 | 1,000 | 1,000 | 0,943 | 0 |
| GO:1903305 | regulation of regulated secretory pathway | 0/38 | 121/11590 | 1,000 | 1,000 | 0,943 | 0 |
| GO:1903307 | positive regulation of regulated secretory pathway | 0/38 | 44/11590 | 1,000 | 1,000 | 0,943 | 0 |
| GO:1903532 | positive regulation of secretion by cell | 0/38 | 248/11590 | 1,000 | 1,000 | 0,943 | 0 |
| GO:1990849 | vacuolar localization | 0/38 | 70/11590 | 1,000 | 1,000 | 0,943 | 0 |
| GO:2000112 | regulation of cellular macromolecule biosynthetic process | 0/38 | 371/11590 | 1,000 | 1,000 | 0,943 | 0 |
| GO:2000113 | negative regulation of cellular macromolecule biosynthetic process | 0/38 | 147/11590 | 1,000 | 1,000 | 0,943 | 0 |
| GO:2000765 | regulation of cytoplasmic translation | 0/38 | 20/11590 | 1,000 | 1,000 | 0,943 | 0 |

**Supplementary Table 5.** Enrichment analysis results for the SNPs-related genes of the PRS-WMH before the clumping.
*Legend: GeneRatio: ratio of input genes that are annotated in a term (GeneRatio=k/n, where k: overlap of the SNPs-related gene IDs with the specific gene set related to a biological pathway; n: size of the overlap of the SNPs-related gene IDs with all the members of the collection of gene sets); BgRatio: ratio of all genes that are annotated in a term (BgRatio=count/setSize, where count: number of genes that belong to a given gene-set; setSize: total number of genes in the collection of gene sets); P-value: probability of seeing at least X number of genes out of the total n SNPs-related gene IDs in the list annotated to a particular GO term, given the proportion of genes in the whole genome that are annotated to that GO term; FDR: p-value after false discovery rate correction; Q-value: proportion of false positives incurred when the test is significant (FDR correction among the significant results); Counts: number of genes annotated to the GO term.*

| ID | Description | GeneRatio | BgRatio | pvalue | FDR | qvalue | geneID | Count |
|---|---|---|---|---|---|---|---|---|
| GO:0060999 | positive regulation of dendritic spine development | 2/17 | 30/11590 | 0,001 | 0,227 | 0,189 | APOE/STAU2 | 2 |
| GO:0048167 | regulation of synaptic plasticity | 3/17 | 158/11590 | 0,001 | 0,227 | 0,189 | APOE/SLC38A1/STAU2 | 3 |
| GO:0060998 | regulation of dendritic spine development | 2/17 | 44/11590 | 0,002 | 0,227 | 0,189 | APOE/STAU2 | 2 |
| GO:0097061 | dendritic spine organization | 2/17 | 66/11590 | 0,004 | 0,227 | 0,189 | APOE/STAU2 | 2 |
| GO:0099175 | regulation of postsynapse organization | 2/17 | 68/11590 | 0,004 | 0,227 | 0,189 | APOE/STAU2 | 2 |
| GO:0060996 | dendritic spine development | 2/17 | 76/11590 | 0,005 | 0,227 | 0,189 | APOE/STAU2 | 2 |
| GO:0106027 | neuron projection organization | 2/17 | 76/11590 | 0,005 | 0,227 | 0,189 | APOE/STAU2 | 2 |
| GO:0010232 | vascular transport | 2/17 | 77/11590 | 0,006 | 0,227 | 0,189 | APOE/SLC38A1 | 2 |
| GO:0150104 | transport across blood-brain barrier | 2/17 | 77/11590 | 0,006 | 0,227 | 0,189 | APOE/SLC38A1 | 2 |
| GO:0007266 | Rho protein signal transduction | 2/17 | 115/11590 | 0,012 | 0,227 | 0,189 | APOE/ARHGEF12 | 2 |
| GO:0050804 | modulation of chemical synaptic transmission | 3/17 | 354/11590 | 0,014 | 0,227 | 0,189 | APOE/SLC38A1/STAU2 | 3 |
| GO:0099177 | regulation of trans-synaptic signaling | 3/17 | 355/11590 | 0,014 | 0,227 | 0,189 | APOE/SLC38A1/STAU2 | 3 |
| GO:0006707 | cholesterol catabolic process | 1/17 | 10/11590 | 0,015 | 0,227 | 0,189 | APOE | 1 |
| GO:0008298 | intracellular mRNA localization | 1/17 | 10/11590 | 0,015 | 0,227 | 0,189 | STAU2 | 1 |
| GO:0015937 | coenzyme A biosynthetic process | 1/17 | 10/11590 | 0,015 | 0,227 | 0,189 | DCAKD | 1 |
| GO:0016127 | sterol catabolic process | 1/17 | 10/11590 | 0,015 | 0,227 | 0,189 | APOE | 1 |
| GO:0034370 | triglyceride-rich lipoprotein particle remodeling | 1/17 | 10/11590 | 0,015 | 0,227 | 0,189 | APOE | 1 |
| GO:0034372 | very-low-density lipoprotein particle remodeling | 1/17 | 10/11590 | 0,015 | 0,227 | 0,189 | APOE | 1 |
| GO:0034384 | high-density lipoprotein particle clearance | 1/17 | 10/11590 | 0,015 | 0,227 | 0,189 | APOE | 1 |
| GO:1903365 | regulation of fear response | 1/17 | 10/11590 | 0,015 | 0,227 | 0,189 | APOE | 1 |
| GO:1903961 | positive regulation of anion transmembrane transport | 1/17 | 10/11590 | 0,015 | 0,227 | 0,189 | SLC38A1 | 1 |
| GO:0099173 | postsynapse organization | 2/17 | 131/11590 | 0,015 | 0,227 | 0,189 | APOE/STAU2 | 2 |
| GO:0033700 | phospholipid efflux | 1/17 | 11/11590 | 0,016 | 0,227 | 0,189 | APOE | 1 |

| | | | | | | | | |
|---|---|---|---|---|---|---|---|---|
| GO:0042159 | lipoprotein catabolic process | 1/17 | 11/11590 | 0,016 | 0,227 | 0,189 | APOE | 1 |
| GO:0090205 | positive regulation of cholesterol metabolic process | 1/17 | 11/11590 | 0,016 | 0,227 | 0,189 | APOE | 1 |
| GO:1900272 | negative regulation of long-term synaptic potentiation | 1/17 | 11/11590 | 0,016 | 0,227 | 0,189 | APOE | 1 |
| GO:1902950 | regulation of dendritic spine maintenance | 1/17 | 11/11590 | 0,016 | 0,227 | 0,189 | APOE | 1 |
| GO:0010976 | positive regulation of neuron projection development | 2/17 | 134/11590 | 0,016 | 0,227 | 0,189 | APOE/STAU2 | 2 |
| GO:0048592 | eye morphogenesis | 2/17 | 136/11590 | 0,017 | 0,227 | 0,189 | EFEMP1/STAU2 | 2 |
| GO:0009886 | post-embryonic animal morphogenesis | 1/17 | 12/11590 | 0,017 | 0,227 | 0,189 | EFEMP1 | 1 |
| GO:0032328 | alanine transport | 1/17 | 12/11590 | 0,017 | 0,227 | 0,189 | SLC38A1 | 1 |
| GO:0032488 | Cdc42 protein signal transduction | 1/17 | 12/11590 | 0,017 | 0,227 | 0,189 | APOE | 1 |
| GO:0034380 | high-density lipoprotein particle assembly | 1/17 | 12/11590 | 0,017 | 0,227 | 0,189 | APOE | 1 |
| GO:0051044 | positive regulation of membrane protein ectodomain proteolysis | 1/17 | 12/11590 | 0,017 | 0,227 | 0,189 | APOE | 1 |
| GO:1900452 | regulation of long-term synaptic depression | 1/17 | 12/11590 | 0,017 | 0,227 | 0,189 | STAU2 | 1 |
| GO:1905907 | negative regulation of amyloid fibril formation | 1/17 | 12/11590 | 0,017 | 0,227 | 0,189 | APOE | 1 |
| GO:2001140 | positive regulation of phospholipid transport | 1/17 | 12/11590 | 0,017 | 0,227 | 0,189 | APOE | 1 |
| GO:0035641 | locomotory exploration behavior | 1/17 | 13/11590 | 0,019 | 0,227 | 0,189 | APOE | 1 |
| GO:0055089 | fatty acid homeostasis | 1/17 | 13/11590 | 0,019 | 0,227 | 0,189 | APOE | 1 |
| GO:0098935 | dendritic transport | 1/17 | 13/11590 | 0,019 | 0,227 | 0,189 | STAU2 | 1 |
| GO:1902931 | negative regulation of alcohol biosynthetic process | 1/17 | 13/11590 | 0,019 | 0,227 | 0,189 | APOE | 1 |
| GO:2001138 | regulation of phospholipid transport | 1/17 | 13/11590 | 0,019 | 0,227 | 0,189 | APOE | 1 |
| GO:0044794 | positive regulation by host of viral process | 1/17 | 14/11590 | 0,020 | 0,227 | 0,189 | APOE | 1 |
| GO:1905906 | regulation of amyloid fibril formation | 1/17 | 14/11590 | 0,020 | 0,227 | 0,189 | APOE | 1 |

| | | | | | | | | |
|---|---|---|---|---|---|---|---|---|
| GO:0010958 | regulation of amino acid import across plasma membrane | 1/17 | 15/11590 | 0,022 | 0,227 | 0,189 | SLC38A1 | 1 |
| GO:0015936 | coenzyme A metabolic process | 1/17 | 15/11590 | 0,022 | 0,227 | 0,189 | DCAKD | 1 |
| GO:0034374 | low-density lipoprotein particle remodeling | 1/17 | 15/11590 | 0,022 | 0,227 | 0,189 | APOE | 1 |
| GO:0043691 | reverse cholesterol transport | 1/17 | 15/11590 | 0,022 | 0,227 | 0,189 | APOE | 1 |
| GO:0061003 | positive regulation of dendritic spine morphogenesis | 1/17 | 15/11590 | 0,022 | 0,227 | 0,189 | STAU2 | 1 |
| GO:1903789 | regulation of amino acid transmembrane transport | 1/17 | 15/11590 | 0,022 | 0,227 | 0,189 | SLC38A1 | 1 |
| GO:0050807 | regulation of synapse organization | 2/17 | 158/11590 | 0,022 | 0,227 | 0,189 | APOE/STAU2 | 2 |
| GO:0050803 | regulation of synapse structure or activity | 2/17 | 161/11590 | 0,023 | 0,227 | 0,189 | APOE/STAU2 | 2 |
| GO:0034375 | high-density lipoprotein particle remodeling | 1/17 | 16/11590 | 0,023 | 0,227 | 0,189 | APOE | 1 |
| GO:0048569 | post-embryonic animal organ development | 1/17 | 16/11590 | 0,023 | 0,227 | 0,189 | EFEMP1 | 1 |
| GO:1900221 | regulation of amyloid-beta clearance | 1/17 | 16/11590 | 0,023 | 0,227 | 0,189 | APOE | 1 |
| GO:0030100 | regulation of endocytosis | 2/17 | 164/11590 | 0,024 | 0,227 | 0,189 | APOE/STON1 | 2 |
| GO:0010544 | negative regulation of platelet activation | 1/17 | 17/11590 | 0,025 | 0,227 | 0,189 | APOE | 1 |
| GO:0043117 | positive regulation of vascular permeability | 1/17 | 17/11590 | 0,025 | 0,227 | 0,189 | APOE | 1 |
| GO:1902430 | negative regulation of amyloid-beta formation | 1/17 | 17/11590 | 0,025 | 0,227 | 0,189 | APOE | 1 |
| GO:0045540 | regulation of cholesterol biosynthetic process | 1/17 | 18/11590 | 0,026 | 0,227 | 0,189 | APOE | 1 |
| GO:0106118 | regulation of sterol biosynthetic process | 1/17 | 18/11590 | 0,026 | 0,227 | 0,189 | APOE | 1 |
| GO:0051000 | positive regulation of nitric-oxide synthase activity | 1/17 | 19/11590 | 0,028 | 0,227 | 0,189 | APOE | 1 |
| GO:0097062 | dendritic spine maintenance | 1/17 | 19/11590 | 0,028 | 0,227 | 0,189 | APOE | 1 |
| GO:0032331 | negative regulation of chondrocyte differentiation | 1/17 | 20/11590 | 0,029 | 0,227 | 0,189 | EFEMP1 | 1 |

| | | | | | | | | |
|---|---|---|---|---|---|---|---|---|
| GO:0051043 | regulation of membrane protein ectodomain proteolysis | 1/17 | 20/11590 | 0,029 | 0,227 | 0,189 | APOE | 1 |
| GO:1902992 | negative regulation of amyloid precursor protein catabolic process | 1/17 | 20/11590 | 0,029 | 0,227 | 0,189 | APOE | 1 |
| GO:1903959 | regulation of anion transmembrane transport | 1/17 | 21/11590 | 0,030 | 0,227 | 0,189 | SLC38A1 | 1 |
| GO:0006897 | endocytosis | 3/17 | 480/11590 | 0,031 | 0,227 | 0,189 | APOE/MTMR9/STON1 | 3 |
| GO:0006541 | glutamine metabolic process | 1/17 | 22/11590 | 0,032 | 0,227 | 0,189 | SLC38A1 | 1 |
| GO:0010894 | negative regulation of steroid biosynthetic process | 1/17 | 22/11590 | 0,032 | 0,227 | 0,189 | APOE | 1 |
| GO:0032801 | receptor catabolic process | 1/17 | 22/11590 | 0,032 | 0,227 | 0,189 | APOE | 1 |
| GO:0034377 | plasma lipoprotein particle assembly | 1/17 | 22/11590 | 0,032 | 0,227 | 0,189 | APOE | 1 |
| GO:0051957 | positive regulation of amino acid transport | 1/17 | 22/11590 | 0,032 | 0,227 | 0,189 | SLC38A1 | 1 |
| GO:0150146 | cell junction disassembly | 1/17 | 22/11590 | 0,032 | 0,227 | 0,189 | STON1 | 1 |
| GO:0016358 | dendrite development | 2/17 | 194/11590 | 0,032 | 0,227 | 0,189 | APOE/STAU2 | 2 |
| GO:0006929 | substrate-dependent cell migration | 1/17 | 23/11590 | 0,033 | 0,227 | 0,189 | STON1 | 1 |
| GO:0010875 | positive regulation of cholesterol efflux | 1/17 | 23/11590 | 0,033 | 0,227 | 0,189 | APOE | 1 |
| GO:0006891 | intra-Golgi vesicle-mediated transport | 1/17 | 24/11590 | 0,035 | 0,227 | 0,189 | COG2 | 1 |
| GO:0035640 | exploration behavior | 1/17 | 24/11590 | 0,035 | 0,227 | 0,189 | APOE | 1 |
| GO:0045939 | negative regulation of steroid metabolic process | 1/17 | 24/11590 | 0,035 | 0,227 | 0,189 | APOE | 1 |
| GO:0045940 | positive regulation of steroid metabolic process | 1/17 | 24/11590 | 0,035 | 0,227 | 0,189 | APOE | 1 |
| GO:0065005 | protein-lipid complex assembly | 1/17 | 24/11590 | 0,035 | 0,227 | 0,189 | APOE | 1 |
| GO:0019068 | virion assembly | 1/17 | 25/11590 | 0,036 | 0,227 | 0,189 | APOE | 1 |
| GO:0055090 | acylglycerol homeostasis | 1/17 | 25/11590 | 0,036 | 0,227 | 0,189 | APOE | 1 |
| GO:0060292 | long-term synaptic depression | 1/17 | 25/11590 | 0,036 | 0,227 | 0,189 | STAU2 | 1 |
| GO:0061037 | negative regulation of cartilage development | 1/17 | 25/11590 | 0,036 | 0,227 | 0,189 | EFEMP1 | 1 |
| GO:0070328 | triglyceride homeostasis | 1/17 | 25/11590 | 0,036 | 0,227 | 0,189 | APOE | 1 |

| | | | | | | | | |
|---|---|---|---|---|---|---|---|---|
| GO:0002021 | response to dietary excess | 1/17 | 26/11590 | 0,037 | 0,227 | 0,189 | APOE | 1 |
| GO:0006706 | steroid catabolic process | 1/17 | 26/11590 | 0,037 | 0,227 | 0,189 | APOE | 1 |
| GO:0007263 | nitric oxide mediated signal transduction | 1/17 | 26/11590 | 0,037 | 0,227 | 0,189 | APOE | 1 |
| GO:0090596 | sensory organ morphogenesis | 2/17 | 215/11590 | 0,039 | 0,227 | 0,189 | EFEMP1/STAU2 | 2 |
| GO:0007271 | synaptic transmission, cholinergic | 1/17 | 27/11590 | 0,039 | 0,227 | 0,189 | APOE | 1 |
| GO:0010922 | positive regulation of phosphatase activity | 1/17 | 27/11590 | 0,039 | 0,227 | 0,189 | MTMR9 | 1 |
| GO:0034368 | protein-lipid complex remodeling | 1/17 | 27/11590 | 0,039 | 0,227 | 0,189 | APOE | 1 |
| GO:0034369 | plasma lipoprotein particle remodeling | 1/17 | 27/11590 | 0,039 | 0,227 | 0,189 | APOE | 1 |
| GO:0001941 | postsynaptic membrane organization | 1/17 | 28/11590 | 0,040 | 0,227 | 0,189 | APOE | 1 |
| GO:0034367 | protein-containing complex remodeling | 1/17 | 28/11590 | 0,040 | 0,227 | 0,189 | APOE | 1 |
| GO:0046856 | phosphatidylinositol dephosphorylation | 1/17 | 28/11590 | 0,040 | 0,227 | 0,189 | MTMR9 | 1 |
| GO:0055094 | response to lipoprotein particle | 1/17 | 28/11590 | 0,040 | 0,227 | 0,189 | APOE | 1 |
| GO:0090181 | regulation of cholesterol metabolic process | 1/17 | 28/11590 | 0,040 | 0,227 | 0,189 | APOE | 1 |
| GO:0090207 | regulation of triglyceride metabolic process | 1/17 | 28/11590 | 0,040 | 0,227 | 0,189 | APOE | 1 |
| GO:0010874 | regulation of cholesterol efflux | 1/17 | 29/11590 | 0,042 | 0,227 | 0,189 | APOE | 1 |
| GO:0032770 | positive regulation of monooxygenase activity | 1/17 | 29/11590 | 0,042 | 0,227 | 0,189 | APOE | 1 |
| GO:0050775 | positive regulation of dendrite morphogenesis | 1/17 | 29/11590 | 0,042 | 0,227 | 0,189 | STAU2 | 1 |
| GO:0007616 | long-term memory | 1/17 | 30/11590 | 0,043 | 0,227 | 0,189 | APOE | 1 |
| GO:0032228 | regulation of synaptic transmission, GABAergic | 1/17 | 30/11590 | 0,043 | 0,227 | 0,189 | SLC38A1 | 1 |
| GO:0032373 | positive regulation of sterol transport | 1/17 | 31/11590 | 0,045 | 0,227 | 0,189 | APOE | 1 |
| GO:0032376 | positive regulation of cholesterol transport | 1/17 | 31/11590 | 0,045 | 0,227 | 0,189 | APOE | 1 |
| GO:0071402 | cellular response to lipoprotein particle stimulus | 1/17 | 31/11590 | 0,045 | 0,227 | 0,189 | APOE | 1 |

| | | | | | | | | |
|---|---|---|---|---|---|---|---|---|
| GO:1903725 | regulation of phospholipid metabolic process | 1/17 | 31/11590 | 0,045 | 0,227 | 0,189 | MTMR9 | 1 |
| GO:0051056 | regulation of small GTPase mediated signal transduction | 2/17 | 240/11590 | 0,047 | 0,227 | 0,189 | APOE/ARHGEF12 | 2 |
| GO:0019934 | cGMP-mediated signaling | 1/17 | 33/11590 | 0,047 | 0,227 | 0,189 | APOE | 1 |
| GO:0043537 | negative regulation of blood vessel endothelial cell migration | 1/17 | 33/11590 | 0,047 | 0,227 | 0,189 | APOE | 1 |
| GO:0048048 | embryonic eye morphogenesis | 1/17 | 33/11590 | 0,047 | 0,227 | 0,189 | EFEMP1 | 1 |
| GO:0050999 | regulation of nitric-oxide synthase activity | 1/17 | 33/11590 | 0,047 | 0,227 | 0,189 | APOE | 1 |
| GO:0061001 | regulation of dendritic spine morphogenesis | 1/17 | 33/11590 | 0,047 | 0,227 | 0,189 | STAU2 | 1 |
| GO:1902003 | regulation of amyloid-beta formation | 1/17 | 33/11590 | 0,047 | 0,227 | 0,189 | APOE | 1 |
| GO:0034381 | plasma lipoprotein particle clearance | 1/17 | 34/11590 | 0,049 | 0,227 | 0,189 | APOE | 1 |
| GO:0042554 | superoxide anion generation | 1/17 | 34/11590 | 0,049 | 0,227 | 0,189 | SH3PXD2A | 1 |
| GO:1990000 | amyloid fibril formation | 1/17 | 34/11590 | 0,049 | 0,227 | 0,189 | APOE | 1 |
| GO:0003018 | vascular process in circulatory system | 2/17 | 244/11590 | 0,049 | 0,227 | 0,189 | APOE/SLC38A1 | 2 |
| GO:0001662 | behavioral fear response | 1/17 | 35/11590 | 0,050 | 0,227 | 0,189 | APOE | 1 |
| GO:0097242 | amyloid-beta clearance | 1/17 | 35/11590 | 0,050 | 0,227 | 0,189 | APOE | 1 |
| GO:0015849 | organic acid transport | 2/17 | 248/11590 | 0,050 | 0,227 | 0,189 | APOE/SLC38A1 | 2 |
| GO:0002209 | behavioral defense response | 1/17 | 36/11590 | 0,052 | 0,227 | 0,189 | APOE | 1 |
| GO:0046839 | phospholipid dephosphorylation | 1/17 | 36/11590 | 0,052 | 0,227 | 0,189 | MTMR9 | 1 |
| GO:0001937 | negative regulation of endothelial cell proliferation | 1/17 | 37/11590 | 0,053 | 0,227 | 0,189 | APOE | 1 |
| GO:0044788 | modulation by host of viral process | 1/17 | 37/11590 | 0,053 | 0,227 | 0,189 | APOE | 1 |
| GO:0006509 | membrane protein ectodomain proteolysis | 1/17 | 38/11590 | 0,054 | 0,227 | 0,189 | APOE | 1 |
| GO:0097178 | ruffle assembly | 1/17 | 38/11590 | 0,054 | 0,227 | 0,189 | STON1 | 1 |
| GO:1900271 | regulation of long-term synaptic potentiation | 1/17 | 38/11590 | 0,054 | 0,227 | 0,189 | APOE | 1 |
| GO:0051954 | positive regulation of amine transport | 1/17 | 39/11590 | 0,056 | 0,227 | 0,189 | SLC38A1 | 1 |

| | | | | | | | | |
|---|---|---|---|---|---|---|---|---|
| GO:0071827 | plasma lipoprotein particle organization | 1/17 | 39/11590 | 0,056 | 0,227 | 0,189 | APOE | 1 |
| GO:0120009 | intermembrane lipid transfer | 1/17 | 39/11590 | 0,056 | 0,227 | 0,189 | APOE | 1 |
| GO:1902991 | regulation of amyloid precursor protein catabolic process | 1/17 | 39/11590 | 0,056 | 0,227 | 0,189 | APOE | 1 |
| GO:0051955 | regulation of amino acid transport | 1/17 | 40/11590 | 0,057 | 0,227 | 0,189 | SLC38A1 | 1 |
| GO:0015804 | neutral amino acid transport | 1/17 | 41/11590 | 0,059 | 0,227 | 0,189 | SLC38A1 | 1 |
| GO:0032892 | positive regulation of organic acid transport | 1/17 | 41/11590 | 0,059 | 0,227 | 0,189 | SLC38A1 | 1 |
| GO:0034205 | amyloid-beta formation | 1/17 | 41/11590 | 0,059 | 0,227 | 0,189 | APOE | 1 |
| GO:0042596 | fear response | 1/17 | 41/11590 | 0,059 | 0,227 | 0,189 | APOE | 1 |
| GO:0071825 | protein-lipid complex subunit organization | 1/17 | 41/11590 | 0,059 | 0,227 | 0,189 | APOE | 1 |
| GO:0001504 | neurotransmitter uptake | 1/17 | 42/11590 | 0,060 | 0,227 | 0,189 | SLC38A1 | 1 |
| GO:0019216 | regulation of lipid metabolic process | 2/17 | 276/11590 | 0,061 | 0,227 | 0,189 | APOE/MTMR9 | 2 |
| GO:0032330 | regulation of chondrocyte differentiation | 1/17 | 43/11590 | 0,061 | 0,227 | 0,189 | EFEMP1 | 1 |
| GO:0051965 | positive regulation of synapse assembly | 1/17 | 43/11590 | 0,061 | 0,227 | 0,189 | STAU2 | 1 |
| GO:0033344 | cholesterol efflux | 1/17 | 44/11590 | 0,063 | 0,227 | 0,189 | APOE | 1 |
| GO:0051489 | regulation of filopodium assembly | 1/17 | 44/11590 | 0,063 | 0,227 | 0,189 | STAU2 | 1 |
| GO:1903793 | positive regulation of anion transport | 1/17 | 44/11590 | 0,063 | 0,227 | 0,189 | SLC38A1 | 1 |
| GO:0032371 | regulation of sterol transport | 1/17 | 45/11590 | 0,064 | 0,227 | 0,189 | APOE | 1 |
| GO:0032374 | regulation of cholesterol transport | 1/17 | 45/11590 | 0,064 | 0,227 | 0,189 | APOE | 1 |
| GO:0032768 | regulation of monooxygenase activity | 1/17 | 45/11590 | 0,064 | 0,227 | 0,189 | APOE | 1 |
| GO:0043114 | regulation of vascular permeability | 1/17 | 45/11590 | 0,064 | 0,227 | 0,189 | APOE | 1 |
| GO:0048168 | regulation of neuronal synaptic plasticity | 1/17 | 45/11590 | 0,064 | 0,227 | 0,189 | APOE | 1 |
| GO:0089718 | amino acid import across plasma membrane | 1/17 | 45/11590 | 0,064 | 0,227 | 0,189 | SLC38A1 | 1 |
| GO:0007265 | Ras protein signal transduction | 2/17 | 285/11590 | 0,064 | 0,227 | 0,189 | APOE/ARHGEF12 | 2 |

| | | | | | | | | |
|---|---|---|---|---|---|---|---|---|
| GO:0010543 | regulation of platelet activation | 1/17 | 46/11590 | 0,065 | 0,227 | 0,189 | APOE | 1 |
| GO:0042311 | vasodilation | 1/17 | 46/11590 | 0,065 | 0,227 | 0,189 | APOE | 1 |
| GO:0043113 | receptor clustering | 1/17 | 46/11590 | 0,065 | 0,227 | 0,189 | APOE | 1 |
| GO:0051055 | negative regulation of lipid biosynthetic process | 1/17 | 46/11590 | 0,065 | 0,227 | 0,189 | APOE | 1 |
| GO:0006695 | cholesterol biosynthetic process | 1/17 | 47/11590 | 0,067 | 0,227 | 0,189 | APOE | 1 |
| GO:0030195 | negative regulation of blood coagulation | 1/17 | 47/11590 | 0,067 | 0,227 | 0,189 | APOE | 1 |
| GO:0033619 | membrane protein proteolysis | 1/17 | 47/11590 | 0,067 | 0,227 | 0,189 | APOE | 1 |
| GO:0046164 | alcohol catabolic process | 1/17 | 47/11590 | 0,067 | 0,227 | 0,189 | APOE | 1 |
| GO:0060997 | dendritic spine morphogenesis | 1/17 | 47/11590 | 0,067 | 0,227 | 0,189 | STAU2 | 1 |
| GO:1902653 | secondary alcohol biosynthetic process | 1/17 | 47/11590 | 0,067 | 0,227 | 0,189 | APOE | 1 |
| GO:1902930 | regulation of alcohol biosynthetic process | 1/17 | 47/11590 | 0,067 | 0,227 | 0,189 | APOE | 1 |
| GO:0010596 | negative regulation of endothelial cell migration | 1/17 | 48/11590 | 0,068 | 0,227 | 0,189 | APOE | 1 |
| GO:0035306 | positive regulation of dephosphorylation | 1/17 | 48/11590 | 0,068 | 0,227 | 0,189 | MTMR9 | 1 |
| GO:0048662 | negative regulation of smooth muscle cell proliferation | 1/17 | 48/11590 | 0,068 | 0,227 | 0,189 | APOE | 1 |
| GO:0050709 | negative regulation of protein secretion | 1/17 | 48/11590 | 0,068 | 0,227 | 0,189 | APOE | 1 |
| GO:0051932 | synaptic transmission, GABAergic | 1/17 | 48/11590 | 0,068 | 0,227 | 0,189 | SLC38A1 | 1 |
| GO:1900047 | negative regulation of hemostasis | 1/17 | 48/11590 | 0,068 | 0,227 | 0,189 | APOE | 1 |
| GO:0046486 | glycerolipid metabolic process | 2/17 | 297/11590 | 0,069 | 0,227 | 0,189 | APOE/MTMR9 | 2 |
| GO:0043112 | receptor metabolic process | 1/17 | 49/11590 | 0,070 | 0,227 | 0,189 | APOE | 1 |
| GO:0043407 | negative regulation of MAP kinase activity | 1/17 | 49/11590 | 0,070 | 0,227 | 0,189 | APOE | 1 |
| GO:0050819 | negative regulation of coagulation | 1/17 | 49/11590 | 0,070 | 0,227 | 0,189 | APOE | 1 |
| GO:0031346 | positive regulation of cell projection organization | 2/17 | 300/11590 | 0,070 | 0,227 | 0,189 | APOE/STAU2 | 2 |

| | | | | | | | | |
|---|---|---|---|---|---|---|---|---|
| GO:0000768 | syncytium formation by plasma membrane fusion | 1/17 | 50/11590 | 0,071 | 0,227 | 0,189 | SH3PXD2A | 1 |
| GO:0031529 | ruffle organization | 1/17 | 50/11590 | 0,071 | 0,227 | 0,189 | STON1 | 1 |
| GO:0033866 | nucleoside bisphosphate biosynthetic process | 1/17 | 50/11590 | 0,071 | 0,227 | 0,189 | DCAKD | 1 |
| GO:0034030 | ribonucleoside bisphosphate biosynthetic process | 1/17 | 50/11590 | 0,071 | 0,227 | 0,189 | DCAKD | 1 |
| GO:0034033 | purine nucleoside bisphosphate biosynthetic process | 1/17 | 50/11590 | 0,071 | 0,227 | 0,189 | DCAKD | 1 |
| GO:0050435 | amyloid-beta metabolic process | 1/17 | 50/11590 | 0,071 | 0,227 | 0,189 | APOE | 1 |
| GO:0140253 | cell-cell fusion | 1/17 | 50/11590 | 0,071 | 0,227 | 0,189 | SH3PXD2A | 1 |
| GO:0051353 | positive regulation of oxidoreductase activity | 1/17 | 51/11590 | 0,072 | 0,230 | 0,192 | APOE | 1 |
| GO:0042987 | amyloid precursor protein catabolic process | 1/17 | 52/11590 | 0,074 | 0,230 | 0,193 | APOE | 1 |
| GO:0043954 | cellular component maintenance | 1/17 | 52/11590 | 0,074 | 0,230 | 0,193 | APOE | 1 |
| GO:0048008 | platelet-derived growth factor receptor signaling pathway | 1/17 | 52/11590 | 0,074 | 0,230 | 0,193 | STON1 | 1 |
| GO:0006949 | syncytium formation | 1/17 | 53/11590 | 0,075 | 0,231 | 0,193 | SH3PXD2A | 1 |
| GO:0048488 | synaptic vesicle endocytosis | 1/17 | 53/11590 | 0,075 | 0,231 | 0,193 | STON1 | 1 |
| GO:0140238 | presynaptic endocytosis | 1/17 | 53/11590 | 0,075 | 0,231 | 0,193 | STON1 | 1 |
| GO:0016126 | sterol biosynthetic process | 1/17 | 54/11590 | 0,076 | 0,234 | 0,196 | APOE | 1 |
| GO:0001654 | eye development | 2/17 | 316/11590 | 0,077 | 0,235 | 0,196 | EFEMP1/STAU2 | 2 |
| GO:0015909 | long-chain fatty acid transport | 1/17 | 55/11590 | 0,078 | 0,235 | 0,196 | APOE | 1 |
| GO:0150063 | visual system development | 2/17 | 319/11590 | 0,078 | 0,235 | 0,196 | EFEMP1/STAU2 | 2 |
| GO:0006801 | superoxide metabolic process | 1/17 | 56/11590 | 0,079 | 0,235 | 0,196 | SH3PXD2A | 1 |
| GO:0046847 | filopodium assembly | 1/17 | 56/11590 | 0,079 | 0,235 | 0,196 | STAU2 | 1 |
| GO:0048814 | regulation of dendrite morphogenesis | 1/17 | 56/11590 | 0,079 | 0,235 | 0,196 | STAU2 | 1 |
| GO:0048880 | sensory system development | 2/17 | 324/11590 | 0,080 | 0,237 | 0,198 | EFEMP1/STAU2 | 2 |
| GO:0035418 | protein localization to synapse | 1/17 | 57/11590 | 0,080 | 0,237 | 0,198 | STAU2 | 1 |
| GO:0015807 | L-amino acid transport | 1/17 | 58/11590 | 0,082 | 0,239 | 0,200 | SLC38A1 | 1 |

| | | | | | | | | |
|---|---|---|---|---|---|---|---|---|
| GO:0050805 | negative regulation of synaptic transmission | 1/17 | 59/11590 | 0,083 | 0,239 | 0,200 | STAU2 | 1 |
| GO:0061035 | regulation of cartilage development | 1/17 | 59/11590 | 0,083 | 0,239 | 0,200 | EFEMP1 | 1 |
| GO:1902475 | L-alpha-amino acid transmembrane transport | 1/17 | 59/11590 | 0,083 | 0,239 | 0,200 | SLC38A1 | 1 |
| GO:0050808 | synapse organization | 2/17 | 331/11590 | 0,083 | 0,239 | 0,200 | APOE/STAU2 | 2 |
| GO:0008088 | axo-dendritic transport | 1/17 | 60/11590 | 0,085 | 0,239 | 0,200 | STAU2 | 1 |
| GO:0050795 | regulation of behavior | 1/17 | 60/11590 | 0,085 | 0,239 | 0,200 | APOE | 1 |
| GO:0097006 | regulation of plasma lipoprotein particle levels | 1/17 | 60/11590 | 0,085 | 0,239 | 0,200 | APOE | 1 |
| GO:0010633 | negative regulation of epithelial cell migration | 1/17 | 61/11590 | 0,086 | 0,240 | 0,201 | APOE | 1 |
| GO:0050810 | regulation of steroid biosynthetic process | 1/17 | 61/11590 | 0,086 | 0,240 | 0,201 | APOE | 1 |
| GO:0034329 | cell junction assembly | 2/17 | 339/11590 | 0,087 | 0,241 | 0,201 | STON1/STAU2 | 2 |
| GO:0009064 | glutamine family amino acid metabolic process | 1/17 | 62/11590 | 0,087 | 0,241 | 0,201 | SLC38A1 | 1 |
| GO:0036465 | synaptic vesicle recycling | 1/17 | 62/11590 | 0,087 | 0,241 | 0,201 | STON1 | 1 |
| GO:0010921 | regulation of phosphatase activity | 1/17 | 63/11590 | 0,089 | 0,243 | 0,203 | MTMR9 | 1 |
| GO:0048844 | artery morphogenesis | 1/17 | 65/11590 | 0,091 | 0,247 | 0,206 | APOE | 1 |
| GO:0008652 | cellular amino acid biosynthetic process | 1/17 | 66/11590 | 0,093 | 0,247 | 0,206 | SLC38A1 | 1 |
| GO:0019915 | lipid storage | 1/17 | 66/11590 | 0,093 | 0,247 | 0,206 | APOE | 1 |
| GO:0030193 | regulation of blood coagulation | 1/17 | 66/11590 | 0,093 | 0,247 | 0,206 | APOE | 1 |
| GO:0061045 | negative regulation of wound healing | 1/17 | 66/11590 | 0,093 | 0,247 | 0,206 | APOE | 1 |
| GO:1901616 | organic hydroxy compound catabolic process | 1/17 | 66/11590 | 0,093 | 0,247 | 0,206 | APOE | 1 |
| GO:0006979 | response to oxidative stress | 2/17 | 352/11590 | 0,093 | 0,247 | 0,206 | APOE/STAU2 | 2 |
| GO:0032890 | regulation of organic acid transport | 1/17 | 67/11590 | 0,094 | 0,248 | 0,207 | SLC38A1 | 1 |
| GO:0042982 | amyloid precursor protein metabolic process | 1/17 | 67/11590 | 0,094 | 0,248 | 0,207 | APOE | 1 |

| | | | | | | | | |
|---|---|---|---|---|---|---|---|---|
| GO:0051851 | modulation by host of symbiont process | 1/17 | 68/11590 | 0,095 | 0,249 | 0,208 | APOE | 1 |
| GO:1900046 | regulation of hemostasis | 1/17 | 68/11590 | 0,095 | 0,249 | 0,208 | APOE | 1 |
| GO:0046889 | positive regulation of lipid biosynthetic process | 1/17 | 69/11590 | 0,097 | 0,250 | 0,209 | APOE | 1 |
| GO:0050818 | regulation of coagulation | 1/17 | 69/11590 | 0,097 | 0,250 | 0,209 | APOE | 1 |
| GO:0010507 | negative regulation of autophagy | 1/17 | 70/11590 | 0,098 | 0,250 | 0,209 | MTMR9 | 1 |
| GO:0042158 | lipoprotein biosynthetic process | 1/17 | 70/11590 | 0,098 | 0,250 | 0,209 | APOE | 1 |
| GO:0042632 | cholesterol homeostasis | 1/17 | 70/11590 | 0,098 | 0,250 | 0,209 | APOE | 1 |
| GO:0055092 | sterol homeostasis | 1/17 | 71/11590 | 0,099 | 0,252 | 0,210 | APOE | 1 |
| GO:0098869 | cellular oxidant detoxification | 1/17 | 71/11590 | 0,099 | 0,252 | 0,210 | APOE | 1 |
| GO:0006641 | triglyceride metabolic process | 1/17 | 72/11590 | 0,101 | 0,252 | 0,210 | APOE | 1 |
| GO:0010770 | positive regulation of cell morphogenesis involved in differentiation | 1/17 | 72/11590 | 0,101 | 0,252 | 0,210 | STAU2 | 1 |
| GO:0051963 | regulation of synapse assembly | 1/17 | 72/11590 | 0,101 | 0,252 | 0,210 | STAU2 | 1 |
| GO:0010975 | regulation of neuron projection development | 2/17 | 371/11590 | 0,101 | 0,252 | 0,210 | APOE/STAU2 | 2 |
| GO:0015914 | phospholipid transport | 1/17 | 73/11590 | 0,102 | 0,252 | 0,210 | APOE | 1 |
| GO:0033555 | multicellular organismal response to stress | 1/17 | 73/11590 | 0,102 | 0,252 | 0,210 | APOE | 1 |
| GO:0032370 | positive regulation of lipid transport | 1/17 | 74/11590 | 0,103 | 0,252 | 0,211 | APOE | 1 |
| GO:0035023 | regulation of Rho protein signal transduction | 1/17 | 74/11590 | 0,103 | 0,252 | 0,211 | APOE | 1 |
| GO:0060291 | long-term synaptic potentiation | 1/17 | 74/11590 | 0,103 | 0,252 | 0,211 | APOE | 1 |
| GO:0019935 | cyclic-nucleotide-mediated signaling | 1/17 | 77/11590 | 0,107 | 0,257 | 0,215 | APOE | 1 |
| GO:0048041 | focal adhesion assembly | 1/17 | 77/11590 | 0,107 | 0,257 | 0,215 | STON1 | 1 |
| GO:1903035 | negative regulation of response to wounding | 1/17 | 77/11590 | 0,107 | 0,257 | 0,215 | APOE | 1 |
| GO:0003333 | amino acid transmembrane transport | 1/17 | 78/11590 | 0,109 | 0,257 | 0,215 | SLC38A1 | 1 |
| GO:0030301 | cholesterol transport | 1/17 | 78/11590 | 0,109 | 0,257 | 0,215 | APOE | 1 |

| | | | | | | | | |
|---|---|---|---|---|---|---|---|---|
| GO:0015908 | fatty acid transport | 1/17 | 79/11590 | 0,110 | 0,257 | 0,215 | APOE | 1 |
| GO:0030516 | regulation of axon extension | 1/17 | 79/11590 | 0,110 | 0,257 | 0,215 | APOE | 1 |
| GO:0032092 | positive regulation of protein binding | 1/17 | 79/11590 | 0,110 | 0,257 | 0,215 | APOE | 1 |
| GO:0044070 | regulation of anion transport | 1/17 | 79/11590 | 0,110 | 0,257 | 0,215 | SLC38A1 | 1 |
| GO:0045833 | negative regulation of lipid metabolic process | 1/17 | 79/11590 | 0,110 | 0,257 | 0,215 | APOE | 1 |
| GO:0009791 | post-embryonic development | 1/17 | 80/11590 | 0,111 | 0,259 | 0,217 | EFEMP1 | 1 |
| GO:0120031 | plasma membrane bounded cell projection assembly | 2/17 | 393/11590 | 0,112 | 0,259 | 0,217 | STON1/STAU2 | 2 |
| GO:0019218 | regulation of steroid metabolic process | 1/17 | 81/11590 | 0,112 | 0,259 | 0,217 | APOE | 1 |
| GO:0045807 | positive regulation of endocytosis | 1/17 | 81/11590 | 0,112 | 0,259 | 0,217 | APOE | 1 |
| GO:1901890 | positive regulation of cell junction assembly | 1/17 | 82/11590 | 0,114 | 0,259 | 0,217 | STAU2 | 1 |
| GO:1902414 | protein localization to cell junction | 1/17 | 82/11590 | 0,114 | 0,259 | 0,217 | STAU2 | 1 |
| GO:0030316 | osteoclast differentiation | 1/17 | 83/11590 | 0,115 | 0,259 | 0,217 | SH3PXD2A | 1 |
| GO:0050773 | regulation of dendrite development | 1/17 | 83/11590 | 0,115 | 0,259 | 0,217 | STAU2 | 1 |
| GO:0030031 | cell projection assembly | 2/17 | 401/11590 | 0,115 | 0,259 | 0,217 | STON1/STAU2 | 2 |
| GO:0007264 | small GTPase mediated signal transduction | 2/17 | 402/11590 | 0,116 | 0,259 | 0,217 | APOE/ARHGEF12 | 2 |
| GO:0051341 | regulation of oxidoreductase activity | 1/17 | 84/11590 | 0,116 | 0,259 | 0,217 | APOE | 1 |
| GO:0062014 | negative regulation of small molecule metabolic process | 1/17 | 84/11590 | 0,116 | 0,259 | 0,217 | APOE | 1 |
| GO:1990748 | cellular detoxification | 1/17 | 84/11590 | 0,116 | 0,259 | 0,217 | APOE | 1 |
| GO:0043535 | regulation of blood vessel endothelial cell migration | 1/17 | 85/11590 | 0,118 | 0,260 | 0,218 | APOE | 1 |
| GO:0051952 | regulation of amine transport | 1/17 | 85/11590 | 0,118 | 0,260 | 0,218 | SLC38A1 | 1 |
| GO:0007044 | cell-substrate junction assembly | 1/17 | 87/11590 | 0,120 | 0,265 | 0,221 | STON1 | 1 |
| GO:0010769 | regulation of cell morphogenesis involved in differentiation | 1/17 | 88/11590 | 0,122 | 0,267 | 0,223 | STAU2 | 1 |
| GO:0051224 | negative regulation of protein transport | 1/17 | 89/11590 | 0,123 | 0,267 | 0,224 | APOE | 1 |
| GO:0060840 | artery development | 1/17 | 89/11590 | 0,123 | 0,267 | 0,224 | APOE | 1 |

| | | | | | | | | |
|---|---|---|---|---|---|---|---|---|
| GO:0015918 | sterol transport | 1/17 | 90/11590 | 0,124 | 0,267 | 0,224 | APOE | 1 |
| GO:0033865 | nucleoside bisphosphate metabolic process | 1/17 | 91/11590 | 0,125 | 0,267 | 0,224 | DCAKD | 1 |
| GO:0033875 | ribonucleoside bisphosphate metabolic process | 1/17 | 91/11590 | 0,125 | 0,267 | 0,224 | DCAKD | 1 |
| GO:0034032 | purine nucleoside bisphosphate metabolic process | 1/17 | 91/11590 | 0,125 | 0,267 | 0,224 | DCAKD | 1 |
| GO:0097237 | cellular response to toxic substance | 1/17 | 91/11590 | 0,125 | 0,267 | 0,224 | APOE | 1 |
| GO:1905954 | positive regulation of lipid localization | 1/17 | 91/11590 | 0,125 | 0,267 | 0,224 | APOE | 1 |
| GO:0015837 | amine transport | 1/17 | 92/11590 | 0,127 | 0,268 | 0,224 | SLC38A1 | 1 |
| GO:0006639 | acylglycerol metabolic process | 1/17 | 93/11590 | 0,128 | 0,268 | 0,224 | APOE | 1 |
| GO:0007173 | epidermal growth factor receptor signaling pathway | 1/17 | 93/11590 | 0,128 | 0,268 | 0,224 | EFEMP1 | 1 |
| GO:0150115 | cell-substrate junction organization | 1/17 | 93/11590 | 0,128 | 0,268 | 0,224 | STON1 | 1 |
| GO:1904950 | negative regulation of establishment of protein localization | 1/17 | 93/11590 | 0,128 | 0,268 | 0,224 | APOE | 1 |
| GO:0006638 | neutral lipid metabolic process | 1/17 | 94/11590 | 0,129 | 0,269 | 0,225 | APOE | 1 |
| GO:0061387 | regulation of extent of cell growth | 1/17 | 94/11590 | 0,129 | 0,269 | 0,225 | APOE | 1 |
| GO:0051702 | biological process involved in interaction with symbiont | 1/17 | 95/11590 | 0,131 | 0,270 | 0,225 | APOE | 1 |
| GO:0071901 | negative regulation of protein serine/threonine kinase activity | 1/17 | 95/11590 | 0,131 | 0,270 | 0,225 | APOE | 1 |
| GO:0002062 | chondrocyte differentiation | 1/17 | 96/11590 | 0,132 | 0,271 | 0,227 | EFEMP1 | 1 |
| GO:0060627 | regulation of vesicle-mediated transport | 2/17 | 440/11590 | 0,135 | 0,276 | 0,230 | APOE/STON1 | 2 |
| GO:0007613 | memory | 1/17 | 99/11590 | 0,136 | 0,276 | 0,231 | APOE | 1 |
| GO:0042157 | lipoprotein metabolic process | 1/17 | 99/11590 | 0,136 | 0,276 | 0,231 | APOE | 1 |
| GO:0090090 | negative regulation of canonical Wnt signaling pathway | 1/17 | 100/11590 | 0,137 | 0,278 | 0,232 | APOE | 1 |
| GO:0015748 | organophosphate ester transport | 1/17 | 101/11590 | 0,138 | 0,280 | 0,234 | APOE | 1 |
| GO:0001501 | skeletal system development | 2/17 | 450/11590 | 0,140 | 0,281 | 0,235 | EFEMP1/VCAN | 2 |
| GO:0035303 | regulation of dephosphorylation | 1/17 | 103/11590 | 0,141 | 0,282 | 0,236 | MTMR9 | 1 |

| | | | | | | | | |
|---|---|---|---|---|---|---|---|---|
| GO:0007423 | sensory organ development | 2/17 | 453/11590 | 0,141 | 0,282 | 0,236 | EFEMP1/STAU2 | 2 |
| GO:0098754 | detoxification | 1/17 | 105/11590 | 0,143 | 0,286 | 0,239 | APOE | 1 |
| GO:0038127 | ERBB signaling pathway | 1/17 | 106/11590 | 0,145 | 0,286 | 0,239 | EFEMP1 | 1 |
| GO:0048675 | axon extension | 1/17 | 106/11590 | 0,145 | 0,286 | 0,239 | APOE | 1 |
| GO:0048667 | cell morphogenesis involved in neuron differentiation | 2/17 | 461/11590 | 0,145 | 0,286 | 0,239 | APOE/STAU2 | 2 |
| GO:0044283 | small molecule biosynthetic process | 2/17 | 470/11590 | 0,150 | 0,293 | 0,245 | APOE/SLC38A1 | 2 |
| GO:0032368 | regulation of lipid transport | 1/17 | 110/11590 | 0,150 | 0,293 | 0,245 | APOE | 1 |
| GO:0043534 | blood vessel endothelial cell migration | 1/17 | 110/11590 | 0,150 | 0,293 | 0,245 | APOE | 1 |
| GO:0007030 | Golgi organization | 1/17 | 111/11590 | 0,151 | 0,294 | 0,246 | COG2 | 1 |
| GO:0006865 | amino acid transport | 1/17 | 115/11590 | 0,156 | 0,303 | 0,253 | SLC38A1 | 1 |
| GO:0010977 | negative regulation of neuron projection development | 1/17 | 117/11590 | 0,159 | 0,304 | 0,255 | APOE | 1 |
| GO:0008203 | cholesterol metabolic process | 1/17 | 118/11590 | 0,160 | 0,304 | 0,255 | APOE | 1 |
| GO:0030168 | platelet activation | 1/17 | 118/11590 | 0,160 | 0,304 | 0,255 | APOE | 1 |
| GO:1903825 | organic acid transmembrane transport | 1/17 | 118/11590 | 0,160 | 0,304 | 0,255 | SLC38A1 | 1 |
| GO:1905039 | carboxylic acid transmembrane transport | 1/17 | 118/11590 | 0,160 | 0,304 | 0,255 | SLC38A1 | 1 |
| GO:0046165 | alcohol biosynthetic process | 1/17 | 119/11590 | 0,161 | 0,304 | 0,255 | APOE | 1 |
| GO:0048813 | dendrite morphogenesis | 1/17 | 119/11590 | 0,161 | 0,304 | 0,255 | STAU2 | 1 |
| GO:0061041 | regulation of wound healing | 1/17 | 119/11590 | 0,161 | 0,304 | 0,255 | APOE | 1 |
| GO:0001936 | regulation of endothelial cell proliferation | 1/17 | 120/11590 | 0,162 | 0,305 | 0,255 | APOE | 1 |
| GO:0030178 | negative regulation of Wnt signaling pathway | 1/17 | 121/11590 | 0,164 | 0,305 | 0,255 | APOE | 1 |
| GO:0055088 | lipid homeostasis | 1/17 | 121/11590 | 0,164 | 0,305 | 0,255 | APOE | 1 |
| GO:1903531 | negative regulation of secretion by cell | 1/17 | 121/11590 | 0,164 | 0,305 | 0,255 | APOE | 1 |
| GO:0050728 | negative regulation of inflammatory response | 1/17 | 122/11590 | 0,165 | 0,307 | 0,256 | APOE | 1 |
| GO:0062013 | positive regulation of small molecule metabolic process | 1/17 | 123/11590 | 0,166 | 0,308 | 0,257 | APOE | 1 |

| | | | | | | | | |
|---|---|---|---|---|---|---|---|---|
| GO:0050680 | negative regulation of epithelial cell proliferation | 1/17 | 124/11590 | 0,167 | 0,309 | 0,258 | APOE | 1 |
| GO:0008037 | cell recognition | 1/17 | 125/11590 | 0,168 | 0,311 | 0,260 | VCAN | 1 |
| GO:1902652 | secondary alcohol metabolic process | 1/17 | 126/11590 | 0,170 | 0,312 | 0,261 | APOE | 1 |
| GO:0010970 | transport along microtubule | 1/17 | 128/11590 | 0,172 | 0,313 | 0,262 | STAU2 | 1 |
| GO:0045834 | positive regulation of lipid metabolic process | 1/17 | 128/11590 | 0,172 | 0,313 | 0,262 | APOE | 1 |
| GO:0046488 | phosphatidylinositol metabolic process | 1/17 | 128/11590 | 0,172 | 0,313 | 0,262 | MTMR9 | 1 |
| GO:0016125 | sterol metabolic process | 1/17 | 129/11590 | 0,173 | 0,315 | 0,263 | APOE | 1 |
| GO:0007416 | synapse assembly | 1/17 | 131/11590 | 0,176 | 0,316 | 0,265 | STAU2 | 1 |
| GO:1902904 | negative regulation of supramolecular fiber organization | 1/17 | 131/11590 | 0,176 | 0,316 | 0,265 | APOE | 1 |
| GO:0043524 | negative regulation of neuron apoptotic process | 1/17 | 132/11590 | 0,177 | 0,316 | 0,265 | APOE | 1 |
| GO:0048660 | regulation of smooth muscle cell proliferation | 1/17 | 132/11590 | 0,177 | 0,316 | 0,265 | APOE | 1 |
| GO:0050806 | positive regulation of synaptic transmission | 1/17 | 132/11590 | 0,177 | 0,316 | 0,265 | APOE | 1 |
| GO:1905952 | regulation of lipid localization | 1/17 | 133/11590 | 0,178 | 0,318 | 0,266 | APOE | 1 |
| GO:0001935 | endothelial cell proliferation | 1/17 | 134/11590 | 0,179 | 0,318 | 0,266 | APOE | 1 |
| GO:0035150 | regulation of tube size | 1/17 | 135/11590 | 0,181 | 0,318 | 0,266 | APOE | 1 |
| GO:0035296 | regulation of tube diameter | 1/17 | 135/11590 | 0,181 | 0,318 | 0,266 | APOE | 1 |
| GO:0097746 | blood vessel diameter maintenance | 1/17 | 135/11590 | 0,181 | 0,318 | 0,266 | APOE | 1 |
| GO:0048659 | smooth muscle cell proliferation | 1/17 | 136/11590 | 0,182 | 0,319 | 0,267 | APOE | 1 |
| GO:0043409 | negative regulation of MAPK cascade | 1/17 | 137/11590 | 0,183 | 0,320 | 0,268 | APOE | 1 |
| GO:0031503 | protein-containing complex localization | 1/17 | 138/11590 | 0,184 | 0,322 | 0,269 | STAU2 | 1 |
| GO:0051048 | negative regulation of secretion | 1/17 | 139/11590 | 0,186 | 0,323 | 0,270 | APOE | 1 |
| GO:0046890 | regulation of lipid biosynthetic process | 1/17 | 141/11590 | 0,188 | 0,325 | 0,272 | APOE | 1 |
| GO:1904064 | positive regulation of cation transmembrane transport | 1/17 | 141/11590 | 0,188 | 0,325 | 0,272 | SLC38A1 | 1 |

| | | | | | | | | |
|---|---|---|---|---|---|---|---|---|
| GO:1990138 | neuron projection extension | 1/17 | 144/11590 | 0,192 | 0,330 | 0,276 | APOE | 1 |
| GO:1903034 | regulation of response to wounding | 1/17 | 146/11590 | 0,194 | 0,334 | 0,279 | APOE | 1 |
| GO:1903828 | negative regulation of protein localization | 1/17 | 147/11590 | 0,195 | 0,335 | 0,280 | APOE | 1 |
| GO:0034249 | negative regulation of cellular amide metabolic process | 1/17 | 150/11590 | 0,199 | 0,338 | 0,283 | APOE | 1 |
| GO:0099111 | microtubule-based transport | 1/17 | 150/11590 | 0,199 | 0,338 | 0,283 | STAU2 | 1 |
| GO:0006694 | steroid biosynthetic process | 1/17 | 151/11590 | 0,200 | 0,338 | 0,283 | APOE | 1 |
| GO:0010594 | regulation of endothelial cell migration | 1/17 | 151/11590 | 0,200 | 0,338 | 0,283 | APOE | 1 |
| GO:0061136 | regulation of proteasomal protein catabolic process | 1/17 | 152/11590 | 0,201 | 0,338 | 0,283 | APOE | 1 |
| GO:0099504 | synaptic vesicle cycle | 1/17 | 152/11590 | 0,201 | 0,338 | 0,283 | STON1 | 1 |
| GO:1902905 | positive regulation of supramolecular fiber organization | 1/17 | 152/11590 | 0,201 | 0,338 | 0,283 | APOE | 1 |
| GO:0034767 | positive regulation of ion transmembrane transport | 1/17 | 153/11590 | 0,202 | 0,339 | 0,283 | SLC38A1 | 1 |
| GO:0031345 | negative regulation of cell projection organization | 1/17 | 155/11590 | 0,205 | 0,341 | 0,285 | APOE | 1 |
| GO:0051099 | positive regulation of binding | 1/17 | 155/11590 | 0,205 | 0,341 | 0,285 | APOE | 1 |
| GO:0008361 | regulation of cell size | 1/17 | 158/11590 | 0,208 | 0,344 | 0,288 | APOE | 1 |
| GO:0030705 | cytoskeleton-dependent intracellular transport | 1/17 | 158/11590 | 0,208 | 0,344 | 0,288 | STAU2 | 1 |
| GO:0120032 | regulation of plasma membrane bounded cell projection assembly | 1/17 | 158/11590 | 0,208 | 0,344 | 0,288 | STAU2 | 1 |
| GO:0046578 | regulation of Ras protein signal transduction | 1/17 | 159/11590 | 0,209 | 0,344 | 0,288 | APOE | 1 |
| GO:0060491 | regulation of cell projection assembly | 1/17 | 159/11590 | 0,209 | 0,344 | 0,288 | STAU2 | 1 |
| GO:1901888 | regulation of cell junction assembly | 1/17 | 161/11590 | 0,212 | 0,347 | 0,290 | STAU2 | 1 |
| GO:0043405 | regulation of MAP kinase activity | 1/17 | 164/11590 | 0,215 | 0,350 | 0,292 | APOE | 1 |
| GO:0045732 | positive regulation of protein catabolic process | 1/17 | 164/11590 | 0,215 | 0,350 | 0,292 | APOE | 1 |
| GO:0070085 | glycosylation | 1/17 | 164/11590 | 0,215 | 0,350 | 0,292 | COG2 | 1 |

| | | | | | | | | |
|---|---|---|---|---|---|---|---|---|
| GO:0098739 | import across plasma membrane | 1/17 | 165/11590 | 0,216 | 0,350 | 0,292 | SLC38A1 | 1 |
| GO:1901605 | alpha-amino acid metabolic process | 1/17 | 165/11590 | 0,216 | 0,350 | 0,292 | SLC38A1 | 1 |
| GO:0007601 | visual perception | 1/17 | 166/11590 | 0,218 | 0,350 | 0,293 | EFEMP1 | 1 |
| GO:0006403 | RNA localization | 1/17 | 167/11590 | 0,219 | 0,350 | 0,293 | STAU2 | 1 |
| GO:0006469 | negative regulation of protein kinase activity | 1/17 | 167/11590 | 0,219 | 0,350 | 0,293 | APOE | 1 |
| GO:0007565 | female pregnancy | 1/17 | 168/11590 | 0,220 | 0,350 | 0,293 | SLC38A1 | 1 |
| GO:0050953 | sensory perception of light stimulus | 1/17 | 168/11590 | 0,220 | 0,350 | 0,293 | EFEMP1 | 1 |
| GO:0007626 | locomotory behavior | 1/17 | 169/11590 | 0,221 | 0,350 | 0,293 | APOE | 1 |
| GO:0099003 | vesicle-mediated transport in synapse | 1/17 | 169/11590 | 0,221 | 0,350 | 0,293 | STON1 | 1 |
| GO:0000302 | response to reactive oxygen species | 1/17 | 170/11590 | 0,222 | 0,350 | 0,293 | APOE | 1 |
| GO:0043393 | regulation of protein binding | 1/17 | 170/11590 | 0,222 | 0,350 | 0,293 | APOE | 1 |
| GO:0051216 | cartilage development | 1/17 | 172/11590 | 0,225 | 0,353 | 0,295 | EFEMP1 | 1 |
| GO:0030258 | lipid modification | 1/17 | 173/11590 | 0,226 | 0,354 | 0,296 | MTMR9 | 1 |
| GO:0009152 | purine ribonucleotide biosynthetic process | 1/17 | 175/11590 | 0,228 | 0,357 | 0,298 | DCAKD | 1 |
| GO:0050821 | protein stabilization | 1/17 | 177/11590 | 0,230 | 0,359 | 0,300 | MTMR9 | 1 |
| GO:0050866 | negative regulation of cell activation | 1/17 | 178/11590 | 0,231 | 0,359 | 0,300 | APOE | 1 |
| GO:1903050 | regulation of proteolysis involved in protein catabolic process | 1/17 | 178/11590 | 0,231 | 0,359 | 0,300 | APOE | 1 |
| GO:0006836 | neurotransmitter transport | 1/17 | 179/11590 | 0,233 | 0,359 | 0,300 | SLC38A1 | 1 |
| GO:0070374 | positive regulation of ERK1 and ERK2 cascade | 1/17 | 179/11590 | 0,233 | 0,359 | 0,300 | APOE | 1 |
| GO:0002573 | myeloid leukocyte differentiation | 1/17 | 181/11590 | 0,235 | 0,361 | 0,302 | SH3PXD2A | 1 |
| GO:1901215 | negative regulation of neuron death | 1/17 | 181/11590 | 0,235 | 0,361 | 0,302 | APOE | 1 |
| GO:0001505 | regulation of neurotransmitter levels | 1/17 | 185/11590 | 0,239 | 0,365 | 0,305 | SLC38A1 | 1 |
| GO:0009260 | ribonucleotide biosynthetic process | 1/17 | 186/11590 | 0,241 | 0,365 | 0,305 | DCAKD | 1 |
| GO:0033673 | negative regulation of kinase activity | 1/17 | 186/11590 | 0,241 | 0,365 | 0,305 | APOE | 1 |
| GO:0044703 | multi-organism reproductive process | 1/17 | 186/11590 | 0,241 | 0,365 | 0,305 | SLC38A1 | 1 |
| GO:0051651 | maintenance of location in cell | 1/17 | 186/11590 | 0,241 | 0,365 | 0,305 | APOE | 1 |

| | | | | | | | | |
|---|---|---|---|---|---|---|---|---|
| GO:0033002 | muscle cell proliferation | 1/17 | 187/11590 | 0,242 | 0,365 | 0,305 | APOE | 1 |
| GO:0060828 | regulation of canonical Wnt signaling pathway | 1/17 | 188/11590 | 0,243 | 0,366 | 0,306 | APOE | 1 |
| GO:0006814 | sodium ion transport | 1/17 | 189/11590 | 0,244 | 0,366 | 0,306 | SLC38A1 | 1 |
| GO:0043523 | regulation of neuron apoptotic process | 1/17 | 189/11590 | 0,244 | 0,366 | 0,306 | APOE | 1 |
| GO:0072593 | reactive oxygen species metabolic process | 1/17 | 190/11590 | 0,245 | 0,366 | 0,306 | SH3PXD2A | 1 |
| GO:0048588 | developmental cell growth | 1/17 | 191/11590 | 0,246 | 0,366 | 0,306 | APOE | 1 |
| GO:0031330 | negative regulation of cellular catabolic process | 1/17 | 192/11590 | 0,247 | 0,366 | 0,306 | MTMR9 | 1 |
| GO:0044706 | multi-multicellular organism process | 1/17 | 192/11590 | 0,247 | 0,366 | 0,306 | SLC38A1 | 1 |
| GO:0006164 | purine nucleotide biosynthetic process | 1/17 | 193/11590 | 0,248 | 0,366 | 0,306 | DCAKD | 1 |
| GO:0045088 | regulation of innate immune response | 1/17 | 193/11590 | 0,248 | 0,366 | 0,306 | APOE | 1 |
| GO:0046390 | ribose phosphate biosynthetic process | 1/17 | 193/11590 | 0,248 | 0,366 | 0,306 | DCAKD | 1 |
| GO:0009636 | response to toxic substance | 1/17 | 194/11590 | 0,250 | 0,367 | 0,307 | APOE | 1 |
| GO:0001649 | osteoblast differentiation | 1/17 | 195/11590 | 0,251 | 0,367 | 0,307 | VCAN | 1 |
| GO:0007281 | germ cell development | 1/17 | 195/11590 | 0,251 | 0,367 | 0,307 | STAU2 | 1 |
| GO:0060560 | developmental growth involved in morphogenesis | 1/17 | 196/11590 | 0,252 | 0,367 | 0,307 | APOE | 1 |
| GO:0034764 | positive regulation of transmembrane transport | 1/17 | 198/11590 | 0,254 | 0,368 | 0,307 | SLC38A1 | 1 |
| GO:0043542 | endothelial cell migration | 1/17 | 198/11590 | 0,254 | 0,368 | 0,307 | APOE | 1 |
| GO:0031348 | negative regulation of defense response | 1/17 | 199/11590 | 0,255 | 0,368 | 0,307 | APOE | 1 |
| GO:0050769 | positive regulation of neurogenesis | 1/17 | 199/11590 | 0,255 | 0,368 | 0,307 | STAU2 | 1 |
| GO:0072522 | purine-containing compound biosynthetic process | 1/17 | 199/11590 | 0,255 | 0,368 | 0,307 | DCAKD | 1 |
| GO:0007596 | blood coagulation | 1/17 | 200/11590 | 0,256 | 0,368 | 0,308 | APOE | 1 |
| GO:0006354 | DNA-templated transcription elongation | 1/17 | 202/11590 | 0,259 | 0,370 | 0,309 | FHL5 | 1 |
| GO:0098656 | anion transmembrane transport | 1/17 | 202/11590 | 0,259 | 0,370 | 0,309 | SLC38A1 | 1 |

| | | | | | | | | |
|---|---|---|---|---|---|---|---|---|
| GO:0050817 | coagulation | 1/17 | 203/11590 | 0,260 | 0,371 | 0,310 | APOE | 1 |
| GO:0007599 | hemostasis | 1/17 | 205/11590 | 0,262 | 0,373 | 0,312 | APOE | 1 |
| GO:0007160 | cell-matrix adhesion | 1/17 | 206/11590 | 0,263 | 0,373 | 0,312 | STON1 | 1 |
| GO:0010632 | regulation of epithelial cell migration | 1/17 | 207/11590 | 0,264 | 0,374 | 0,313 | APOE | 1 |
| GO:0051348 | negative regulation of transferase activity | 1/17 | 208/11590 | 0,265 | 0,375 | 0,313 | APOE | 1 |
| GO:0050708 | regulation of protein secretion | 1/17 | 210/11590 | 0,267 | 0,375 | 0,314 | APOE | 1 |
| GO:0098657 | import into cell | 1/17 | 210/11590 | 0,267 | 0,375 | 0,314 | SLC38A1 | 1 |
| GO:1901617 | organic hydroxy compound biosynthetic process | 1/17 | 210/11590 | 0,267 | 0,375 | 0,314 | APOE | 1 |
| GO:0006898 | receptor-mediated endocytosis | 1/17 | 214/11590 | 0,272 | 0,379 | 0,317 | APOE | 1 |
| GO:0015850 | organic hydroxy compound transport | 1/17 | 214/11590 | 0,272 | 0,379 | 0,317 | APOE | 1 |
| GO:0046942 | carboxylic acid transport | 1/17 | 220/11590 | 0,278 | 0,388 | 0,324 | SLC38A1 | 1 |
| GO:0051402 | neuron apoptotic process | 1/17 | 223/11590 | 0,281 | 0,391 | 0,327 | APOE | 1 |
| GO:0048562 | embryonic organ morphogenesis | 1/17 | 228/11590 | 0,287 | 0,397 | 0,332 | EFEMP1 | 1 |
| GO:0048193 | Golgi vesicle transport | 1/17 | 229/11590 | 0,288 | 0,397 | 0,332 | COG2 | 1 |
| GO:0006520 | cellular amino acid metabolic process | 1/17 | 231/11590 | 0,290 | 0,397 | 0,332 | SLC38A1 | 1 |
| GO:0034599 | cellular response to oxidative stress | 1/17 | 231/11590 | 0,290 | 0,397 | 0,332 | STAU2 | 1 |
| GO:0051962 | positive regulation of nervous system development | 1/17 | 231/11590 | 0,290 | 0,397 | 0,332 | STAU2 | 1 |
| GO:0060070 | canonical Wnt signaling pathway | 1/17 | 232/11590 | 0,291 | 0,397 | 0,332 | APOE | 1 |
| GO:0007611 | learning or memory | 1/17 | 233/11590 | 0,292 | 0,397 | 0,332 | APOE | 1 |
| GO:0009165 | nucleotide biosynthetic process | 1/17 | 233/11590 | 0,292 | 0,397 | 0,332 | DCAKD | 1 |
| GO:0061448 | connective tissue development | 1/17 | 233/11590 | 0,292 | 0,397 | 0,332 | EFEMP1 | 1 |
| GO:0006650 | glycerophospholipid metabolic process | 1/17 | 234/11590 | 0,293 | 0,397 | 0,332 | MTMR9 | 1 |
| GO:1901293 | nucleoside phosphate biosynthetic process | 1/17 | 234/11590 | 0,293 | 0,397 | 0,332 | DCAKD | 1 |
| GO:0006874 | cellular calcium ion homeostasis | 1/17 | 235/11590 | 0,294 | 0,398 | 0,333 | APOE | 1 |
| GO:0030111 | regulation of Wnt signaling pathway | 1/17 | 243/11590 | 0,303 | 0,406 | 0,340 | APOE | 1 |
| GO:0043270 | positive regulation of ion transport | 1/17 | 243/11590 | 0,303 | 0,406 | 0,340 | SLC38A1 | 1 |

| | | | | | | | | |
|---|---|---|---|---|---|---|---|---|
| GO:0070372 | regulation of ERK1 and ERK2 cascade | 1/17 | 243/11590 | 0,303 | 0,406 | 0,340 | APOE | 1 |
| GO:0016042 | lipid catabolic process | 1/17 | 246/11590 | 0,306 | 0,410 | 0,342 | APOE | 1 |
| GO:0030336 | negative regulation of cell migration | 1/17 | 247/11590 | 0,307 | 0,410 | 0,343 | APOE | 1 |
| GO:0046394 | carboxylic acid biosynthetic process | 1/17 | 250/11590 | 0,310 | 0,413 | 0,345 | SLC38A1 | 1 |
| GO:0016053 | organic acid biosynthetic process | 1/17 | 252/11590 | 0,312 | 0,414 | 0,346 | SLC38A1 | 1 |
| GO:0022604 | regulation of cell morphogenesis | 1/17 | 252/11590 | 0,312 | 0,414 | 0,346 | STAU2 | 1 |
| GO:0055074 | calcium ion homeostasis | 1/17 | 256/11590 | 0,316 | 0,418 | 0,349 | APOE | 1 |
| GO:0007018 | microtubule-based movement | 1/17 | 257/11590 | 0,317 | 0,418 | 0,349 | STAU2 | 1 |
| GO:0044403 | biological process involved in symbiotic interaction | 1/17 | 257/11590 | 0,317 | 0,418 | 0,349 | APOE | 1 |
| GO:0001933 | negative regulation of protein phosphorylation | 1/17 | 259/11590 | 0,319 | 0,418 | 0,349 | APOE | 1 |
| GO:0009895 | negative regulation of catabolic process | 1/17 | 259/11590 | 0,319 | 0,418 | 0,349 | MTMR9 | 1 |
| GO:2000146 | negative regulation of cell motility | 1/17 | 259/11590 | 0,319 | 0,418 | 0,349 | APOE | 1 |
| GO:0050890 | cognition | 1/17 | 262/11590 | 0,322 | 0,420 | 0,351 | APOE | 1 |
| GO:0072503 | cellular divalent inorganic cation homeostasis | 1/17 | 262/11590 | 0,322 | 0,420 | 0,351 | APOE | 1 |
| GO:0031647 | regulation of protein stability | 1/17 | 264/11590 | 0,324 | 0,421 | 0,352 | MTMR9 | 1 |
| GO:0070371 | ERK1 and ERK2 cascade | 1/17 | 264/11590 | 0,324 | 0,421 | 0,352 | APOE | 1 |
| GO:0008202 | steroid metabolic process | 1/17 | 268/11590 | 0,328 | 0,424 | 0,354 | APOE | 1 |
| GO:0010720 | positive regulation of cell development | 1/17 | 269/11590 | 0,329 | 0,424 | 0,354 | STAU2 | 1 |
| GO:0016311 | dephosphorylation | 1/17 | 269/11590 | 0,329 | 0,424 | 0,354 | MTMR9 | 1 |
| GO:0019932 | second-messenger-mediated signaling | 1/17 | 269/11590 | 0,329 | 0,424 | 0,354 | APOE | 1 |
| GO:0048638 | regulation of developmental growth | 1/17 | 271/11590 | 0,331 | 0,424 | 0,354 | APOE | 1 |
| GO:0062012 | regulation of small molecule metabolic process | 1/17 | 271/11590 | 0,331 | 0,424 | 0,354 | APOE | 1 |
| GO:0010631 | epithelial cell migration | 1/17 | 272/11590 | 0,332 | 0,424 | 0,354 | APOE | 1 |
| GO:0019058 | viral life cycle | 1/17 | 272/11590 | 0,332 | 0,424 | 0,354 | APOE | 1 |
| GO:0051235 | maintenance of location | 1/17 | 274/11590 | 0,334 | 0,424 | 0,355 | APOE | 1 |

| | | | | | | | | |
|---|---|---|---|---|---|---|---|---|
| GO:0090132 | epithelium migration | 1/17 | 274/11590 | 0,334 | 0,424 | 0,355 | APOE | 1 |
| GO:0022412 | cellular process involved in reproduction in multicellular organism | 1/17 | 275/11590 | 0,335 | 0,424 | 0,355 | STAU2 | 1 |
| GO:0010506 | regulation of autophagy | 1/17 | 276/11590 | 0,336 | 0,424 | 0,355 | MTMR9 | 1 |
| GO:0015711 | organic anion transport | 1/17 | 276/11590 | 0,336 | 0,424 | 0,355 | SLC38A1 | 1 |
| GO:1901214 | regulation of neuron death | 1/17 | 278/11590 | 0,338 | 0,426 | 0,356 | APOE | 1 |
| GO:0090130 | tissue migration | 1/17 | 279/11590 | 0,339 | 0,426 | 0,356 | APOE | 1 |
| GO:0062197 | cellular response to chemical stress | 1/17 | 282/11590 | 0,342 | 0,429 | 0,358 | STAU2 | 1 |
| GO:0009306 | protein secretion | 1/17 | 283/11590 | 0,343 | 0,429 | 0,358 | APOE | 1 |
| GO:0043010 | camera-type eye development | 1/17 | 283/11590 | 0,343 | 0,429 | 0,358 | EFEMP1 | 1 |
| GO:0035592 | establishment of protein localization to extracellular region | 1/17 | 284/11590 | 0,344 | 0,429 | 0,359 | APOE | 1 |
| GO:0002831 | regulation of response to biotic stimulus | 1/17 | 285/11590 | 0,345 | 0,429 | 0,359 | APOE | 1 |
| GO:0040013 | negative regulation of locomotion | 1/17 | 288/11590 | 0,348 | 0,431 | 0,360 | APOE | 1 |
| GO:0042176 | regulation of protein catabolic process | 1/17 | 289/11590 | 0,349 | 0,431 | 0,360 | APOE | 1 |
| GO:0071692 | protein localization to extracellular region | 1/17 | 289/11590 | 0,349 | 0,431 | 0,360 | APOE | 1 |
| GO:0006644 | phospholipid metabolic process | 1/17 | 290/11590 | 0,350 | 0,431 | 0,360 | MTMR9 | 1 |
| GO:0072507 | divalent inorganic cation homeostasis | 1/17 | 290/11590 | 0,350 | 0,431 | 0,360 | APOE | 1 |
| GO:0032956 | regulation of actin cytoskeleton organization | 1/17 | 291/11590 | 0,351 | 0,431 | 0,360 | STAU2 | 1 |
| GO:1904062 | regulation of cation transmembrane transport | 1/17 | 293/11590 | 0,353 | 0,433 | 0,362 | SLC38A1 | 1 |
| GO:0050727 | regulation of inflammatory response | 1/17 | 295/11590 | 0,355 | 0,434 | 0,363 | APOE | 1 |
| GO:0032535 | regulation of cellular component size | 1/17 | 297/11590 | 0,357 | 0,435 | 0,363 | APOE | 1 |
| GO:0042326 | negative regulation of phosphorylation | 1/17 | 297/11590 | 0,357 | 0,435 | 0,363 | APOE | 1 |
| GO:0006066 | alcohol metabolic process | 1/17 | 302/11590 | 0,362 | 0,440 | 0,367 | APOE | 1 |
| GO:0045862 | positive regulation of proteolysis | 1/17 | 303/11590 | 0,363 | 0,440 | 0,368 | APOE | 1 |
| GO:1902903 | regulation of supramolecular fiber organization | 1/17 | 307/11590 | 0,367 | 0,444 | 0,371 | APOE | 1 |

| | | | | | | | | |
|---|---|---|---|---|---|---|---|---|
| GO:0050678 | regulation of epithelial cell proliferation | 1/17 | 309/11590 | 0,369 | 0,445 | 0,372 | APOE | 1 |
| GO:0006869 | lipid transport | 1/17 | 312/11590 | 0,371 | 0,445 | 0,372 | APOE | 1 |
| GO:0031331 | positive regulation of cellular catabolic process | 1/17 | 313/11590 | 0,372 | 0,445 | 0,372 | APOE | 1 |
| GO:0044282 | small molecule catabolic process | 1/17 | 313/11590 | 0,372 | 0,445 | 0,372 | APOE | 1 |
| GO:0050767 | regulation of neurogenesis | 1/17 | 313/11590 | 0,372 | 0,445 | 0,372 | STAU2 | 1 |
| GO:0071900 | regulation of protein serine/threonine kinase activity | 1/17 | 313/11590 | 0,372 | 0,445 | 0,372 | APOE | 1 |
| GO:0051098 | regulation of binding | 1/17 | 317/11590 | 0,376 | 0,448 | 0,374 | APOE | 1 |
| GO:0070997 | neuron death | 1/17 | 317/11590 | 0,376 | 0,448 | 0,374 | APOE | 1 |
| GO:0031589 | cell-substrate adhesion | 1/17 | 318/11590 | 0,377 | 0,448 | 0,374 | STON1 | 1 |
| GO:0001701 | in utero embryonic development | 1/17 | 321/11590 | 0,380 | 0,450 | 0,376 | SH3PXD2A | 1 |
| GO:0018108 | peptidyl-tyrosine phosphorylation | 1/17 | 325/11590 | 0,384 | 0,453 | 0,379 | EFEMP1 | 1 |
| GO:0043254 | regulation of protein-containing complex assembly | 1/17 | 325/11590 | 0,384 | 0,453 | 0,379 | APOE | 1 |
| GO:0018212 | peptidyl-tyrosine modification | 1/17 | 327/11590 | 0,385 | 0,453 | 0,379 | EFEMP1 | 1 |
| GO:0032970 | regulation of actin filament-based process | 1/17 | 327/11590 | 0,385 | 0,453 | 0,379 | STAU2 | 1 |
| GO:0030099 | myeloid cell differentiation | 1/17 | 328/11590 | 0,386 | 0,453 | 0,379 | SH3PXD2A | 1 |
| GO:0006875 | cellular metal ion homeostasis | 1/17 | 333/11590 | 0,391 | 0,457 | 0,382 | APOE | 1 |
| GO:0032102 | negative regulation of response to external stimulus | 1/17 | 333/11590 | 0,391 | 0,457 | 0,382 | APOE | 1 |
| GO:0051051 | negative regulation of transport | 1/17 | 335/11590 | 0,393 | 0,458 | 0,383 | APOE | 1 |
| GO:0050878 | regulation of body fluid levels | 1/17 | 341/11590 | 0,398 | 0,462 | 0,387 | APOE | 1 |
| GO:0045936 | negative regulation of phosphate metabolic process | 1/17 | 342/11590 | 0,399 | 0,462 | 0,387 | APOE | 1 |
| GO:0010563 | negative regulation of phosphorus metabolic process | 1/17 | 343/11590 | 0,400 | 0,462 | 0,387 | APOE | 1 |
| GO:0016055 | Wnt signaling pathway | 1/17 | 343/11590 | 0,400 | 0,462 | 0,387 | APOE | 1 |
| GO:0001558 | regulation of cell growth | 1/17 | 344/11590 | 0,401 | 0,462 | 0,387 | APOE | 1 |

| | | | | | | | | |
|---|---|---|---|---|---|---|---|---|
| GO:0198738 | cell-cell signaling by wnt | 1/17 | 344/11590 | 0,401 | 0,462 | 0,387 | APOE | 1 |
| GO:0010876 | lipid localization | 1/17 | 348/11590 | 0,405 | 0,466 | 0,389 | APOE | 1 |
| GO:0009150 | purine ribonucleotide metabolic process | 1/17 | 349/11590 | 0,406 | 0,466 | 0,389 | DCAKD | 1 |
| GO:0001503 | ossification | 1/17 | 352/11590 | 0,408 | 0,468 | 0,391 | VCAN | 1 |
| GO:0034248 | regulation of cellular amide metabolic process | 1/17 | 358/11590 | 0,414 | 0,473 | 0,396 | APOE | 1 |
| GO:0048568 | embryonic organ development | 1/17 | 361/11590 | 0,416 | 0,475 | 0,397 | EFEMP1 | 1 |
| GO:0016032 | viral process | 1/17 | 362/11590 | 0,417 | 0,475 | 0,397 | APOE | 1 |
| GO:0007409 | axonogenesis | 1/17 | 365/11590 | 0,420 | 0,477 | 0,398 | APOE | 1 |
| GO:0009259 | ribonucleotide metabolic process | 1/17 | 365/11590 | 0,420 | 0,477 | 0,398 | DCAKD | 1 |
| GO:0050673 | epithelial cell proliferation | 1/17 | 369/11590 | 0,423 | 0,480 | 0,401 | APOE | 1 |
| GO:0006163 | purine nucleotide metabolic process | 1/17 | 371/11590 | 0,425 | 0,481 | 0,402 | DCAKD | 1 |
| GO:0051960 | regulation of nervous system development | 1/17 | 372/11590 | 0,426 | 0,481 | 0,402 | STAU2 | 1 |
| GO:0019693 | ribose phosphate metabolic process | 1/17 | 373/11590 | 0,427 | 0,481 | 0,402 | DCAKD | 1 |
| GO:0042060 | wound healing | 1/17 | 375/11590 | 0,429 | 0,482 | 0,403 | APOE | 1 |
| GO:0001667 | ameboidal-type cell migration | 1/17 | 379/11590 | 0,432 | 0,485 | 0,405 | APOE | 1 |
| GO:0034765 | regulation of ion transmembrane transport | 1/17 | 382/11590 | 0,435 | 0,487 | 0,407 | SLC38A1 | 1 |
| GO:0006820 | anion transport | 1/17 | 386/11590 | 0,438 | 0,489 | 0,409 | SLC38A1 | 1 |
| GO:0010498 | proteasomal protein catabolic process | 1/17 | 386/11590 | 0,438 | 0,489 | 0,409 | APOE | 1 |
| GO:0030003 | cellular cation homeostasis | 1/17 | 392/11590 | 0,443 | 0,493 | 0,413 | APOE | 1 |
| GO:0072521 | purine-containing compound metabolic process | 1/17 | 395/11590 | 0,446 | 0,494 | 0,413 | DCAKD | 1 |
| GO:0009896 | positive regulation of catabolic process | 1/17 | 397/11590 | 0,447 | 0,494 | 0,413 | APOE | 1 |
| GO:0022411 | cellular component disassembly | 1/17 | 397/11590 | 0,447 | 0,494 | 0,413 | STON1 | 1 |
| GO:1901361 | organic cyclic compound catabolic process | 1/17 | 397/11590 | 0,447 | 0,494 | 0,413 | APOE | 1 |
| GO:0031400 | negative regulation of protein modification process | 1/17 | 398/11590 | 0,448 | 0,494 | 0,413 | APOE | 1 |

| | | | | | | | | |
|---|---|---|---|---|---|---|---|---|
| GO:0031667 | response to nutrient levels | 1/17 | 403/11590 | 0,452 | 0,498 | 0,416 | APOE | 1 |
| GO:0061564 | axon development | 1/17 | 404/11590 | 0,453 | 0,498 | 0,416 | APOE | 1 |
| GO:0006873 | cellular ion homeostasis | 1/17 | 406/11590 | 0,455 | 0,498 | 0,416 | APOE | 1 |
| GO:0016049 | cell growth | 1/17 | 407/11590 | 0,456 | 0,498 | 0,416 | APOE | 1 |
| GO:0044089 | positive regulation of cellular component biogenesis | 1/17 | 407/11590 | 0,456 | 0,498 | 0,416 | STAU2 | 1 |
| GO:0051223 | regulation of protein transport | 1/17 | 412/11590 | 0,460 | 0,501 | 0,419 | APOE | 1 |
| GO:0043410 | positive regulation of MAPK cascade | 1/17 | 413/11590 | 0,461 | 0,501 | 0,419 | APOE | 1 |
| GO:0010256 | endomembrane system organization | 1/17 | 414/11590 | 0,461 | 0,501 | 0,419 | COG2 | 1 |
| GO:1902532 | negative regulation of intracellular signal transduction | 1/17 | 415/11590 | 0,462 | 0,501 | 0,419 | APOE | 1 |
| GO:0055065 | metal ion homeostasis | 1/17 | 420/11590 | 0,466 | 0,505 | 0,422 | APOE | 1 |
| GO:0006914 | autophagy | 1/17 | 424/11590 | 0,470 | 0,507 | 0,423 | MTMR9 | 1 |
| GO:0061919 | process utilizing autophagic mechanism | 1/17 | 424/11590 | 0,470 | 0,507 | 0,423 | MTMR9 | 1 |
| GO:0090066 | regulation of anatomical structure size | 1/17 | 425/11590 | 0,470 | 0,507 | 0,423 | APOE | 1 |
| GO:0009991 | response to extracellular stimulus | 1/17 | 429/11590 | 0,474 | 0,509 | 0,426 | APOE | 1 |
| GO:0070201 | regulation of establishment of protein localization | 1/17 | 433/11590 | 0,477 | 0,511 | 0,428 | APOE | 1 |
| GO:0060284 | regulation of cell development | 1/17 | 434/11590 | 0,478 | 0,511 | 0,428 | STAU2 | 1 |
| GO:0051493 | regulation of cytoskeleton organization | 1/17 | 438/11590 | 0,481 | 0,514 | 0,430 | STAU2 | 1 |
| GO:1905114 | cell surface receptor signaling pathway involved in cell-cell signaling | 1/17 | 443/11590 | 0,485 | 0,517 | 0,432 | APOE | 1 |
| GO:0072657 | protein localization to membrane | 1/17 | 447/11590 | 0,488 | 0,520 | 0,434 | APOE | 1 |
| GO:0008015 | blood circulation | 1/17 | 452/11590 | 0,492 | 0,523 | 0,437 | APOE | 1 |
| GO:0034762 | regulation of transmembrane transport | 1/17 | 454/11590 | 0,493 | 0,524 | 0,438 | SLC38A1 | 1 |
| GO:0009117 | nucleotide metabolic process | 1/17 | 461/11590 | 0,499 | 0,527 | 0,441 | DCAKD | 1 |
| GO:0051345 | positive regulation of hydrolase activity | 1/17 | 461/11590 | 0,499 | 0,527 | 0,441 | MTMR9 | 1 |
| GO:1901615 | organic hydroxy compound metabolic process | 1/17 | 463/11590 | 0,500 | 0,528 | 0,441 | APOE | 1 |

| | | | | | | | | |
|---|---|---|---|---|---|---|---|---|
| GO:0007600 | sensory perception | 1/17 | 464/11590 | 0,501 | 0,528 | 0,441 | EFEMP1 | 1 |
| GO:0006753 | nucleoside phosphate metabolic process | 1/17 | 467/11590 | 0,503 | 0,529 | 0,443 | DCAKD | 1 |
| GO:0032787 | monocarboxylic acid metabolic process | 1/17 | 472/11590 | 0,507 | 0,532 | 0,445 | SLC38A1 | 1 |
| GO:0090407 | organophosphate biosynthetic process | 1/17 | 474/11590 | 0,509 | 0,533 | 0,446 | DCAKD | 1 |
| GO:0044057 | regulation of system process | 1/17 | 479/11590 | 0,512 | 0,535 | 0,447 | APOE | 1 |
| GO:0051347 | positive regulation of transferase activity | 1/17 | 480/11590 | 0,513 | 0,535 | 0,447 | APOE | 1 |
| GO:1903530 | regulation of secretion by cell | 1/17 | 480/11590 | 0,513 | 0,535 | 0,447 | APOE | 1 |
| GO:0055080 | cation homeostasis | 1/17 | 483/11590 | 0,515 | 0,536 | 0,448 | APOE | 1 |
| GO:0048598 | embryonic morphogenesis | 1/17 | 490/11590 | 0,520 | 0,540 | 0,451 | EFEMP1 | 1 |
| GO:0007276 | gamete generation | 1/17 | 491/11590 | 0,521 | 0,540 | 0,451 | STAU2 | 1 |
| GO:1901137 | carbohydrate derivative biosynthetic process | 1/17 | 491/11590 | 0,521 | 0,540 | 0,451 | DCAKD | 1 |
| GO:0009611 | response to wounding | 1/17 | 493/11590 | 0,523 | 0,540 | 0,452 | APOE | 1 |
| GO:0098771 | inorganic ion homeostasis | 1/17 | 496/11590 | 0,525 | 0,541 | 0,453 | APOE | 1 |
| GO:0006352 | DNA-templated transcription initiation | 0/17 | 115/11590 | 1,000 | 1,000 | 0,836 | | 0 |
| GO:0006367 | transcription initiation at RNA polymerase II promoter | 0/17 | 80/11590 | 1,000 | 1,000 | 0,836 | | 0 |
| GO:0007338 | single fertilization | 0/17 | 98/11590 | 1,000 | 1,000 | 0,836 | | 0 |
| GO:0007339 | binding of sperm to zona pellucida | 0/17 | 26/11590 | 1,000 | 1,000 | 0,836 | | 0 |
| GO:0009116 | nucleoside metabolic process | 0/17 | 44/11590 | 1,000 | 1,000 | 0,836 | | 0 |
| GO:0009119 | ribonucleoside metabolic process | 0/17 | 28/11590 | 1,000 | 1,000 | 0,836 | | 0 |
| GO:0009163 | nucleoside biosynthetic process | 0/17 | 11/11590 | 1,000 | 1,000 | 0,836 | | 0 |
| GO:0009566 | fertilization | 0/17 | 124/11590 | 1,000 | 1,000 | 0,836 | | 0 |
| GO:0009988 | cell-cell recognition | 0/17 | 47/11590 | 1,000 | 1,000 | 0,836 | | 0 |
| GO:0010508 | positive regulation of autophagy | 0/17 | 113/11590 | 1,000 | 1,000 | 0,836 | | 0 |
| GO:0034404 | nucleobase-containing small molecule biosynthetic process | 0/17 | 11/11590 | 1,000 | 1,000 | 0,836 | | 0 |
| GO:0035036 | sperm-egg recognition | 0/17 | 30/11590 | 1,000 | 1,000 | 0,836 | | 0 |

| | | | | | | | |
|---|---|---|---|---|---|---|---|
| GO:0042278 | purine nucleoside metabolic process | 0/17 | 20/11590 | 1,000 | 1,000 | 0,836 | 0 |
| GO:0043094 | cellular metabolic compound salvage | 0/17 | 22/11590 | 1,000 | 1,000 | 0,836 | 0 |
| GO:0043101 | purine-containing compound salvage | 0/17 | 12/11590 | 1,000 | 1,000 | 0,836 | 0 |
| GO:0046128 | purine ribonucleoside metabolic process | 0/17 | 17/11590 | 1,000 | 1,000 | 0,836 | 0 |
| GO:1901657 | glycosyl compound metabolic process | 0/17 | 62/11590 | 1,000 | 1,000 | 0,836 | 0 |
| GO:1901659 | glycosyl compound biosynthetic process | 0/17 | 12/11590 | 1,000 | 1,000 | 0,836 | 0 |

**Supplementary Table 6.** Enrichment analysis results for the SNPs-related genes of the PRS-WMH after the clumping.
*Legend: GeneRatio: ratio of input genes that are annotated in a term (GeneRatio=k/n, where k: overlap of the SNPs-related gene IDs with the specific gene set related to a biological pathway; n: size of the overlap of the SNPs-related gene IDs with all the members of the collection of gene sets); BgRatio: ratio of all genes that are annotated in a term (BgRatio=count/setSize, where count: number of genes that belong to a given gene-set; setSize: total number of genes in the collection of gene sets); P-value: probability of seeing at least X number of genes out of the total n SNPs-related gene IDs in the list annotated to a particular GO term, given the proportion of genes in the whole genome that are annotated to that GO term; FDR: p-value after false discovery rate correction; Q-value: proportion of false positives incurred when the test is significant (FDR correction among the significant results); Counts: number of genes annotated to the GO term.*
